# Longitudinal Metabolic Profiling of Women Using Selective Serotonin Reuptake Inhibitors During Pregnancy

**DOI:** 10.1101/2025.05.23.25328202

**Authors:** Anna Itkonen, Olli Kärkkäinen, Heidi Sahlman, Leea Keski-Nisula, Jaana Rysä

## Abstract

**Introduction:** Selective serotonin reuptake inhibitors (SSRIs) are the most prescribed antidepressants for pregnant women. While SSRIs are known to alter the circulating metabolic profile in non-pregnant individuals, the association between SSRIs and the changes in circulating metabolome during pregnancy remains unstudied. Pregnancy itself induces significant metabolic adjustments to meet the increased nutritional demands, and these maternal metabolic changes are crucial for the normal development and growth of the fetus.

**Objectives:** To study the impact of SSRI usage on circulating maternal metabolome during pregnancy.

**Methods:** A targeted nuclear magnetic resonance (NMR) spectroscopy method was used to analyze maternal serum samples obtained from the 1^st^ trimester of pregnancy and at the time of the delivery from both SSRI users (n =122) and non-depressive controls without antidepressants (n =117) for concentrations of metabolites and lipoproteins.

**Results:** During the 1^st^ trimester of pregnancy, SSRI usage was associated with increased lipid content in sixteen very low-density lipoprotein (VLDL) and chylomicron subtypes. At delivery, SSRI users exhibited alterations in lipoprotein lipid and fatty acid ratios. Similarly, while investigating the influence of SSRI usage on the pregnancy-driven changes in the metabolome, the interplay between pregnancy progression and SSRI usage lowered the lipoprotein lipid ratios.

**Conclusion:** Our analysis revealed a significant association between SSRIs and lipid metabolism. However, the observed changes were minor, suggesting a limited clinical impact. The findings enhance our understanding of the safe usage of SSRI medication during pregnancy.

## 1. Introduction

Pregnant women have an increased susceptibility to depression during and shortly after pregnancy (Gelaye et al. 2016). The most prescribed antidepressants during pregnancy are selective serotonin reuptake inhibitors (SSRIs) (Stephansson et al. 2013), with a worldwide prevalence estimate of 3% (Molenaar et al. 2020). Both depression and antidepressant usage may alter the metabolic profile and increase the risk for metabolic complications, as observed in non-pregnant adults and animal models (Beyazyüz et al. 2013; Bhattacharyya et al. 2019). Furthermore, pregnancy itself, antenatal depression, chemical exposures during pregnancy and pregnancy complications have been linked to altered maternal metabolic profile (Henriksson et al. 2019; Jääskeläinen et al. 2018, 2021; Laketic et al. 2022; Lehikoinen et al. 2018; Mao et al. 2021; Mitro et al. 2020).

Maternal metabolism undergoes adjustments due to increased nutrient demands of the pregnancy, leading to elevated levels of several hormones, glucose, and lipids (Haddad-Tóvolli and Claret 2023; Herrera 2002; Montelongo et al. 1992). The maternal metabolic environment is significantly directed to fetal development, relying on the metabolite transfer from maternal circulation across the placenta (Desai and Hales 1997; Hadden and McLaughlin 2009). Therefore, disruptions in maternal metabolism may have long-term consequences.

The pathogenesis of depression is involved in the dysregulation of metabolic pathways, such as neurotransmission, nitrogen, methylation, and lipid metabolism (Bot et al. 2020; Pu et al. 2020). In depressive patients, the usage of SSRIs modulates acylcarnitine, lipids, and amino acids profiles (Bhattacharyya et al. 2019; Caspani et al. 2021; MahmoudianDehkordi et al. 2021; Pan et al. 2018). Additionally, the SSRI usage correlates with increased plasma levels of triacylglycerides in early treatment weeks and hypercholesterolemia in later use (Chávez-Castillo et al. 2018).

However, studies on the impact of SSRI medication on circulating metabolome during pregnancy are lacking. To address this, we employed a targeted nuclear magnetic resonance (NMR) spectroscopy-based approach. This approach enables comprehensive profiling of metabolites and lipoproteins (Würtz et al. 2017), which can elucidate the molecular effects of SSRI usage in pregnant women, ensuring safe depression treatment during pregnancy and promoting normal fetal development.

## 2. Materials & methods

### 2.1. Study cohort and blood samples

This study utilized samples and data from the Kuopio Birth Cohort (KuBiCo) (www.kubico.fi) (Huuskonen et al. 2018). All participants in the KuBiCo study provided a written informed consent. The study was conducted in accordance with the guidelines outlined in the Declaration of Helsinki. Ethical approval was obtained from the Research Ethics Committee of the Hospital District of Central Finland in Jyväskylä, Finland on November 15, 2011 (18U/2011).

Maternal venous blood samples were collected during the first trimester (between gestational weeks 9 to 11) and during the delivery within the years 2013-2020 from women giving birth at Kuopio University Hospital (KUH). Trained laboratory staff or midwives collected the venous blood samples, and the separated serum was stored at −80 °C until analysis. A total of 409 maternal serum samples were collected from 239 pregnancies. Of these, 179 samples were from women using SSRIs (122 pregnancies), and 230 samples from non-depressive controls without any antidepressant medication (117 pregnancies). Samples were available at both timepoints for 57 SSRI users and 113 controls. However, some individuals had only samples taken at one timepoint; in SSRI users 18 women had samples at the first trimester and 47 samples at delivery and in control group two women had samples at the first trimester and other two at delivery.

Data on SSRI usage and the diagnosis of gestational diabetes (GDM) (ICD-10 code O24.4) and preeclampsia (ICD-10 code O14.9) were collected from the KUH Birth Register, which contains demographic and clinical information about all mothers participating in the study. The Birth Register also included a self-reporting section where the use of over-the-counter and prescription drugs, as well as smoking during pregnancy, could be reported. The smoking status of women was confirmed by measuring serum cotinine level from delivery blood samples using a previously described method (Sahlman et al. 2022).

### 2.2. Nuclear magnetic resonance spectroscopy

The maternal serum samples from the first trimester and delivery were subjected to a widely used high throughput targeted NMR metabolomics platform (Nightingale Health Ltd., Helsinki, Finland)(Julkunen et al. 2023; Würtz et al. 2017). In several studies SSRIs are linked with alterations in the lipid and amino acid profiles (Caspani et al. 2021; Fjukstad et al. 2016; MahmoudianDehkordi et al. 2021; Pan et al. 2018). Consequently, this NMR metabolomics platform includes measurement of selected small molecule metabolites, such as amino acids and fatty acids, as well as larger molecules like lipoproteins, and selected ratios between these compounds. A total of 250 variables were analyzed. The protocol has been described previously in detail (Soininen et al. 2015; Würtz et al. 2017).

### 2.3. Statistical analyses

Descriptive statistics included means, standard deviations, frequencies, and percentages. Regarding the background characteristics, continuous variables were analyzed with independent samples t-test or Wilcoxon rank-sum test and categorical variables with Pearson Chi-square test. Data visualization employed GraphPad Prism version 9.2.0 for Windows (GraphPad Software, San Diego, California USA, www.graphpad.com). In our analysis, we accounted for the duration of SSRI treatment by including individuals who were actively using SSRIs or had discontinued the usage within the same trimester when the measurement was done.

Metabolic measures at two timepoints (the first trimester and delivery) and the changes occurring in the metabolic profile between the first trimester and delivery (delta, Δ) in both SSRI users and controls were compared using Wilcoxon rank-sum test and Rank-Biserial correlations. Multivariate analysis employed principal component analysis (PCA). Due to the correlated nature of the measured variables, we used the number of latent components needed to explain 95% of the variance in the data to adjust the α-level for multiple testing (Bonferroni method). Sixteen latent components were required to explain 95% of the variance, resulting in an adjusted α-level of 0.0031. Differences with p-values between 0.05 and 0.0031 were considered as trends. Statistical analyses were performed using JASP (Version 0.16.3).

Linear regression and linear mixed model analyses were conducted on scaled and log1p-normalised metabolites or metabolite ratios to further inspect variables with significant differences between groups. The linear regression model was adjusted for BMI and GDM to account for their known influences on the circulating metabolome (Mokkala et al. 2020; Wahab et al. 2022). Study group (SSRI users vs. controls) and additional medication use were included as cofactors. Additionally, cotinine level was included as a predictor for delivery timepoint. In the linear mixed model analysis, timepoint and study group were included as fixed effect factors, and KuBiCo-ID as a random effect grouping factor. Finally, Spearman’s correlation coefficients were used for non-parametric continuous variables.

## 3. Results

### 3.1. Descriptive characteristics

Descriptive characteristics of the participants are represented in Table 1. The SSRI users had higher BMI, serum cotinine levels, placental weights, prevalence of GDM, and they used additional medication more often compared to the controls. Moreover, the Apgar scores at 1 minute and at 5 minutes were significantly lower for newborns born to SSRI users compared to the controls.

**Table 1.** Descriptive characteristics of study women with selective serotonin reuptake inhibitor users (SSRI) and controls.

| Variable | Control<br>N = 117 | SSRI<br>N = 122 | p-value* |
| --- | --- | --- | --- |
| Age, years | 30.4±5.0 | 30.6 ± 5.3 | 0.837 |
| Parity | 0.69 ± 0.9 | 0.91± 1.3 | 0.128 |
| Self-reported smoking <sup>1</sup> |  |  |  |
| Not smoking | 85 (72.7) | 75 (61.5) |  |
| Quit smoking before pregnancy | 7 (6.0) | 7 (5.7) | 0.822<sup>a</sup> |
| Smoked during pregnancy | 24 (20.5) | 35 (28.7) | 0.104<sup>a</sup> |
| Use of additional medication | 44 (37.6) | 74 (60.7) | < **0.001** |
| BMI at the beginning of pregnancy, kg/m<sup>2</sup> <sup>1</sup> | 24.7 ± 5.1 | 27.6± 7.2 | < **0.001** |
| <25 | 81 (69.2) | 55 (45.1) |  |
| 25-30 | 21 (18.0) | 30 (24.6) | **0.026<sup>b</sup>** |
| >30 | 15 (12.8) | 37 (30.3) | < **0.001<sup>b</sup>** |
| BMI before delivery, kg/m<sup>2</sup> <sup>2</sup> | 29.8± 5.1 | 32.1± 6.6 | **0.003** |
| <25 | 15 (12.8) | 12 (9.8) |  |
| 25-30 | 56 (47.9) | 41 (33.6) | 0.839<sup>b</sup> |
| > 30 | 44 (37.6) | 69 (56.6) | 0.120<sup>b</sup> |
| Change in BMI during pregnancy, kg/m<sup>2</sup> <sup>3</sup> | 4.9 ± 1.9 | 4.8± 3.3 | 0.782 |
| Number of fetuses | 1.0 (0.0) | 1.0 (0.1) |  |
| GDM <sup>1</sup> | 22 (19.1) | 37 (30.3) | **0.046** |
| Pre-eclampsia <sup>1</sup> | 4 (3.4) | 5 (4.0) | 0.841 |
| Cotinine, ng/ml <sup>1</sup> | 2.8± 20.4 | 4.7 ± 16.2 | **0.002<sup>e</sup>** |
| **Obstetric and early neonatal characteristics** |  |  |  |
| Placental weight (g) <sup>1</sup> | 581.8 ± 134.9 | 618.0 ± 129.2 | **0.014<sup>e</sup>** |
| Gestational weeks at delivery |  |  |  |
| Term ≥ 37 | 115 (98.3) | 123 (97.6) |  |
| Moderately preterm 32-36 | 2 (1.7) | 2 (1.6) | 0.973<sup>c</sup> |
| Very preterm ≤ 31 | 0 | 1 (0.79) | <sup>d</sup> |
| Birth weight, grams <sup>1</sup> | 3517.8± 476.9 | 3460.3± 471.8 | 0.352 |
| Male sex <sup>1</sup> |  |  | 0.268 |
| male | 53 (45.3) | 63 (51.6) |  |
| Apgar scores at 1min <sup>1</sup> | 8.7± 0.8 | 8.3± 1.4 | **0.005** |
| Apgar scores at 5min <sup>1</sup> | 9.1 ± 0.6 | 8.7± 1.1 | **0.003** |

Data is shown as mean ± SD or N (%); \* Independent samples *t*-test or Wilcoxon rank-sum test for continuous variables, Pearson Chi-square test for categorical variables; <sup>1</sup> n = 230; <sup>2</sup> n = 237; <sup>3</sup> n = 229; Logistic regression <sup>a</sup>
the values were correlated to "not smoking", <sup>b</sup> the values were correlated to "BMI < 25", <sup>c</sup> the values were correlated to "term > 37"; <sup>d</sup> n = 0 in control group; <sup>e</sup> Wilcoxon rank-sum test; p < 0.05 bolded

Among the SSRI users, the most common indication for SSRI medication was depression (64.8%), although SSRIs were also prescribed for other mental disorders (Table S1). Majority of women (90.2%) using SSRIs had already initiated medication before pregnancy, and 61.5% (n = 75) continued medication throughout the entire pregnancy. Discontinuation of the medication was most common during the last trimester (14.8%, n = 18), while 8.2% (n = 10) of women ceased taking the medication during the first trimester and 5.7% (n = 7) during the second trimester. Moreover, 9.8% (n = 12) of women initiated SSRI medication during pregnancy. Citalopram was identified as the most frequently used SSRI medication, accounting for 36.8% (n = 45) of the cases, followed by escitalopram (27.9%, n = 34), sertraline (25.4%, n = 31), and other SSRIs (9.8%, n = 12) (Table S1).

### 3.2. SSRI usage is associated with altered lipoprotein composition

In the targeted NMR analysis, 250 variables were analyzed (Table S2). In the PCA, SSRI users and controls showed slight separation based on the first two principal components (Figure S1a). Furthermore, the metabolic profile patterns differed between the first trimester and delivery, as expected (Figure S1b). Figure 1 represents the significance and magnitude of the observed changes between study groups at both time points.

**Fig 1.**
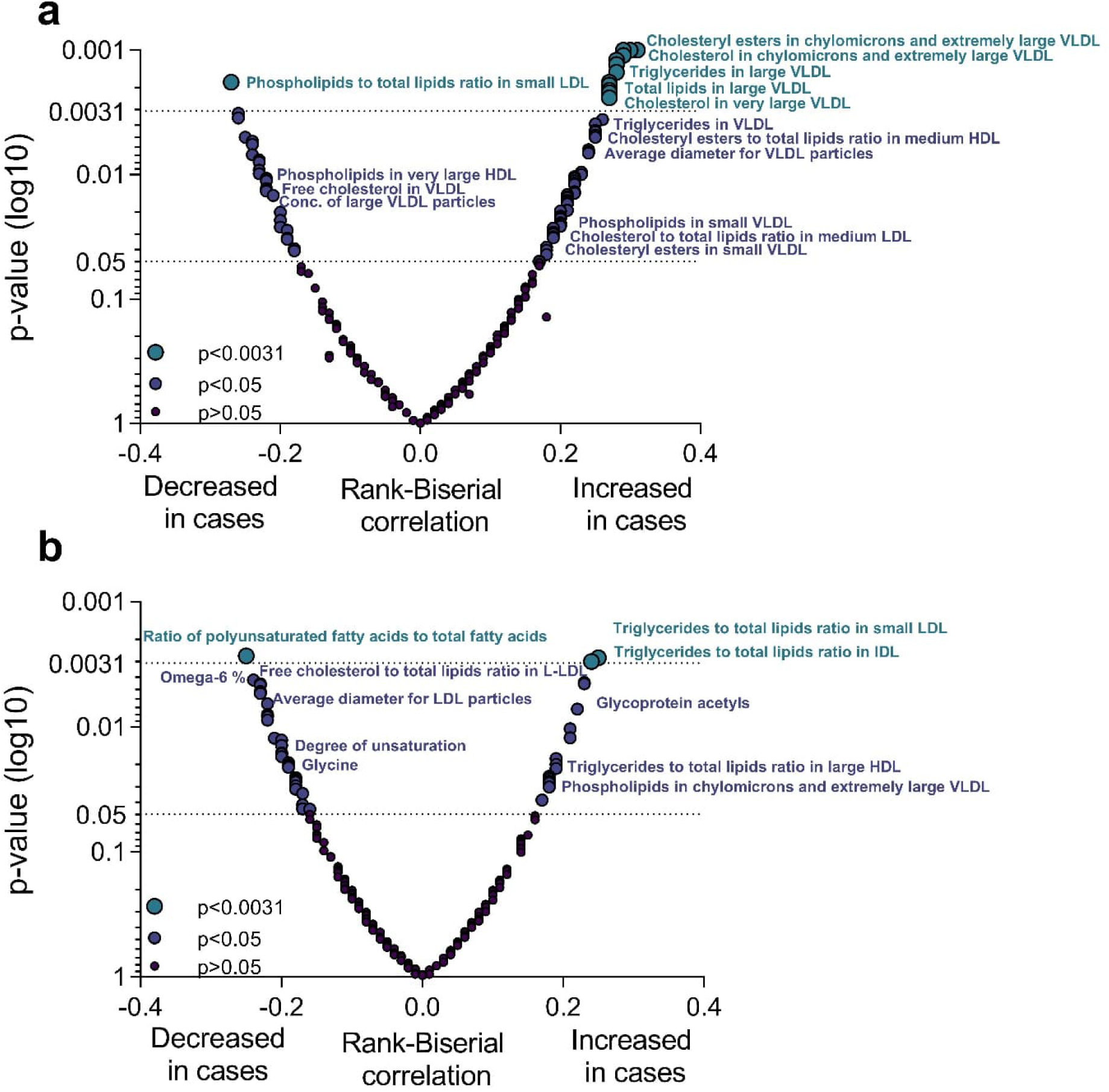
Volcano plot summarizing the effect of selective serotonin reuptake inhibitor (SSRI) use on the metabolic variables. The significance and magnitude of the observed changes in the first trimester (panel a) and at delivery (panel b) between SSRI users and control women. Wilcoxon rank-sum test was used to compare the metabolite levels between groups (n = 239). The effect size of SSRI use compared to control group is represented as rank-biserial correlation. Omega-6%, ratio of omega-6 fatty acids to total fatty acids

When comparing the first trimester samples between SSRI users and non-users, there were significant differences in 96 detected variables (p-value < 0.05 (16 expected by chance)), of which 19 variables had p-values below the multiple testing adjusted α-level < 0.0031 (Figure S2 and S3). 17 out of these 19 are presented in Figure 2a-b. Due to extremely low concentrations, concentration of chylomicrons and very large and extremely large very low-density lipoprotein (VLDL) particles were excluded from Figure 2. The significantly altered metabolites were all related to the composition of chylomicrons and VLDL particles: the level of triglycerides, phospholipids, cholesteryl esters, total lipids, and free cholesterol in these lipoprotein particles were increased in SSRI users (Figure 2a). Moreover, the circulating concentrations of chylomicrons, as well as very large and extremely large VLDL particles were elevated (excluded from Figure 2). Significant changes also occurred in the lipoprotein lipid ratios, where cholesteryl esters to total lipids ratio in small and medium low-density lipoprotein (LDL) was increased and phospholipids to total lipids ratio in small LDL was decreased in SSRI users (Figure 2b). Furthermore, the SSRI users exhibited trends of increased acetoacetate, glucose, monounsaturated fatty acids, ratio of apolipoprotein B to apolipoprotein A1, ratio of monounsaturated fatty acids to total fatty acids, and VLDL-, triglyceride- and LDL-related variables (Figures S2 and S3). In contrast, decreasing trends were observed in high-density lipoprotein (HDL)-related variables, the ratio of polyunsaturated fatty acids to total fatty acids, the ratio of polyunsaturated fatty acids to monounsaturated fatty acids, the ratio of omega-6 fatty acids to total fatty acids, and the ratio of linoleic acid to total fatty acids (Figures S2 and S3).

**Fig 2.**
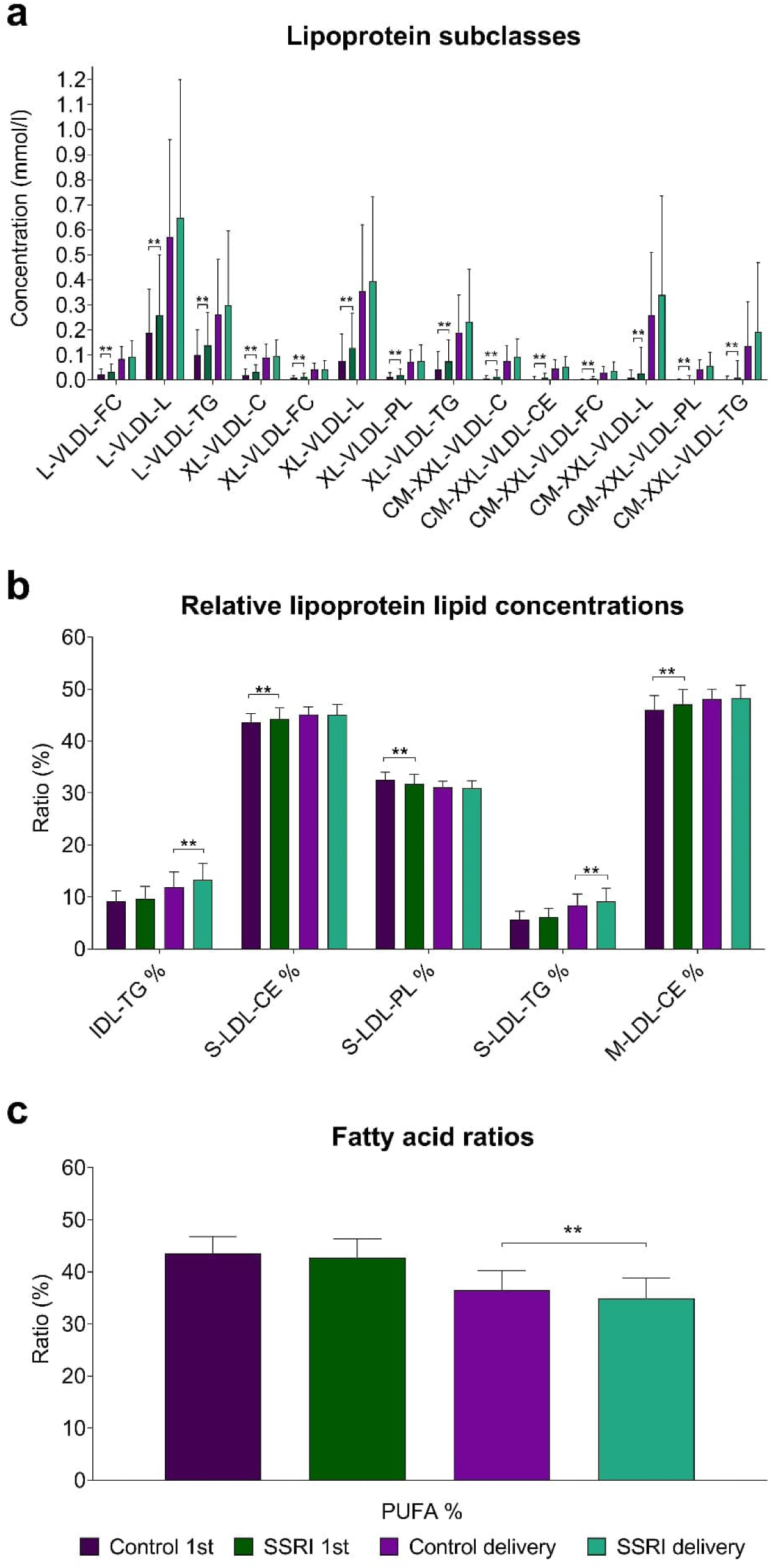
Comparison of the lipoprotein and metabolite levels from the first trimester and delivery timepoints of pregnancy between selective serotonin reuptake inhibitor (SSRI) users and control women. Wilcoxon rank-sum test was used to compare the metabolite levels between groups (n = 239). ** Significant differences at the multiple testing corrected α-level are represented (p < 0.0031). Median values with interquartile ranges (error bars) are shown. Panel a represents lipoprotein subclasses as concentrations (mmol/l), panel b represents relative lipoprotein lipid concentrations as ratios (%) and panel c represents fatty acid ratios (%). L-VLDL-FC, free cholesterol in large very low-density lipoprotein; L-VLDL-L total lipids in large very low-density lipoprotein; L-VLDL-TG, triglycerides in large very low-density lipoprotein; XL-VLDL-C, cholesterol in very large very low-density lipoprotein; XL-VLDL-FC, free cholesterol in very large very low-density lipoprotein; XL-VLDL-L total lipids in very large very low-density lipoprotein; XL-VLDL-PL, phospholipids in very large very low-density lipoprotein; XL-VLDL-TG, triglycerides in very large very low-density lipoprotein; CM-XXL-VLDL-C, cholesterol in chylomicrons and extremely large very low-density lipoprotein; CM-XXL-VLDL-CE, cholesteryl esters in chylomicrons and extremely large very low-density lipoprotein; CM-XXL-VLDL-FC, free cholesterol in chylomicrons and extremely large very low-density lipoprotein; CM-XXL-VLDL-L, total lipids in chylomicrons and extremely large very low-density lipoprotein; CM-XXL-VLDL-PL, phospholipids in chylomicrons and extremely large very low-density lipoprotein; CM-XXL-VLDL-TG, triglycerides in chylomicrons and extremely large very low-density lipoprotein; IDL-TG %, triglycerides to total lipid ratios in intermediate-density lipoprotein; S-LDL-CE %, cholesteryl esters to total lipids ration in small low-density lipoprotein; S-LDL-PL %, phospholipids to total lipids ration in small low-density lipoprotein; S-LDL-TG %, triglycerides to total lipids ration in small low-density lipoprotein; M-LDL-CE %, cholesteryl esters to total lipids ration in medium low-density lipoprotein; PUFA%, ratio of polyunsaturated fatty acids to total fatty acids

When comparing the maternal samples obtained at delivery, 50 variables exhibited a p-value < 0.05, and three remained significant after Bonferroni correction (Figures S2 and S3). Of the significantly altered variables, SSRI users had an increased ratio of triglycerides to total lipids in small LDL (S-LDL-TG%) (Mdn = 9.1 % vs. 8.4, p < 0.003, r = 0.25) and triglycerides to total lipids in intermediate-density lipoprotein (IDL-TG%) (Mdn = 13.3% vs. 12%, p < 0.003, r = 0.24) and a decreased ratio of polyunsaturated fatty acids to total fatty acids (PUFA%) (Mdn = 34.8% vs. 36.5%, p < 0.003, r = −0.25) (Figure 2b-c). Considering the trends, most of the altered variables were associated with a decreased rate of the relative lipoprotein lipid concentrations with a few exceptions of elevated lipid contents in lipoprotein subclass groups (Figure S3). Furthermore, SSRI users exhibited an increase in glycoprotein acetyls, the ratio of triglycerides to phosphoglycerides, and the ratio of saturated fatty acids to total fatty acids. They also showed a trend of decreased glycine, degree of unsaturation of fatty acids, the ratio of omega-6 fatty acids and linoleic acids to total fatty acids, and the ratio of polyunsaturated fatty acids to monounsaturated fatty acids (Figure S2).

When confounding variables (BMI, GDM, cotinine level, additional medication use) were taken into account, the SSRI usage remained as a significant distinguishing factor for all the metabolic variables at both timepoints, except for the lipoprotein lipid ratios (triglycerides in chylomicrons and extremely large VLDL, phospholipids to total lipids ratio in small LDL, cholesteryl esters to total lipids ratio in medium LDL and, cholesteryl esters to total lipids ratio in small LDL) in the first trimester (Tables S3 and S4). Furthermore, BMI correlated with all the significantly altered variables in the first trimester.

While comparing the effect of SSRI usage on pregnancy-driven metabolic changes, 24 variables exhibited a p-value < 0.05 (Figure S4). Cholesterol to total lipids ratio in medium LDL (M-LDL-C%) (Δ = −2.1% vs. −0.84 %, p < 0.001, r = −0.34) (Figure 3a) and cholesterol to total lipids ratio in large LDL (L-LDL-C%) (Δ = −2.93% vs. −1.78 %, p < 0.003, r = −0.32) remained significant after correction for multiple testing (Figure 3b). Interestingly, the direction of the effect was similar in the control group, but more pronounced in SSRI users. Levels of several cholesterol variables (total cholesterol, VLDL, LDL, HDL) trended towards decrease in SSRI users as well as several variables in lipoprotein subclass group (Figure S4). On the contrary, the ratio of triglycerides to total lipids in IDL and medium and large LDL trended towards increase in SSRI users (Figure S4). In the linear mixed model analysis, an interaction of pregnancy progression and SSRI usage was observed for M-LDL-C % (F(1, 149.62) = 7.60, p = 0.007) and for L-LDL-C% (F(1, 149.86) = 10.11, p = 0.002).

**Fig 3.**
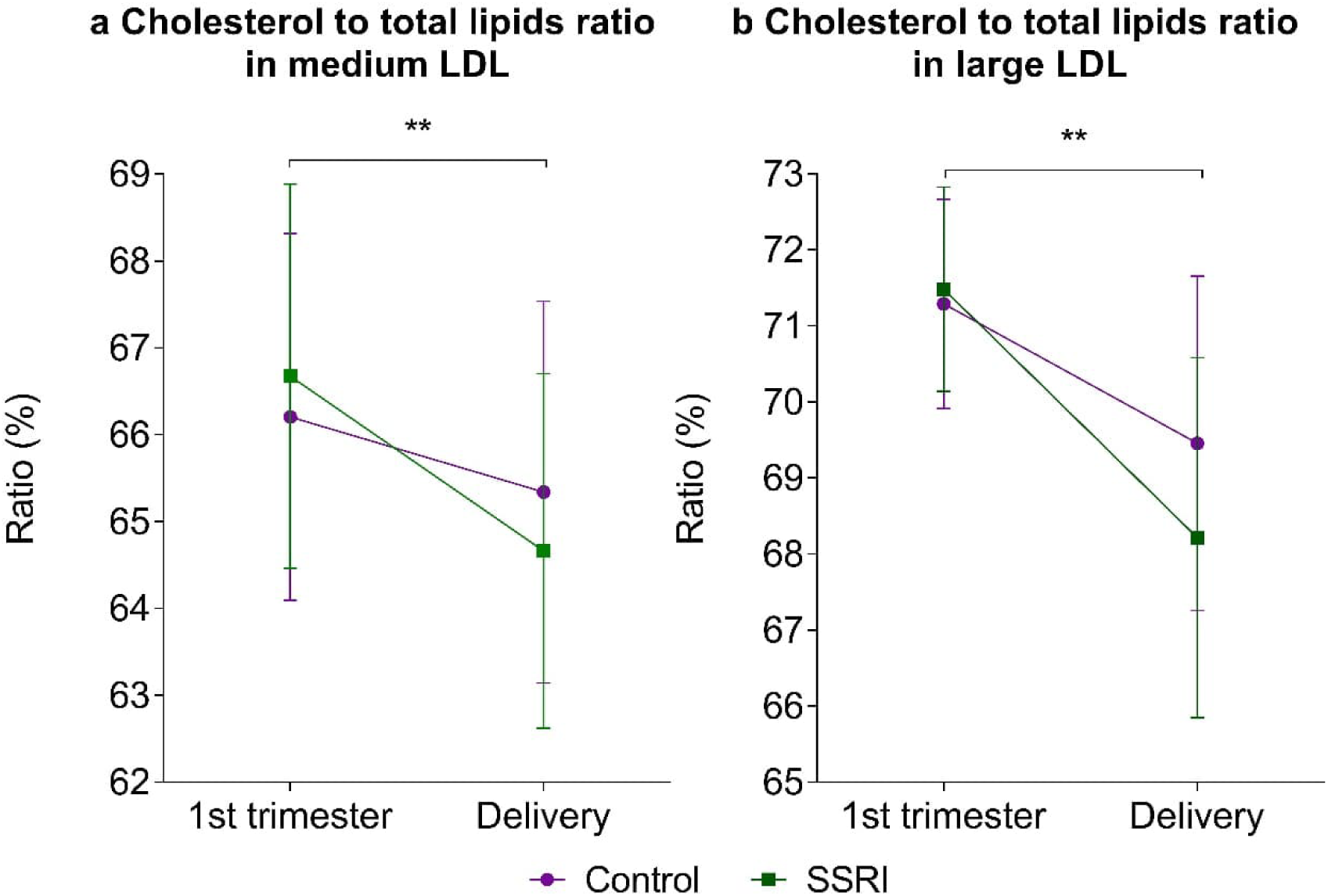
Comparison of the changes during pregnancy between SSRI users and control women. Wilcoxon rank-sum test was used to compare the variables between groups (n = 150). ** Significant differences at the multiple testing corrected α-level are represented (p < 0.0031). Error bars represent the median value and interquartile range. Panel a represents cholesterol to total lipids ratio in medium low-density lipoprotein (M-LDL-C%) and panel b represents cholesterol to total lipids ratio in large LDL (L-LDL-C%)

Furthermore, among the SSRI users, the Apgar score at 5 minutes demonstrated negative correlations with several VLDL subtypes and PUFA%, which was not observed in the controls (Figure 4). Additionally, maternal BMI in the first trimester (r = 0.24, p = 0.009) and at delivery (r = 0.30, p = 0.001) correlated with placental weight, a pattern not seen in the controls (Figure S5a-b). Finally, as a post-hoc analysis, we investigated the metabolic profile differences based on SSRI indication (depression vs. other mental disorder), revealing minimal variations – five variables differed at delivery, of which none remained significant, according to the multiple testing adjusted α-level < 0.0031 (Figure S6).

**Fig 4.**
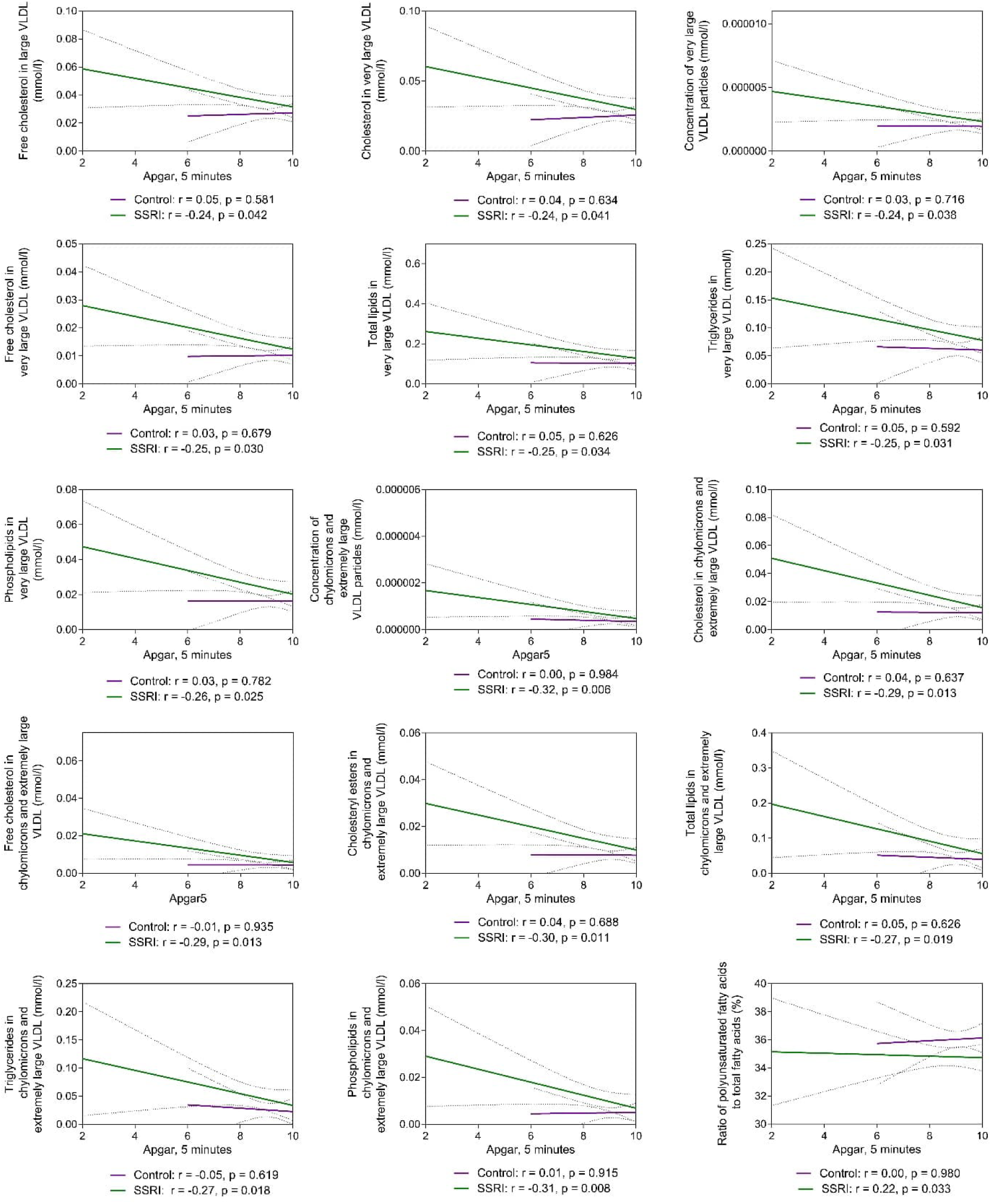
Correlation between metabolic variables and Apgar scores at 5 minutes in selective serotonin reuptake inhibitor (SSRI) users (n = 74) and control women (n = 115). Spearman’s correlations, with a 95 % confidence interval at a significance level of p < 0.05, are depicted. LDL, low-density lipoprotein; VLDL, very low-density lipoprotein

## 4. Discussion

This study indicates that SSRI usage during pregnancy is associated with relatively minor changes in the circulating metabolome, primarily affecting lipid metabolism. To our knowledge, this paper is the first to study the effect of SSRI treatment on circulating maternal metabolome with targeted NMR metabolite profiling. Prior studies have documented the impact of SSRIs in cohorts of non-pregnant individuals (Bhattacharyya et al. 2019; Caspani et al. 2021; MahmoudianDehkordi et al. 2021; Pan et al. 2018) alongside gravid murine models (Ramsteijn et al. 2020).

In the first trimester, the cohorts showed differences in VLDL and chylomicron particle composition. At the time of delivery, significant differences between the two groups were found in the ratio of triglycerides to total lipids in LDL and IDL (S-LDL-TG%, IDL-TG%) and fatty acid ratios (PUFA%). Furthermore, the interaction of pregnancy progression and SSRI usage prompted a descending effect in the ratio of cholesterol to total lipids in medium and large LDL particles (M-LDL-C% and L-LDL-C%).

### 4.1. SSRI-induced metabolic alterations

The impact of SSRIs on metabolic disturbances remains unclear. MahmoudianDehkori et al. (2021) proposed that changes in the metabolic profile induced by SSRIs might reflect mitochondrial energy processes involved in depression recovery (MahmoudianDehkordi et al. 2021). Additionally, long-term SSRI usage might trigger proinflammatory responses, tied to pathophysiology of conditions such as metabolic syndrome and type 2 diabetes (Chávez-Castillo et al. 2018). In our cohort, the level of glycoprotein acetyls, marker of low-grade inflammation associated with antenatal depression and GDM in overweight and obese women (Lahti-Pulkkinen et al. 2020), showed an increasing trend at delivery in SSRI users. On the other hand, the depression-related pathways, such as the persistent activation of the hypothalamic-pituitary-adrenal (HPA) axis and chronic inflammation, might disrupt lipid levels (Otte et al. 2016).

In our study, SSRI usage was associated with an increased level of lipid contents in VLDL and other lipoprotein subtypes, and similar alterations have been observed in non-pregnant depressive individuals without SSRIs (Bot et al. 2020). Zheng et al. (2012) identified lipoproteins (LDL, VLDL) as the most significant factor distinguishing depressed patients from healthy controls (Zheng et al. 2012). On the other hand, in murine models, fluoxetine treatment correlates with increased expression of lipogenic enzymes and decreased the expression of lipolytic enzymes (Pan et al. 2018). In depressive patients, SSRI treatment has been associated e.g., with increased serum triglycerides, total cholesterol, LDL, and phosphatidylcholines levels (Caspani et al. 2021; Fjukstad et al. 2016; MahmoudianDehkordi et al. 2021; Pan et al. 2018). Unfortunately, the composition of lipoprotein subtypes was not measured in these previous studies. We observed merely an increasing trend in VLDL cholesterol and total triglycerides levels, while no differences were noted in total cholesterol, LDL, or phosphatidylcholines levels.

Furthermore, SSRI treatment for depression seems to induce alterations in the acylcarnitine, amino acid (e.g., arginine) and biogenic amine profiles (MahmoudianDehkordi et al. 2021). In our study, we observed a decreasing trend in glucose levels and the amino acid glycine in SSRI users during the first trimester and at delivery, respectively. Similarly, altered level of metabolites involved in amino acid (e.g., tryptophan) and energy metabolism (e.g., glucose, citrate, 3-hydroxybutyrate, and creatinine) have been observed in depressive patients (Duan and Xie 2020) and in pregnant individuals with antenatal depression (Henriksson et al. 2019; Laketic et al. 2022; Mao et al. 2021; Mitro et al. 2020). Altogether the studies provide evidence that SSRI usage has the potential to cause metabolic alterations. However, the possibility that the observed changes are related to depression cannot be excluded. Nevertheless, our post-hoc analysis comparing the metabolomes of individuals with depression and other mental health disorders revealed only minor differences.

It seems that the observed metabolic alterations are more pronounced during the first trimester but even out as pregnancy progresses and normal pregnancy-related metabolic changes occur. Similar normalization of metabolic disturbances induced by paroxetine has been observed in gravid mice (Zha et al. 2019).

### 4.2. Clinical impact

Despite being statistically significant, the observed metabolic changes are relatively minor compared to the typical physiological changes in pregnancy. Nevertheless, lipoprotein lipid physiology crucially impacts the mother, the developing fetus, and their future health. Maternal dyslipidemias are linked with adverse perinatal outcomes, as reviewed elsewhere (Herrera 2002; Herrera and Ortega-Senovilla 2017). Therefore, the metabolic safety of SSRI usage during pregnancy must be established. Recently, a possible association between increased levels of specific lipid contents in VLDL subtypes in the first trimester of pregnancy and the risk of fetal congenital heart defects, particularly in obese mothers was noted (Huida et al. 2023). Since the use of SSRIs during pregnancy may pose a risk for the development of fetal congenital heart disease (Nie et al. 2022), the possible link must be elaborated. Within our cohort, small negative correlations were found between the Apgar scores at 5 minutes, indicator of newborn well-being, and level of several VLDL subtypes among SSRI users. Moreover, the Apgar scores 1 and 5 minutes after birth were significantly lower in SSRI users. Therefore, further studies are necessary to assess the potential associations between altered the composition of VLDL subtypes and the risk of postnatal metabolic complications as well as to account for the observed individual variation in the metabolite levels.

### 4.3. Strengths and Limitations

This study benefits from a large sample size, including over 400 serum samples from pregnant women using SSRIs and control women. It covers samples from the first trimester and delivery, representing different pregnancy phases and enabling the assessment of metabolic changes during pregnancy. Confounding factors such as BMI and GDM were considered, and a standardized NMR platform with a targeted approach was employed, offering reproducibility and comprehensive metabolic profiling especially for lipoprotein composition. Compared to more traditional clinical assays, NMR spectroscopy allows more extensive profiling of lipids and other metabolites with one analysis. However, compared to non-targeted mass spectrometry metabolomics approaches, the targeted NMR approach does lack sensitivity for low-concentration metabolites and may overlook relevant differentiating metabolites between SSRI users and controls.

Limitations include the inability to differentiate the effects of SSRIs from depression and to measure treatment response of SSRIs, the lack of confirmation of SSRI usage through serum analysis, reliance on medical records for SSRI usage information, and the lack of knowledge on accurate blood sampling times. Additionally, unexplored contributors to metabolome include maternal diet, genetics, and microbiome. Limited matching samples in the first trimester and delivery for the SSRI group hindered comparisons of the magnitude of SSRIs’ effects on metabolic measures between the timepoints. Due to limited statistical power, the study is unable to exclude the impact of individual SSRIs on the metabolic variables despite the evidence of varying effects on metabolic parameters (Beyazyüz et al. 2013).

## 5. Conclusion

In conclusion, SSRI usage was associated with minor alterations in the circulating lipoprotein profile of pregnant women. Given that the observed changes are relatively slight compared to those caused by pregnancy itself, their clinical impact is likely limited. Nevertheless, there is a need for further research into the maternal metabolic alterations prompted by SSRIs, as the altered lipoprotein composition could pose risks to fetal development. The findings contribute to a better understanding of safe use of SSRI medication during pregnancy which is of critical importance due to the risks linked with untreated depression for the mother and developing fetus.

## Supporting information

Supplementary Informartion

## Declarations

### Data Availability

The data are not publicly available due to their containing information that could compromise the privacy of research participants. The study plan approved by the ethical committee and the participant consent terms preclude public sharing of these sensitive data, even in anonymized form.

### Authors Contributions

A.I., O.K., H.S. and J.R. conception and design of the study. H.S. L.K-N, acquisition of data. A.I. and O.K. analysis and interpretation of data. A.I. drafting the article. All authors revised the draft and approved the final version for the submission.

### Ethical Statements

Ethics approval was obtained from the Research Ethics Committee of the Hospital District of Central Finland in Jyväskylä, Finland on November 15, 2011 (18U/2011).

### Consent to participate

Informed consent was obtained from all individual participants included in the study.

## Acknowledgements

This paper belongs to the studies carried out by the Kuopio Birth Cohort consortium (www.KuBiCo.fi) and we thank our colleagues who are responsible for the design and conduct of the KuBiCo. We thank Ms. Pirjo Hänninen, Ms. Sonja Holopainen and MSc. Essi Järvelä for expert laboratory assistance, and the staff of the Department of Obstetrics and Gynecology in Kuopio University Hospital.

