## Supplementary Informartion for "Longitudinal Metabolic Profiling of Women Using Selective Serotonin Reuptake Inhibitors During Pregnancy"

**Table S1.** Information on SSRI usage and indications for SSRIs.

| SSRI use | | |
|---|---|---|
| **SSRI** | **Dose (mg)** | **n (%)** |
| Citalopram | 5–40 | 45 (36.8) |
| Escitalopram | 2.5–20 | 34 (27.9) |
| Sertraline | 25–200 | 31 (25.4) |
| Fluoxetine | 5–60 | 6 (4.9) |
| Paroxetine | 5–40 | 3 (2.5) |
| Fluvoxamine | 150 | 1 (0.8) |
| Escitalopram, Fluoxetine | 20, 40 | 1 (0.8) |
| Escitalopram, Sertraline | 20, 100 | 1 (0.8) |
| **SSRI usage during pregnancy** | | **n (%)** |
| Permanent | | 75 (61.5) |
| Discontinued during 1st trimester | | 10 (8.2) |
| Discontinued during 2nd trimester | | 7 (5.7) |
| Discontinued during 3rd trimester | | 18 (14.8) |
| Initiated during 1st trimester | | 2 (1.6) |
| Initiated during 2nd trimester | | 6 (4.9) |
| Initiated during 3rd trimester | | 4 (3.3) |
| **Indications for SSRIs**[1] | | |
| **Indication** | | **n (%)** |
| Depression | | 79 (64.8) |
| Panic disorder | | 23 (18.9) |
| Anxiety | | 12 (9.8) |
| Other (bipolar disorder, obsessive-compulsive disorder, depersonalization-derealization disorder, social anxiety disorder) | | 5 (4.1) |

[1] n=119; SSRI = Selective serotonin reuptake inhibitor

**Table S2.** Wilcoxon rank-sum test of the difference between women using selective serotonin reuptake inhibitors (SSRIs) during pregnancy and control women. The difference in the variables has been analyzed at two timepoints (1st trimester and at delivery). Moreover, the effect of SSRIs on the pregnancy-driven change in the metabolome (delta) has been compared by Wilcoxon rank-sum test.

| | | | | 1ST TRIMESTER (N=185) | | | | | | | | | | DELIVERY (N=201) | | | | | | | | | | CHANGE BETWEEN DELIVERY AND 1ST TRIMESTER (DELTA) (N=150) | | | | | | | | |
|---|---|---|---|---|---|---|---|---|---|---|---|---|---|---|---|---|---|---|---|---|---|---|---|---|---|---|---|---|---|---|---|---|
| | | | | CONTROL (N=115) | | | | SSRI (N=70) | | | | | | CONTROL (N=112) | | | | SSRI (N=89) | | | | | | CONTROL (N=110) | | | | SSRI (N=40) | | | | |
| GROUP | VARIABLE | ABBRE-VIATION | UNIT | MEDIAN | IQR | MEAN | SD | MEDIAN | IQR | MEAN | SD | r | p | MEDIAN | IQR | MEAN | SD | MEDIAN | IQR | MEAN | SD | r | p | MEDIAN | IQR | MEAN | SD | MEDIAN | IQR | MEAN | SD | r |
| AMINO ACIDS | Alanine | Ala | mmol/l | 0.38 | 0.07 | 0.37 | 0.06 | 0.39 | 0.09 | 0.38 | 0.07 | 0.06 | 0.4978 | 0.45 | 0.13 | 0.47 | 0.11 | 0.46 | 0.10 | 0.47 | 0.08 | 0.04 | 0.6401 | 0.09 | 0.12 | 0.09 | 0.11 | 0.08 | 0.13 | 0.08 | 0.10 | -0.08 |
| AMINO ACIDS | Glutamine | Gln | mmol/l | 0.54 | 0.08 | 0.54 | 0.06 | 0.52 | 0.09 | 0.53 | 0.07 | -0.14 | 0.1239 | 0.48 | 0.11 | 0.50 | 0.10 | 0.48 | 0.10 | 0.48 | 0.08 | -0.06 | 0.4455 | -0.04 | 0.12 | -0.04 | 0.09 | -0.06 | 0.10 | -0.06 | 0.09 | -0.09 |
| AMINO ACIDS | Glycine | Gly | mmol/l | 0.22 | 0.05 | 0.21 | 0.04 | 0.21 | 0.06 | 0.20 | 0.05 | -0.09 | 0.2955 | 0.22 | 0.07 | 0.22 | 0.05 | 0.20 | 0.07 | 0.21 | 0.05 | -0.19 | **0.0192** | 0.00 | 0.06 | 0.01 | 0.05 | 0.01 | 0.05 | 0.01 | 0.05 | 0.00 |
| AMINO ACIDS | Histidine | His | mmol/l | 0.10 | 0.01 | 0.10 | 0.01 | 0.10 | 0.02 | 0.10 | 0.01 | -0.06 | 0.4608 | 0.09 | 0.02 | 0.09 | 0.02 | 0.09 | 0.02 | 0.09 | 0.02 | -0.13 | 0.1085 | -0.01 | 0.03 | -0.01 | 0.02 | -0.01 | 0.03 | -0.01 | 0.02 | -0.04 |
| AMINO ACIDS (AROMATIC) | Phenylalanine | Phe | mmol/l | 0.08 | 0.02 | 0.08 | 0.01 | 0.08 | 0.02 | 0.07 | 0.02 | -0.09 | 0.3061 | 0.09 | 0.03 | 0.09 | 0.02 | 0.09 | 0.02 | 0.09 | 0.01 | -0.01 | 0.8980 | 0.01 | 0.02 | 0.01 | 0.02 | 0.01 | 0.03 | 0.01 | 0.02 | 0.05 |
| AMINO ACIDS (AROMATIC) | Tyrosine | Tyr | mmol/l | 0.06 | 0.02 | 0.06 | 0.02 | 0.06 | 0.02 | 0.06 | 0.02 | -0.08 | 0.3451 | 0.05 | 0.02 | 0.05 | 0.01 | 0.05 | 0.01 | 0.05 | 0.01 | 0.00 | 0.9854 | -0.01 | 0.03 | -0.01 | 0.02 | -0.01 | 0.02 | -0.01 | 0.02 | -0.01 |
| AMINO ACIDS (BRANCHED-CHAIN) | Isoleucine | Ile | mmol/l | 0.05 | 0.03 | 0.05 | 0.02 | 0.06 | 0.03 | 0.05 | 0.02 | 0.04 | 0.6153 | 0.04 | 0.01 | 0.04 | 0.01 | 0.04 | 0.01 | 0.04 | 0.01 | 0.07 | 0.3670 | -0.01 | 0.02 | -0.01 | 0.02 | -0.01 | 0.03 | -0.02 | 0.02 | -0.04 |
| AMINO ACIDS (BRANCHED-CHAIN) | Leucine | Leu | mmol/l | 0.11 | 0.05 | 0.10 | 0.03 | 0.12 | 0.05 | 0.11 | 0.04 | 0.05 | 0.5916 | 0.09 | 0.02 | 0.09 | 0.02 | 0.09 | 0.02 | 0.09 | 0.02 | 0.07 | 0.4003 | -0.02 | 0.05 | -0.03 | 0.04 | -0.03 | 0.05 | -0.03 | 0.04 | -0.01 |
| AMINO ACIDS (BRANCHED-CHAIN) | Total concentration of branched-chain amino acids (leucine + isoleucine + valine) | Total BCAA | mmol/l | 0.40 | 0.15 | 0.39 | 0.09 | 0.41 | 0.15 | 0.37 | 0.12 | 0.03 | 0.7118 | 0.31 | 0.07 | 0.32 | 0.06 | 0.31 | 0.07 | 0.32 | 0.06 | 0.06 | 0.4325 | -0.08 | 0.14 | -0.08 | 0.11 | -0.07 | 0.15 | -0.09 | 0.10 | -0.03 |
| AMINO ACIDS (BRANCHED-CHAIN) | Valine | Val | mmol/l | 0.23 | 0.07 | 0.23 | 0.05 | 0.24 | 0.08 | 0.22 | 0.05 | 0.03 | 0.7437 | 0.18 | 0.04 | 0.19 | 0.03 | 0.19 | 0.04 | 0.19 | 0.03 | 0.07 | 0.3922 | -0.04 | 0.08 | -0.04 | 0.06 | -0.04 | 0.07 | -0.05 | 0.05 | -0.04 |
| APOLIPOPROTEINS | Apolipoprotein A1 | ApoA1 | g/l | 1.83 | 0.29 | 1.85 | 0.21 | 1.78 | 0.30 | 1.82 | 0.24 | -0.12 | 0.1590 | 2.06 | 0.41 | 2.07 | 0.33 | 2.04 | 0.41 | 2.03 | 0.31 | -0.06 | 0.4981 | 0.24 | 0.37 | 0.24 | 0.30 | 0.24 | 0.37 | 0.19 | 0.26 | -0.06 |
| APOLIPOPROTEINS | Apolipoprotein B | ApoB | g/l | 0.71 | 0.22 | 0.70 | 0.16 | 0.76 | 0.19 | 0.73 | 0.17 | 0.16 | 0.0720 | 1.33 | 0.42 | 1.33 | 0.34 | 1.27 | 0.45 | 1.31 | 0.34 | -0.05 | 0.5571 | 0.59 | 0.35 | 0.62 | 0.30 | 0.53 | 0.38 | 0.52 | 0.26 | -0.18 |
| APOLIPOPROTEINS | Ratio of apolipoprotein B to apolipoprotein A1 | ApoB/ApoA1 | ratio | 0.39 | 0.13 | 0.38 | 0.10 | 0.43 | 0.14 | 0.41 | 0.11 | 0.21 | **0.0144** | 0.63 | 0.25 | 0.67 | 0.24 | 0.63 | 0.22 | 0.66 | 0.19 | 0.01 | 0.9504 | 0.23 | 0.20 | 0.27 | 0.20 | 0.20 | 0.11 | 0.22 | 0.13 | -0.15 |
| CHOLESTEROLS | Clinical LDL cholesterol | Clinical LDL-C | mmol/l | 2.20 | 0.89 | 2.14 | 0.57 | 2.36 | 0.65 | 2.31 | 0.63 | 0.13 | 0.1281 | 3.66 | 1.27 | 3.77 | 1.04 | 3.45 | 1.17 | 3.58 | 1.00 | -0.09 | 0.2704 | 1.51 | 1.21 | 1.55 | 0.89 | 1.26 | 0.95 | 1.13 | 0.87 | -0.23 |
| CHOLESTEROLS | HDL cholesterol | HDL-C | mmol/l | 1.81 | 0.40 | 1.89 | 0.26 | 1.71 | 0.40 | 1.73 | 0.31 | -0.21 | **0.0147** | 1.89 | 0.49 | 1.91 | 0.41 | 1.82 | 0.56 | 1.84 | 0.41 | -0.11 | 0.1720 | 0.11 | 0.45 | 0.11 | 0.36 | 0.11 | 0.44 | 0.06 | 0.32 | -0.06 |
| CHOLESTEROLS | LDL cholesterol | LDL-C | mmol/l | 1.59 | 0.54 | 1.55 | 0.35 | 1.70 | 0.42 | 1.64 | 0.40 | 0.15 | 0.0795 | 2.58 | 0.72 | 2.64 | 0.63 | 2.45 | 0.73 | 2.55 | 0.62 | -0.08 | 0.3046 | 1.03 | 0.72 | 1.05 | 0.55 | 0.88 | 0.62 | 0.79 | 0.53 | -0.24 |
| CHOLESTEROLS | Remnant cholesterol (non-HDL. non-LDL -cholesterol) | Remnant-C | mmol/l | 1.33 | 0.40 | 1.33 | 0.33 | 1.41 | 0.41 | 1.32 | 0.37 | 0.11 | 0.2284 | 2.64 | 0.91 | 2.64 | 0.66 | 2.47 | 0.86 | 2.60 | 0.67 | -0.06 | 0.4514 | 1.27 | 0.75 | 1.32 | 0.59 | 1.09 | 0.77 | 1.11 | 0.53 | -0.19 |
| CHOLESTEROLS | Total cholesterol | Total-C | mmol/l | 4.72 | 0.95 | 4.68 | 0.73 | 4.83 | 1.01 | 4.63 | 0.89 | 0.04 | 0.6908 | 7.08 | 1.54 | 7.20 | 1.30 | 6.78 | 1.66 | 6.98 | 1.35 | -0.11 | 0.1987 | 2.34 | 1.48 | 2.48 | 1.19 | 2.07 | 1.69 | 1.96 | 1.18 | -0.21 |
| CHOLESTEROLS | Total cholesterol minus HDL-C | non-HDL-C | mmol/l | 2.91 | 0.97 | 2.87 | 0.66 | 3.11 | 0.82 | 2.96 | 0.75 | 0.13 | 0.1274 | 5.28 | 1.55 | 5.29 | 1.27 | 4.93 | 1.41 | 5.14 | 1.26 | -0.07 | 0.3908 | 2.24 | 1.34 | 2.37 | 1.12 | 2.09 | 1.32 | 1.90 | 1.03 | -0.22 |
| CHOLESTEROLS | VLDL cholesterol | VLDL-C | mmol/l | 0.48 | 0.24 | 0.46 | 0.17 | 0.55 | 0.27 | 0.52 | 0.20 | 0.20 | **0.0254** | 1.27 | 0.57 | 1.29 | 0.42 | 1.25 | 0.57 | 1.31 | 0.44 | 0.01 | 0.8807 | 0.77 | 0.49 | 0.81 | 0.38 | 0.69 | 0.50 | 0.72 | 0.33 | -0.13 |
| CHOLESTERYL ESTERS | Cholesteryl esters in HDL | HDL-CE | mmol/l | 1.40 | 0.29 | 1.45 | 0.20 | 1.32 | 0.31 | 1.33 | 0.24 | -0.22 | **0.0132** | 1.39 | 0.38 | 1.40 | 0.33 | 1.33 | 0.47 | 1.34 | 0.32 | -0.12 | 0.1413 | 0.02 | 0.35 | 0.01 | 0.29 | 0.00 | 0.33 | -0.02 | 0.25 | -0.06 |
| CHOLESTERYL ESTERS | Total esterified cholesterol | Total-CE | mmol/l | 3.48 | 0.69 | 3.45 | 0.52 | 3.54 | 0.69 | 3.40 | 0.63 | 0.02 | 0.7782 | 5.05 | 1.10 | 5.14 | 0.91 | 4.86 | 1.21 | 4.96 | 0.95 | -0.11 | 0.1644 | 1.60 | 1.08 | 1.66 | 0.84 | 1.34 | 1.20 | 1.28 | 0.84 | -0.20 |
| FATTY ACIDS | Degree of unsaturation | Unsaturation | degree | 1.35 | 0.07 | 1.35 | 0.06 | 1.34 | 0.08 | 1.34 | 0.06 | -0.04 | 0.6293 | 1.25 | 0.08 | 1.25 | 0.06 | 1.22 | 0.09 | 1.23 | 0.07 | -0.20 | **0.0140** | -0.10 | 0.08 | -0.11 | 0.06 | -0.11 | 0.09 | -0.12 | 0.07 | -0.10 |
| FATTY ACIDS | Docosahexaenoic acid | DHA | mmol/l | 0.33 | 0.08 | 0.32 | 0.06 | 0.33 | 0.06 | 0.32 | 0.07 | 0.06 | 0.4765 | 0.38 | 0.11 | 0.39 | 0.10 | 0.38 | 0.14 | 0.38 | 0.10 | -0.03 | 0.7334 | 0.05 | 0.08 | 0.06 | 0.09 | 0.04 | 0.08 | 0.04 | 0.07 | -0.16 |
| FATTY ACIDS | Linoleic acid | LA | mmol/l | 3.96 | 0.64 | 3.90 | 0.57 | 4.03 | 0.73 | 3.89 | 0.66 | 0.06 | 0.5086 | 5.48 | 1.11 | 5.58 | 0.98 | 5.45 | 1.41 | 5.52 | 0.99 | -0.03 | 0.7006 | 1.50 | 0.91 | 1.64 | 0.90 | 1.44 | 1.23 | 1.38 | 0.85 | -0.14 |
| FATTY ACIDS | Monounsaturated fatty acids | MUFA | mmol/l | 2.83 | 0.76 | 2.71 | 0.67 | 3.10 | 0.93 | 2.97 | 0.85 | 0.22 | **0.0141** | 5.53 | 1.67 | 5.49 | 1.28 | 5.50 | 1.93 | 5.83 | 1.55 | 0.11 | 0.1806 | 2.60 | 1.67 | 2.69 | 1.19 | 2.35 | 1.79 | 2.63 | 1.31 | -0.05 |
| FATTY ACIDS | Omega-3 fatty acids | Omega-3 | mmol/l | 0.63 | 0.24 | 0.62 | 0.18 | 0.67 | 0.24 | 0.64 | 0.20 | 0.14 | 0.1094 | 0.83 | 0.31 | 0.86 | 0.27 | 0.86 | 0.42 | 0.89 | 0.26 | 0.08 | 0.3270 | 0.20 | 0.23 | 0.22 | 0.24 | 0.14 | 0.22 | 0.18 | 0.18 | -0.10 |
| FATTY ACIDS | Omega-6 fatty acids | Omega-6 | mmol/l | 4.70 | 0.71 | 4.63 | 0.56 | 4.82 | 0.93 | 4.69 | 0.69 | 0.10 | 0.2702 | 6.36 | 1.13 | 6.49 | 0.97 | 6.43 | 1.57 | 6.46 | 1.00 | -0.02 | 0.8157 | 1.77 | 1.00 | 1.80 | 0.90 | 1.47 | 1.25 | 1.51 | 0.86 | -0.17 |
| FATTY ACIDS | Polyunsaturated fatty acids | PUFA | mmol/l | 5.33 | 0.86 | 5.20 | 0.68 | 5.49 | 1.23 | 5.36 | 0.85 | 0.11 | 0.1972 | 7.25 | 1.33 | 7.35 | 1.12 | 7.23 | 1.77 | 7.35 | 1.15 | 0.00 | 0.9543 | 1.96 | 1.20 | 2.02 | 1.06 | 1.65 | 1.58 | 1.69 | 0.95 | -0.16 |
| FATTY ACIDS | Saturated fatty acids | SFA | mmol/l | 4.27 | 0.81 | 4.17 | 0.87 | 4.47 | 1.23 | 4.34 | 0.92 | 0.14 | 0.1003 | 7.61 | 2.07 | 7.71 | 1.51 | 7.98 | 1.86 | 8.11 | 1.69 | 0.11 | 0.1953 | 3.39 | 1.98 | 3.46 | 1.46 | 3.27 | 1.89 | 3.45 | 1.47 | -0.01 |
| FATTY ACIDS | Total fatty acids | Total-FA | mmol/l | 12.43 | 2.22 | 12.14 | 2.11 | 13.06 | 3.28 | 12.70 | 2.48 | 0.16 | 0.0631 | 20.36 | 5.42 | 20.55 | 3.67 | 20.41 | 4.91 | 21.29 | 4.09 | 0.08 | 0.3517 | 7.91 | 4.60 | 8.18 | 3.46 | 7.79 | 4.63 | 7.77 | 3.42 | -0.07 |
| FATTY ACIDS (RATIOS | Ratio of docosahexaenoic acid to total fatty acids | DHA % | % | 2.64 | 0.59 | 2.60 | 0.39 | 2.56 | 0.56 | 2.58 | 0.39 | -0.08 | 0.3539 | 1.90 | 0.48 | 1.93 | 0.42 | 1.79 | 0.52 | 1.83 | 0.46 | -0.15 | 0.0774 | -0.72 | 0.46 | -0.73 | 0.34 | -0.84 | 0.51 | -0.76 | 0.36 | -0.01 |
| FATTY ACIDS (RATIOS) | Ratio of linoleic acid to total fatty acids | LA % | % | 32.07 | 3.27 | 32.22 | 2.73 | 31.06 | 3.40 | 31.12 | 2.56 | -0.23 | **0.0076** | 27.64 | 3.88 | 27.36 | 3.04 | 26.15 | 4.24 | 26.17 | 3.15 | -0.22 | **0.0079** | -4.58 | 2.96 | -4.74 | 3.06 | -4.49 | 4.00 | -4.78 | 3.04 | 0.00 |
| FATTY ACIDS (RATIOS) | Ratio of monounsaturated fatty acids to total fatty acids | MUFA % | % | 22.57 | 2.93 | 22.45 | 1.87 | 23.47 | 2.87 | 22.85 | 2.50 | 0.20 | **0.0245** | 26.45 | 2.75 | 26.51 | 2.06 | 27.20 | 3.66 | 27.10 | 2.74 | 0.14 | 0.0837 | 3.89 | 2.80 | 4.03 | 2.05 | 3.07 | 2.85 | 3.61 | 2.62 | -0.14 |
| FATTY ACIDS (RATIOS) | Ratio of omega-3 fatty acids to total fatty acids | Omega-3 % | % | 5.03 | 1.62 | 4.96 | 1.01 | 5.12 | 1.14 | 5.07 | 1.00 | 0.06 | 0.4871 | 4.18 | 1.55 | 4.17 | 1.02 | 4.09 | 1.41 | 4.19 | 1.01 | 0.02 | 0.8119 | -0.92 | 1.10 | -0.91 | 0.82 | -1.14 | 1.00 | -1.06 | 0.70 | -0.10 |
| FATTY ACIDS (RATIOS) | Ratio of omega-6 fatty acids to omega-3 fatty acids | Omega-6/Omega-3 | % | 7.93 | 2.54 | 7.77 | 1.86 | 7.57 | 1.73 | 7.41 | 1.73 | -0.12 | 0.1658 | 7.63 | 3.13 | 8.15 | 2.30 | 7.75 | 3.25 | 7.80 | 2.30 | -0.09 | 0.2507 | 0.24 | 1.91 | 0.28 | 1.67 | 0.61 | 2.08 | 0.39 | 1.44 | 0.07 |
| FATTY ACIDS (RATIOS) | Ratio of omega-6 fatty acids to total fatty acids | Omega-6 % | % | 38.17 | 3.00 | 38.67 | 2.78 | 37.26 | 3.68 | 37.51 | 2.69 | -0.22 | **0.0112** | 32.08 | 3.41 | 31.88 | 2.88 | 30.68 | 3.69 | 30.70 | 3.17 | -0.24 | **0.0042** | -6.44 | 3.61 | -6.37 | 3.12 | -6.43 | 4.73 | -6.38 | 3.43 | 0.02 |

| | | | | 1ST TRIMESTER (N=185) | | | | | | | | | | DELIVERY (N=201) | | | | | | | | | | CHANGE BETWEEN DELIVERY AND 1ST TRIMESTER (DELTA) (N=150) | | | | | | | | |
|---|---|---|---|---|---|---|---|---|---|---|---|---|---|---|---|---|---|---|---|---|---|---|---|---|---|---|---|---|---|---|---|---|
| | | | | CONTROL (N=115) | | | | SSRI (N=70) | | | | | | CONTROL (N=112) | | | | SSRI (N=89) | | | | | | CONTROL (N=110) | | | | SSRI (N=40) | | | | |
| GROUP | VARIABLE | ABBRE-VIATION | UNIT | MEDIAN | IQR | MEAN | SD | MEDIAN | IQR | MEAN | SD | r | p | MEDIAN | IQR | MEAN | SD | MEDIAN | IQR | MEAN | SD | r | p | MEDIAN | IQR | MEAN | SD | MEDIAN | IQR | MEAN | SD | r |
| **FATTY ACIDS (RATIOS)** | Ratio of polyunsaturated fatty acids to monounsaturated fatty acids | PUFA/MUFA | % | 1.93 | 0.34 | 1.93 | 0.26 | 1.83 | 0.31 | 1.84 | 0.29 | -0.20 | **0.0200** | 1.38 | 0.26 | 1.37 | 0.20 | 1.27 | 0.30 | 1.31 | 0.25 | -0.21 | **0.0123** | -0.59 | 0.32 | -0.57 | 0.25 | -0.50 | 0.37 | -0.52 | 0.28 | 0.14 |
| **FATTY ACIDS (RATIOS)** | Ratio of polyunsaturated fatty acids to total fatty acids | PUFA % | % | 43.20 | 3.17 | 43.54 | 2.73 | 42.38 | 3.60 | 42.68 | 2.85 | -0.18 | **0.0408** | 36.49 | 3.71 | 36.04 | 2.94 | 34.84 | 3.92 | 34.89 | 3.39 | -0.25 | **0.0027** | -7.18 | 3.25 | -7.27 | 3.22 | -7.18 | 5.73 | -7.44 | 3.51 | 0.01 |
| **FATTY ACIDS (RATIOS)** | Ratio of saturated fatty acids to total fatty acids | SFA % | % | 34.22 | 1.67 | 33.88 | 1.87 | 34.15 | 2.22 | 34.15 | 1.61 | 0.01 | 0.9042 | 37.14 | 1.96 | 37.45 | 1.84 | 37.78 | 2.63 | 38.01 | 2.05 | 0.19 | **0.0178** | 3.14 | 2.00 | 3.24 | 2.21 | 3.51 | 2.98 | 3.83 | 2.28 | 0.10 |
| **FLUID BALANCE** | Albumin | Albumin | g/l | 39.13 | 2.71 | 38.97 | 2.94 | 38.65 | 4.65 | 38.60 | 3.07 | -0.10 | 0.2593 | 28.86 | 3.22 | 28.88 | 2.57 | 28.64 | 4.47 | 28.15 | 3.27 | -0.15 | 0.0718 | -10.29 | 3.73 | -10.31 | 3.27 | -10.56 | 4.39 | -10.89 | 3.45 | -0.09 |
| **FLUID BALANCE** | Creatinine[2,3] | Creatinine | µmol/l | 53.38 | 9.96 | 52.65 | 6.88 | 54.32 | 10.02 | 54.35 | 7.61 | 0.08 | 0.3785 | 65.62 | 12.92 | 65.93 | 9.63 | 64.75 | 14.10 | 64.15 | 11.28 | -0.10 | 0.2209 | 12.80 | 13.89 | 12.53 | 9.26 | 8.44 | 16.51 | 10.08 | 12.84 | -0.13 |
| **FREE CHOLESTEROL** | Free cholesterol in HDL | HDL-FC | mmol/l | 0.41 | 0.11 | 0.43 | 0.07 | 0.39 | 0.08 | 0.40 | 0.08 | -0.17 | 0.0596 | 0.50 | 0.13 | 0.51 | 0.09 | 0.49 | 0.11 | 0.50 | 0.10 | -0.06 | 0.4426 | 0.09 | 0.11 | 0.10 | 0.09 | 0.08 | 0.12 | 0.09 | 0.08 | -0.08 |
| **FREE CHOLESTEROL** | Free cholesterol in LDL | LDL-FC | mmol/l | 0.44 | 0.14 | 0.43 | 0.09 | 0.46 | 0.10 | 0.45 | 0.10 | 0.11 | 0.2072 | 0.65 | 0.19 | 0.67 | 0.16 | 0.61 | 0.19 | 0.63 | 0.16 | -0.12 | 0.1348 | 0.21 | 0.18 | 0.23 | 0.14 | 0.17 | 0.19 | 0.15 | 0.15 | -0.22 |
| **FREE CHOLESTEROL** | Free cholesterol in VLDL | VLDL-FC | mmol/l | 0.19 | 0.09 | 0.18 | 0.08 | 0.23 | 0.11 | 0.21 | 0.09 | 0.22 | **0.0107** | 0.53 | 0.26 | 0.54 | 0.18 | 0.54 | 0.26 | 0.56 | 0.20 | 0.05 | 0.5720 | 0.33 | 0.24 | 0.35 | 0.17 | 0.29 | 0.24 | 0.32 | 0.16 | -0.09 |
| **FREE CHOLESTEROL** | Total free cholesterol | Total-FC | mmol/l | 1.24 | 0.29 | 1.23 | 0.22 | 1.28 | 0.30 | 1.22 | 0.25 | 0.06 | 0.4800 | 2.05 | 0.47 | 2.06 | 0.40 | 1.94 | 0.50 | 2.02 | 0.41 | -0.08 | 0.3429 | 0.74 | 0.43 | 0.82 | 0.36 | 0.66 | 0.49 | 0.68 | 0.34 | -0.20 |
| **GLYCOLYSIS RELATED METABOLITES** | Citrate | Citrate | mmol/l | 0.06 | 0.01 | 0.06 | 0.01 | 0.06 | 0.01 | 0.06 | 0.01 | 0.05 | 0.5343 | 0.08 | 0.02 | 0.08 | 0.01 | 0.08 | 0.02 | 0.09 | 0.02 | 0.14 | 0.0938 | 0.02 | 0.02 | 0.02 | 0.01 | 0.03 | 0.03 | 0.03 | 0.02 | 0.14 |
| **GLYCOLYSIS RELATED METABOLITES** | Glucose | Glucose | mmol/l | 4.72 | 0.93 | 4.62 | 0.86 | 5.03 | 0.88 | 4.80 | 0.85 | 0.24 | **0.0067** | 5.07 | 1.43 | 5.46 | 1.30 | 5.43 | 1.32 | 5.66 | 1.22 | 0.14 | 0.0782 | 0.59 | 1.48 | 0.76 | 1.40 | 0.75 | 1.54 | 0.63 | 1.75 | -0.05 |
| **GLYCOLYSIS RELATED METABOLITES** | Glycerol[4] | Glycerol | mmol/l | 0.10 | 0.04 | 0.10 | 0.03 | 0.11 | 0.05 | 0.09 | 0.04 | 0.01 | 0.9025 | 0.15 | 0.05 | 0.15 | 0.04 | 0.16 | 0.05 | 0.16 | 0.05 | 0.12 | 0.1418 | 0.05 | 0.07 | 0.05 | 0.05 | 0.06 | 0.05 | 0.06 | 0.06 | 0.06 |
| **GLYCOLYSIS RELATED METABOLITES** | Lactate[2] | Lactate | mmol/l | 2.28 | 0.73 | 2.22 | 0.59 | 2.42 | 0.92 | 2.32 | 0.66 | 0.12 | 0.1884 | 3.08 | 1.30 | 3.27 | 1.11 | 3.21 | 1.18 | 3.34 | 0.94 | 0.06 | 0.4797 | 0.98 | 1.48 | 1.02 | 1.15 | 0.99 | 1.18 | 1.07 | 1.16 | 0.02 |
| **GLYCOLYSIS RELATED METABOLITES** | Pyruvate | Pyruvate | mmol/l | 0.05 | 0.04 | 0.05 | 0.03 | 0.06 | 0.05 | 0.05 | 0.04 | 0.10 | 0.2659 | 0.11 | 0.05 | 0.12 | 0.05 | 0.12 | 0.05 | 0.13 | 0.05 | 0.08 | 0.3442 | 0.06 | 0.06 | 0.07 | 0.06 | 0.06 | 0.08 | 0.07 | 0.08 | -0.07 |
| **INFLAMMATION** | Glycoprotein acetyls | GlycA | mmol/l | 0.84 | 0.14 | 0.83 | 0.10 | 0.88 | 0.16 | 0.86 | 0.12 | 0.15 | 0.0963 | 1.00 | 0.12 | 1.01 | 0.10 | 1.06 | 0.14 | 1.05 | 0.12 | 0.23 | **0.0044** | 0.17 | 0.15 | 0.16 | 0.10 | 0.15 | 0.15 | 0.16 | 0.11 | 0.00 |
| **KETONE BODIES** | 3-Hydroxybutyrate | bOHbutyrate | mmol/l | 0.04 | 0.02 | 0.03 | 0.04 | 0.04 | 0.03 | 0.03 | 0.05 | 0.04 | 0.6567 | 0.08 | 0.11 | 0.15 | 0.19 | 0.09 | 0.14 | 0.15 | 0.14 | 0.03 | 0.7593 | 0.05 | 0.11 | 0.11 | 0.19 | 0.06 | 0.12 | 0.11 | 0.16 | 0.06 |
| **KETONE BODIES** | Acetate | Acetate | mmol/l | 0.03 | 0.01 | 0.03 | 0.01 | 0.03 | 0.01 | 0.03 | 0.01 | 0.01 | 0.8863 | 0.02 | 0.01 | 0.03 | 0.01 | 0.02 | 0.01 | 0.03 | 0.01 | -0.05 | 0.5786 | 0.00 | 0.02 | 0.00 | 0.02 | -0.01 | 0.02 | 0.00 | 0.01 | -0.11 |
| **KETONE BODIES** | Acetoacetate | Acetoacetate | mmol/l | 0.02 | 0.01 | 0.02 | 0.01 | 0.03 | 0.01 | 0.02 | 0.02 | 0.19 | **0.0315** | 0.04 | 0.04 | 0.06 | 0.06 | 0.04 | 0.04 | 0.06 | 0.04 | -0.04 | 0.6384 | 0.02 | 0.04 | 0.04 | 0.06 | 0.02 | 0.04 | 0.03 | 0.05 | -0.09 |
| **KETONE BODIES** | Acetone | Acetone | mmol/l | 0.02 | 0.00 | 0.01 | 0.00 | 0.02 | 0.00 | 0.02 | 0.00 | 0.11 | 0.1933 | 0.02 | 0.01 | 0.02 | 0.01 | 0.02 | 0.01 | 0.02 | 0.01 | -0.02 | 0.8138 | 0.00 | 0.01 | 0.00 | 0.01 | 0.00 | 0.01 | 0.00 | 0.01 | -0.10 |
| **LIPOPROTEIN PARTICLE CONCENTRATIONS** | Total concentration of lipoprotein particles | Total-P | mmol/l | 0.02 | 0.00 | 0.02 | 0.00 | 0.02 | 0.00 | 0.02 | 0.00 | -0.03 | 0.7055 | 0.02 | 0.00 | 0.02 | 0.00 | 0.02 | 0.00 | 0.02 | 0.00 | -0.06 | 0.4470 | 0.00 | 0.00 | 0.00 | 0.00 | 0.00 | 0.00 | 0.00 | 0.00 | -0.10 |
| **LIPOPROTEIN PARTICLE SIZES** | Average diameter for HDL particles | HDL size | nm | 9.93 | 0.24 | 9.95 | 0.18 | 9.86 | 0.27 | 9.87 | 0.20 | -0.24 | **0.0069** | 10.01 | 0.27 | 10.00 | 0.18 | 9.96 | 0.24 | 9.99 | 0.20 | -0.07 | 0.4240 | 0.06 | 0.18 | 0.07 | 0.15 | 0.09 | 0.26 | 0.11 | 0.18 | 0.07 |
| **LIPOPROTEIN PARTICLE SIZES** | Average diameter for LDL particles | LDL size | nm | 23.96 | 0.08 | 23.97 | 0.07 | 23.95 | 0.08 | 23.96 | 0.06 | -0.13 | 0.1295 | 23.98 | 0.06 | 23.96 | 0.06 | 23.95 | 0.09 | 23.94 | 0.08 | -0.23 | **0.0054** | -0.01 | 0.10 | 0.00 | 0.09 | -0.03 | 0.11 | -0.04 | 0.09 | -0.17 |
| **LIPOPROTEIN PARTICLE SIZES** | Average diameter for VLDL particles | VLDL size | nm | 37.34 | 1.69 | 37.18 | 1.15 | 37.90 | 1.58 | 37.84 | 1.28 | 0.26 | **0.0036** | 38.32 | 1.17 | 38.36 | 0.96 | 38.55 | 1.79 | 38.61 | 1.31 | 0.12 | 0.1508 | 1.15 | 1.88 | 1.06 | 1.18 | 0.82 | 1.33 | 0.77 | 1.15 | -0.15 |
| **LIPOPROTEIN SUBCLASSES** | Cholesteryl esters in chylomicrons and extremely large VLDL | XXL-VLDL-CE | mmol/l | 0.01 | 0.01 | 0.00 | 0.01 | 0.01 | 0.02 | 0.01 | 0.01 | 0.30 | **0.0010** | 0.05 | 0.04 | 0.05 | 0.03 | 0.05 | 0.04 | 0.06 | 0.03 | 0.14 | 0.0864 | 0.04 | 0.03 | 0.04 | 0.03 | 0.04 | 0.03 | 0.05 | 0.03 | 0.02 |
| **LIPOPROTEIN SUBCLASSES** | Cholesteryl esters in IDL | IDL-CE | mmol/l | 0.63 | 0.17 | 0.62 | 0.13 | 0.65 | 0.16 | 0.62 | 0.15 | 0.02 | 0.8485 | 0.99 | 0.26 | 1.02 | 0.22 | 0.94 | 0.28 | 0.97 | 0.23 | -0.12 | 0.1355 | 0.36 | 0.26 | 0.38 | 0.20 | 0.32 | 0.30 | 0.29 | 0.21 | -0.21 |
| **LIPOPROTEIN SUBCLASSES** | Cholesteryl esters in large HDL | L-HDL-CE | mmol/l | 0.42 | 0.18 | 0.45 | 0.12 | 0.38 | 0.20 | 0.39 | 0.14 | -0.25 | **0.0050** | 0.46 | 0.21 | 0.47 | 0.16 | 0.43 | 0.20 | 0.44 | 0.17 | -0.12 | 0.1579 | 0.05 | 0.17 | 0.04 | 0.14 | 0.04 | 0.20 | 0.04 | 0.13 | -0.02 |
| **LIPOPROTEIN SUBCLASSES** | Cholesteryl esters in large LDL | L-LDL-CE | mmol/l | 0.80 | 0.25 | 0.80 | 0.17 | 0.85 | 0.24 | 0.81 | 0.20 | 0.14 | 0.1205 | 1.31 | 0.35 | 1.32 | 0.30 | 1.22 | 0.40 | 1.26 | 0.29 | -0.10 | 0.2264 | 0.49 | 0.36 | 0.51 | 0.27 | 0.39 | 0.36 | 0.37 | 0.26 | -0.25 |
| **LIPOPROTEIN SUBCLASSES** | Cholesteryl esters in large VLDL | L-VLDL-CE | mmol/l | 0.03 | 0.02 | 0.03 | 0.02 | 0.04 | 0.03 | 0.04 | 0.02 | 0.22 | **0.0104** | 0.09 | 0.05 | 0.10 | 0.04 | 0.10 | 0.06 | 0.10 | 0.05 | 0.05 | 0.5571 | 0.06 | 0.05 | 0.07 | 0.04 | 0.05 | 0.05 | 0.06 | 0.04 | -0.07 |
| **LIPOPROTEIN SUBCLASSES** | Cholesteryl esters in LDL | LDL-CE | mmol/l | 1.15 | 0.40 | 1.13 | 0.26 | 1.24 | 0.33 | 1.19 | 0.30 | 0.17 | 0.0506 | 1.95 | 0.55 | 1.98 | 0.47 | 1.89 | 0.54 | 1.92 | 0.46 | -0.07 | 0.3855 | 0.80 | 0.53 | 0.82 | 0.41 | 0.70 | 0.49 | 0.63 | 0.39 | -0.24 |
| **LIPOPROTEIN SUBCLASSES** | Concentration of chylomicrons and extremely large VLDL particles | XXL-VLDL-P | mmol/l | 0.0000004 | 0.0000004 | 0.00000003 | 0.0000007 | 0.0000007 | 0.0000010 | 0.0000003 | 0.0000009 | 0.31 | **0.0010** | 0.000002 | 0.000002 | 0.000003 | 0.000002 | 0.000003 | 0.000003 | 0.000003 | 0.000002 | 0.19 | **0.0215** | 0.000002 | 0.000002 | 0.000002 | 0.000002 | 0.000002 | 0.000003 | 0.000003 | 0.000002 | 0.07 |
| **LIPOPROTEIN SUBCLASSES** | Concentration of HDL particles | HDL-P | mmol/l | 0.018 | 0.003 | 0.0183 | 0.002 | 0.02 | 0.003 | 0.02 | 0.002 | -0.05 | 0.5399 | 0.02 | 0.004 | 0.02 | 0.003 | 0.02 | 0.004 | 0.02 | 0.003 | -0.06 | 0.4441 | 0.002 | 0.004 | 0.002 | 0.003 | 0.002 | 0.004 | 0.001 | 0.003 | -0.08 |
| **LIPOPROTEIN SUBCLASSES** | Concentration of IDL particles | IDL-P | mmol/l | 0.0003 | 0.00008 | 0.0003 | 0.00006 | 0.0003 | 0.00007 | 0.0003 | 0.00006 | -0.05 | 0.6034 | 0.0005 | 0.0001 | 0.0005 | 0.0001 | 0.0005 | 0.0001 | 0.0005 | 0.0001 | -0.08 | 0.3128 | 0.0002 | 0.0001 | 0.0002 | 0.0001 | 0.0002 | 0.0001 | 0.0002 | 0.0001 | -0.19 |
| **LIPOPROTEIN SUBCLASSES** | Concentration of large HDL particles | L-HDL-P | mmol/l | 0.003 | 0.001 | 0.0028 | 0.001 | 0.002 | 0.001 | 0.002 | 0.001 | -0.23 | **0.0079** | 0.003 | 0.001 | 0.003 | 0.001 | 0.003 | 0.001 | 0.003 | 0.001 | -0.09 | 0.2538 | 0.0006 | 0.001 | 0.0006 | 0.0008 | 0.0005 | 0.0013 | 0.0006 | 0.0008 | -0.04 |
| **LIPOPROTEIN SUBCLASSES** | Concentration of large LDL particles | L-LDL-P | mmol/l | 0.001 | 0.0002 | 0.0006 | 0.0001 | 0.001 | 0.0002 | 0.001 | 0.0002 | 0.13 | 0.1274 | 0.001 | 0.0004 | 0.001 | 0.0003 | 0.001 | 0.0004 | 0.001 | 0.0003 | -0.07 | 0.3922 | 0.0005 | 0.0003 | 0.0005 | 0.0003 | 0.0004 | 0.0004 | 0.0004 | 0.0002 | -0.20 |
| **LIPOPROTEIN SUBCLASSES** | Concentration of large VLDL particles | L-VLDL-P | mmol/l | 0.00001 | 0.00001 | 0.000006 | 0.000004 | 0.000009 | 0.000007 | 0.000008 | 0.000005 | -0.26 | **0.0035** | 0.00002 | 0.00001 | 0.00002 | 0.00001 | 0.00002 | 0.00002 | 0.00002 | 0.00001 | 0.11 | 0.1919 | 0.000013 | 0.000011 | 0.000014 | 0.000009 | 0.000011 | 0.000011 | 0.000014 | 0.000010 | -0.04 |
| **LIPOPROTEIN SUBCLASSES** | Concentration of LDL particles | LDL-P | mmol/l | 0.0010 | 0.0003 | 0.0010 | 0.0002 | 0.001 | 0.0003 | 0.001 | 0.0002 | 0.17 | **0.0499** | 0.002 | 0.001 | 0.002 | 0.0005 | 0.002 | 0.001 | 0.002 | 0.0005 | -0.05 | 0.5201 | 0.0008 | 0.0005 | 0.0008 | 0.0004 | 0.0007 | 0.0005 | 0.0007 | 0.0004 | -0.19 |
| **LIPOPROTEIN SUBCLASSES** | Concentration of medium HDL particles | M-HDL-P | mmol/l | 0.0052 | 0.001 | 0.0053 | 0.001 | 0.005 | 0.001 | 0.005 | 0.001 | -0.11 | 0.2197 | 0.01 | 0.001 | 0.01 | 0.001 | 0.01 | 0.002 | 0.01 | 0.001 | -0.07 | 0.4240 | 0.0004 | 0.0013 | 0.0004 | 0.0011 | 0.0005 | 0.0013 | 0.0003 | 0.0009 | -0.05 |
| **LIPOPROTEIN SUBCLASSES** | Concentration of medium LDL particles | M-LDL-P | mmol/l | 0.0002 | 0.00009 | 0.0002 | 0.00006 | 0.0003 | 0.00006 | 0.0002 | 0.00006 | 0.22 | **0.0137** | 0.0004 | 0.0001 | 0.0004 | 0.0001 | 0.0004 | 0.0002 | 0.0004 | 0.0001 | -0.02 | 0.8251 | 0.0002 | 0.0001 | 0.0002 | 0.0001 | 0.0002 | 0.0001 | 0.0002 | 0.0001 | -0.16 |
| **LIPOPROTEIN SUBCLASSES** | Concentration of medium VLDL particles | M-VLDL-P | mmol/l | 0.00003 | 0.00001 | 0.00002 | 0.000009 | 0.00003 | 0.000014 | 0.00003 | 0.000011 | 0.21 | **0.0192** | 0.00006 | 0.00003 | 0.00006 | 0.00002 | 0.0001 | 0.00003 | 0.0001 | 0.00002 | -0.02 | 0.8432 | 0.00004 | 0.00003 | 0.00004 | 0.00002 | 0.00003 | 0.00002 | 0.00003 | 0.00002 | -0.13 |
| **LIPOPROTEIN SUBCLASSES** | Concentration of small HDL particles | S-HDL-P | mmol/l | 0.0100 | 0.001 | 0.0100 | 0.001 | 0.010 | 0.0015 | 0.010 | 0.0013 | 0.10 | 0.2533 | 0.01 | 0.002 | 0.01 | 0.001 | 0.01 | 0.002 | 0.01 | 0.002 | 0.00 | 0.9679 | 0.0006 | 0.0019 | 0.0008 | 0.0014 | 0.0002 | 0.0022 | 0.0003 | 0.0017 | -0.13 |
| **LIPOPROTEIN SUBCLASSES** | Concentration of small LDL particles | S-LDL-P | mmol/l | 0.0001 | 0.00004 | 0.0001 | 0.00003 | 0.0002 | 0.00003 | 0.0001 | 0.00003 | 0.20 | **0.0243** | 0.0002 | 0.0001 | 0.0003 | 0.0001 | 0.0002 | 0.0001 | 0.0003 | 0.0001 | 0.00 | 0.9844 | 0.0001 | 0.0001 | 0.0001 | 0.0001 | 0.0001 | 0.0001 | 0.0001 | 0.0000 | -0.13 |

| GROUP | VARIABLE | ABBRE-VIATION | UNIT | 1ST TRIMESTER (N=185) | | | | | | | | | | DELIVERY (N=201) | | | | | | | | | | CHANGE BETWEEN DELIVERY AND 1ST TRIMESTER (DELTA) (N=150) | | | | | | | | |
|---|---|---|---|---|---|---|---|---|---|---|---|---|---|---|---|---|---|---|---|---|---|---|---|---|---|---|---|---|---|---|---|---|
| | | | | CONTROL (N=115) | | | | SSRI (N=70) | | | | | | CONTROL (N=112) | | | | SSRI (N=89) | | | | | | CONTROL (N=110) | | | | SSRI (N=40) | | | | |
| | | | | MEDIAN | IQR | MEAN | SD | MEDIAN | IQR | MEAN | SD | r | p | MEDIAN | IQR | MEAN | SD | MEDIAN | IQR | MEAN | SD | r | p | MEDIAN | IQR | MEAN | SD | MEDIAN | IQR | MEAN | SD | r |
| LIPOPROTEIN SUBCLASSES | Concentration of small VLDL particles | S-VLDL-P | mmol/l | 0.00003 | 0.00001 | 0.0000 | 0.00001 | 0.00004 | 0.00002 | 0.00004 | 0.00001 | 0.19 | **0.0291** | 0.00008 | 0.00004 | 0.00009 | 0.00003 | 0.0001 | 0.00004 | 0.0001 | 0.00003 | 0.04 | 0.6099 | 0.00005 | 0.00003 | 0.00005 | 0.00003 | 0.00004 | 0.00004 | 0.00005 | 0.00002 | -0.08 |
| LIPOPROTEIN SUBCLASSES | Concentration of very large HDL particles | XL-HDL-P | mmol/l | 0.0004 | 0.0002 | 0.0004 | 0.0001 | 0.0003 | 0.00015 | 0.0003 | 0.00013 | -0.19 | **0.0322** | 0.0005 | 0.0002 | 0.0005 | 0.0001 | 0.001 | 0.0002 | 0.001 | 0.0002 | -0.07 | 0.4058 | 0.0002 | 0.0001 | 0.0002 | 0.0001 | 0.0002 | 0.0002 | 0.0002 | 0.0001 | 0.02 |
| LIPOPROTEIN SUBCLASSES | Concentration of very large VLDL particles | XL-VLDL-P | mmol/l | 0.000002 | 0.000002 | 0.000002 | 0.000002 | 0.000003 | 0.00000 | 0.000002 | 0.00000 | 0.28 | **0.0015** | 0.000007 | 0.000004 | 0.000007 | 0.000004 | 0.000008 | 0.00001 | 0.00001 | 0.000004 | 0.14 | 0.0933 | 0.000005 | 0.000004 | 0.000005 | 0.000003 | 0.000004 | 0.000005 | 0.000006 | 0.000004 | -0.01 |
| LIPOPROTEIN SUBCLASSES | Concentration of very small VLDL particles | XS-VLDL-P | mmol/l | 0.0001 | 0.00002 | 0.0001 | 0.00001 | 0.0001 | 0.00002 | 0.0001 | 0.00001 | 0.07 | 0.4052 | 0.0001 | 0.00004 | 0.0001 | 0.00003 | 0.0001 | 0.00004 | 0.0001 | 0.00003 | 0.00 | 0.9786 | 0.00006 | 0.00003 | 0.00006 | 0.00003 | 0.00005 | 0.00003 | 0.00006 | 0.00002 | -0.13 |
| LIPOPROTEIN SUBCLASSES | Concentration of VLDL particles | VLDL-P | mmol/l | 0.0001 | 0.00005 | 0.0001 | 0.00004 | 0.0001 | 0.00005 | 0.0001 | 0.00004 | 0.20 | **0.0259** | 0.0003 | 0.0001 | 0.0003 | 0.0001 | 0.0003 | 0.0001 | 0.0003 | 0.0001 | 0.04 | 0.6254 | 0.0002 | 0.0001 | 0.0002 | 0.00008 | 0.0001 | 0.0001 | 0.0002 | 0.00007 | -0.09 |
| LIPOPROTEIN SUBCLASSES | Free cholesterol in chylomicrons and extremely large VLDL | XXL-VLDL-FC | mmol/l | 0.005 | 0.004 | 0.0013 | 0.007 | 0.008 | 0.01 | 0.005 | 0.01 | 0.28 | **0.0012** | 0.03 | 0.02 | 0.03 | 0.02 | 0.04 | 0.04 | 0.041278 | 0.000002 | 0.16 | 0.0552 | 0.03 | 0.02 | 0.03 | 0.02 | 0.03 | 0.02 | 0.03 | 0.03 | 0.03 |
| LIPOPROTEIN SUBCLASSES | Free cholesterol in IDL | IDL-FC | mmol/l | 0.21 | 0.05 | 0.20 | 0.04 | 0.21 | 0.05 | 0.21 | 0.05 | 0.00 | 0.9898 | 0.33 | 0.09 | 0.34 | 0.07 | 0.32 | 0.11 | 0.32 | 0.08 | -0.12 | 0.1304 | 0.12 | 0.09 | 0.13 | 0.07 | 0.10 | 0.10 | 0.10 | 0.07 | -0.21 |
| LIPOPROTEIN SUBCLASSES | Free cholesterol in large HDL | L-HDL-FC | mmol/l | 0.13 | 0.05 | 0.13 | 0.04 | 0.11 | 0.06 | 0.12 | 0.04 | -0.23 | **0.0098** | 0.15 | 0.07 | 0.15 | 0.05 | 0.14 | 0.05 | 0.15 | 0.05 | -0.10 | 0.2408 | 0.03 | 0.05 | 0.03 | 0.04 | 0.02 | 0.06 | 0.03 | 0.04 | -0.04 |
| LIPOPROTEIN SUBCLASSES | Free cholesterol in large LDL | L-LDL-FC | mmol/l | 0.29 | 0.09 | 0.28 | 0.06 | 0.30 | 0.06 | 0.29 | 0.07 | 0.08 | 0.3480 | 0.43 | 0.12 | 0.43 | 0.10 | 0.40 | 0.13 | 0.41 | 0.10 | -0.14 | 0.0841 | 0.13 | 0.12 | 0.15 | 0.09 | 0.12 | 0.13 | 0.10 | 0.10 | -0.23 |
| LIPOPROTEIN SUBCLASSES | Free cholesterol in large VLDL | L-VLDL-FC | mmol/l | 0.03 | 0.02 | 0.02 | 0.02 | 0.04 | 0.03 | 0.03 | 0.02 | 0.27 | **0.0021** | 0.08 | 0.05 | 0.09 | 0.04 | 0.09 | 0.07 | 0.10 | 0.05 | 0.10 | 0.2208 | 0.06 | 0.05 | 0.06 | 0.04 | 0.04 | 0.05 | 0.06 | 0.04 | -0.06 |
| LIPOPROTEIN SUBCLASSES | Free cholesterol in medium HDL | M-HDL-FC | mmol/l | 0.13 | 0.03 | 0.13 | 0.02 | 0.12 | 0.03 | 0.12 | 0.02 | -0.12 | 0.1764 | 0.15 | 0.04 | 0.14 | 0.03 | 0.14 | 0.04 | 0.14 | 0.03 | -0.06 | 0.4797 | 0.02 | 0.04 | 0.02 | 0.03 | 0.02 | 0.04 | 0.01 | 0.03 | -0.07 |
| LIPOPROTEIN SUBCLASSES | Free cholesterol in medium LDL | M-LDL-FC | mmol/l | 0.11 | 0.04 | 0.10 | 0.03 | 0.11 | 0.03 | 0.11 | 0.03 | 0.16 | 0.0747 | 0.16 | 0.05 | 0.17 | 0.05 | 0.16 | 0.05 | 0.16 | 0.04 | -0.10 | 0.2127 | 0.06 | 0.05 | 0.06 | 0.04 | 0.05 | 0.05 | 0.04 | 0.04 | -0.22 |
| LIPOPROTEIN SUBCLASSES | Free cholesterol in medium VLDL | M-VLDL-FC | mmol/l | 0.05 | 0.03 | 0.05 | 0.02 | 0.06 | 0.03 | 0.06 | 0.02 | 0.19 | **0.0290** | 0.14 | 0.07 | 0.14 | 0.05 | 0.14 | 0.07 | 0.14 | 0.05 | -0.01 | 0.8884 | 0.08 | 0.06 | 0.09 | 0.05 | 0.07 | 0.06 | 0.08 | 0.04 | -0.15 |
| LIPOPROTEIN SUBCLASSES | Free cholesterol in small HDL | S-HDL-FC | mmol/l | 0.13 | 0.02 | 0.13 | 0.01 | 0.13 | 0.02 | 0.13 | 0.02 | 0.08 | 0.3918 | 0.18 | 0.03 | 0.17 | 0.02 | 0.17 | 0.03 | 0.17 | 0.02 | 0.01 | 0.9484 | 0.04 | 0.03 | 0.04 | 0.02 | 0.04 | 0.03 | 0.04 | 0.02 | -0.09 |
| LIPOPROTEIN SUBCLASSES | Free cholesterol in small LDL | S-LDL-FC | mmol/l | 0.04 | 0.02 | 0.04 | 0.01 | 0.05 | 0.01 | 0.05 | 0.01 | 0.13 | 0.1484 | 0.06 | 0.02 | 0.06 | 0.02 | 0.06 | 0.02 | 0.06 | 0.02 | -0.08 | 0.3210 | 0.02 | 0.02 | 0.02 | 0.01 | 0.02 | 0.01 | 0.02 | 0.01 | -0.17 |
| LIPOPROTEIN SUBCLASSES | Free cholesterol in small VLDL | S-VLDL-FC | mmol/l | 0.04 | 0.02 | 0.04 | 0.01 | 0.05 | 0.02 | 0.05 | 0.02 | 0.16 | 0.0647 | 0.11 | 0.05 | 0.11 | 0.04 | 0.10 | 0.05 | 0.11 | 0.04 | -0.04 | 0.6454 | 0.06 | 0.04 | 0.07 | 0.03 | 0.05 | 0.05 | 0.06 | 0.03 | -0.15 |
| LIPOPROTEIN SUBCLASSES | Free cholesterol in very large HDL | XL-HDL-FC | mmol/l | 0.03 | 0.01 | 0.03 | 0.01 | 0.03 | 0.01 | 0.03 | 0.01 | -0.18 | **0.0392** | 0.04 | 0.01 | 0.04 | 0.01 | 0.04 | 0.01 | 0.04 | 0.01 | -0.08 | 0.3023 | 0.01 | 0.01 | 0.01 | 0.01 | 0.01 | 0.01 | 0.01 | 0.01 | 0.03 |
| LIPOPROTEIN SUBCLASSES | Free cholesterol in very large VLDL | XL-VLDL-FC | mmol/l | 0.01 | 0.01 | 0.01 | 0.01 | 0.02 | 0.01 | 0.01 | 0.01 | 0.29 | **0.0011** | 0.04 | 0.03 | 0.04 | 0.02 | 0.04 | 0.04 | 0.05 | 0.03 | 0.10 | 0.2021 | 0.03 | 0.03 | 0.03 | 0.02 | 0.03 | 0.03 | 0.03 | 0.02 | -0.04 |
| LIPOPROTEIN SUBCLASSES | Free cholesterol in very small VLDL | XS-VLDL-FC | mmol/l | 0.05 | 0.02 | 0.05 | 0.01 | 0.05 | 0.02 | 0.05 | 0.01 | 0.07 | 0.4239 | 0.12 | 0.04 | 0.12 | 0.03 | 0.12 | 0.04 | 0.12 | 0.03 | -0.01 | 0.9484 | 0.07 | 0.03 | 0.07 | 0.03 | 0.06 | 0.03 | 0.06 | 0.03 | -0.12 |
| LIPOPROTEIN SUBCLASSES | Phospholipids in chylomicrons and extremely large VLDL | XXL-VLDL-PL | mmol/l | 0.00 | 0.00 | 0.00 | 0.01 | 0.01 | 0.01 | 0.00 | 0.02 | 0.31 | **0.0010** | 0.04 | 0.04 | 0.05 | 0.03 | 0.06 | 0.05 | 0.06 | 0.04 | 0.17 | **0.0383** | 0.04 | 0.04 | 0.05 | 0.03 | 0.04 | 0.04 | 0.05 | 0.04 | 0.05 |
| LIPOPROTEIN SUBCLASSES | Phospholipids in HDL | HDL-PL | mmol/l | 2.04 | 0.41 | 2.09 | 0.28 | 1.97 | 0.42 | 2.02 | 0.32 | -0.14 | 0.1051 | 2.30 | 0.52 | 2.30 | 0.42 | 2.25 | 0.51 | 2.26 | 0.39 | -0.06 | 0.4997 | 0.27 | 0.49 | 0.26 | 0.39 | 0.23 | 0.47 | 0.21 | 0.32 | -0.06 |
| LIPOPROTEIN SUBCLASSES | Phospholipids in IDL | IDL-PL | mmol/l | 0.28 | 0.08 | 0.28 | 0.06 | 0.29 | 0.07 | 0.28 | 0.06 | 0.04 | 0.6763 | 0.47 | 0.12 | 0.47 | 0.10 | 0.44 | 0.15 | 0.45 | 0.11 | -0.10 | 0.2302 | 0.18 | 0.11 | 0.19 | 0.09 | 0.13 | 0.12 | 0.15 | 0.10 | -0.23 |
| LIPOPROTEIN SUBCLASSES | Phospholipids in large HDL | L-HDL-PL | mmol/l | 0.57 | 0.22 | 0.61 | 0.14 | 0.52 | 0.20 | 0.54 | 0.16 | -0.23 | **0.0092** | 0.65 | 0.23 | 0.65 | 0.19 | 0.62 | 0.21 | 0.62 | 0.19 | -0.09 | 0.2768 | 0.09 | 0.22 | 0.08 | 0.16 | 0.08 | 0.23 | 0.07 | 0.15 | -0.03 |
| LIPOPROTEIN SUBCLASSES | Phospholipids in large LDL | L-LDL-PL | mmol/l | 0.33 | 0.10 | 0.32 | 0.07 | 0.34 | 0.07 | 0.33 | 0.07 | 0.12 | 0.1667 | 0.51 | 0.13 | 0.52 | 0.12 | 0.49 | 0.14 | 0.50 | 0.12 | -0.09 | 0.2833 | 0.19 | 0.13 | 0.19 | 0.10 | 0.16 | 0.13 | 0.15 | 0.10 | -0.21 |
| LIPOPROTEIN SUBCLASSES | Phospholipids in large VLDL | L-VLDL-PL | mmol/l | 0.04 | 0.04 | 0.03 | 0.03 | 0.06 | 0.05 | 0.05 | 0.04 | 0.25 | **0.0044** | 0.13 | 0.08 | 0.14 | 0.07 | 0.14 | 0.11 | 0.16 | 0.08 | 0.12 | 0.1543 | 0.10 | 0.08 | 0.10 | 0.06 | 0.08 | 0.08 | 0.10 | 0.07 | -0.03 |
| LIPOPROTEIN SUBCLASSES | Phospholipids in LDL | LDL-PL | mmol/l | 0.55 | 0.18 | 0.53 | 0.11 | 0.58 | 0.14 | 0.57 | 0.12 | 0.15 | 0.0839 | 0.88 | 0.25 | 0.90 | 0.21 | 0.85 | 0.23 | 0.88 | 0.21 | -0.07 | 0.4142 | 0.34 | 0.24 | 0.36 | 0.18 | 0.31 | 0.19 | 0.28 | 0.18 | -0.20 |
| LIPOPROTEIN SUBCLASSES | Phospholipids in medium HDL | M-HDL-PL | mmol/l | 0.62 | 0.11 | 0.61 | 0.08 | 0.61 | 0.13 | 0.60 | 0.09 | -0.05 | 0.5399 | 0.67 | 0.17 | 0.67 | 0.13 | 0.66 | 0.14 | 0.66 | 0.11 | -0.02 | 0.7855 | 0.05 | 0.14 | 0.05 | 0.12 | 0.03 | 0.14 | 0.03 | 0.10 | -0.06 |
| LIPOPROTEIN SUBCLASSES | Phospholipids in medium LDL | M-LDL-PL | mmol/l | 0.14 | 0.05 | 0.14 | 0.03 | 0.15 | 0.04 | 0.15 | 0.04 | 0.19 | **0.0269** | 0.25 | 0.08 | 0.25 | 0.07 | 0.24 | 0.08 | 0.25 | 0.06 | -0.03 | 0.6952 | 0.10 | 0.07 | 0.11 | 0.06 | 0.09 | 0.07 | 0.09 | 0.05 | -0.17 |
| LIPOPROTEIN SUBCLASSES | Phospholipids in medium VLDL | M-VLDL-PL | mmol/l | 0.09 | 0.05 | 0.09 | 0.04 | 0.11 | 0.05 | 0.10 | 0.04 | 0.20 | **0.0226** | 0.24 | 0.12 | 0.24 | 0.08 | 0.24 | 0.11 | 0.25 | 0.09 | 0.01 | 0.8961 | 0.14 | 0.10 | 0.15 | 0.08 | 0.13 | 0.09 | 0.13 | 0.07 | -0.14 |
| LIPOPROTEIN SUBCLASSES | Phospholipids in small HDL | S-HDL-PL | mmol/l | 0.71 | 0.09 | 0.71 | 0.07 | 0.72 | 0.12 | 0.71 | 0.10 | 0.09 | 0.3238 | 0.78 | 0.15 | 0.80 | 0.11 | 0.80 | 0.12 | 0.80 | 0.11 | 0.05 | 0.5425 | 0.08 | 0.14 | 0.09 | 0.11 | 0.07 | 0.13 | 0.06 | 0.10 | -0.10 |
| LIPOPROTEIN SUBCLASSES | Phospholipids in small LDL | S-LDL-PL | mmol/l | 0.08 | 0.02 | 0.08 | 0.02 | 0.08 | 0.02 | 0.08 | 0.02 | 0.14 | 0.1185 | 0.13 | 0.04 | 0.13 | 0.03 | 0.13 | 0.04 | 0.13 | 0.03 | -0.01 | 0.9290 | 0.05 | 0.04 | 0.05 | 0.03 | 0.04 | 0.03 | 0.05 | 0.02 | -0.14 |
| LIPOPROTEIN SUBCLASSES | Phospholipids in small VLDL | S-VLDL-PL | mmol/l | 0.08 | 0.03 | 0.07 | 0.02 | 0.09 | 0.03 | 0.08 | 0.03 | 0.18 | **0.0437** | 0.19 | 0.08 | 0.19 | 0.06 | 0.19 | 0.09 | 0.19 | 0.06 | 0.00 | 0.9659 | 0.11 | 0.07 | 0.11 | 0.05 | 0.10 | 0.08 | 0.10 | 0.05 | -0.11 |
| LIPOPROTEIN SUBCLASSES | Phospholipids in very large HDL | XL-HDL-PL | mmol/l | 0.14 | 0.07 | 0.14 | 0.05 | 0.12 | 0.06 | 0.12 | 0.06 | -0.22 | **0.0109** | 0.18 | 0.08 | 0.18 | 0.06 | 0.17 | 0.07 | 0.17 | 0.06 | -0.10 | 0.2172 | 0.04 | 0.06 | 0.04 | 0.05 | 0.04 | 0.07 | 0.05 | 0.05 | 0.02 |
| LIPOPROTEIN SUBCLASSES | Phospholipids in very large VLDL | XL-VLDL-PL | mmol/l | 0.02 | 0.02 | 0.01 | 0.02 | 0.03 | 0.02 | 0.02 | 0.02 | 0.29 | **0.0011** | 0.07 | 0.05 | 0.07 | 0.04 | 0.08 | 0.07 | 0.09 | 0.05 | 0.12 | 0.1368 | 0.06 | 0.05 | 0.06 | 0.04 | 0.04 | 0.05 | 0.06 | 0.04 | -0.02 |
| LIPOPROTEIN SUBCLASSES | Phospholipids in very small VLDL | XS-VLDL-PL | mmol/l | 0.10 | 0.03 | 0.10 | 0.03 | 0.10 | 0.04 | 0.10 | 0.03 | 0.08 | 0.3871 | 0.23 | 0.08 | 0.23 | 0.06 | 0.23 | 0.09 | 0.24 | 0.07 | 0.00 | 0.9581 | 0.13 | 0.07 | 0.14 | 0.05 | 0.12 | 0.06 | 0.13 | 0.05 | -0.09 |
| LIPOPROTEIN SUBCLASSES | Phospholipids in VLDL | VLDL-PL | mmol/l | 0.33 | 0.17 | 0.31 | 0.13 | 0.39 | 0.20 | 0.36 | 0.15 | 0.22 | **0.0112** | 0.91 | 0.44 | 0.93 | 0.32 | 0.95 | 0.51 | 0.98 | 0.35 | 0.07 | 0.4017 | 0.58 | 0.41 | 0.61 | 0.29 | 0.49 | 0.40 | 0.58 | 0.28 | -0.06 |
| LIPOPROTEIN SUBCLASSES | Total lipids in chylomicrons and extremely large VLDL | XXL-VLDL-L | mmol/l | 0.04 | 0.03 | 0.01 | 0.09 | 0.08 | 0.11 | 0.02 | 0.12 | 0.28 | **0.0013** | 0.26 | 0.25 | 0.31 | 0.24 | 0.34 | 0.40 | 0.41 | 0.31 | 0.18 | **0.0272** | 0.22 | 0.25 | 0.27 | 0.23 | 0.24 | 0.34 | 0.33 | 0.32 | 0.06 |
| LIPOPROTEIN SUBCLASSES | Total lipids in IDL | IDL-L | mmol/l | 1.24 | 0.32 | 1.21 | 0.25 | 1.27 | 0.30 | 1.20 | 0.28 | 0.02 | 0.8000 | 2.03 | 0.58 | 2.09 | 0.43 | 1.99 | 0.59 | 2.01 | 0.46 | -0.10 | 0.2255 | 0.78 | 0.51 | 0.85 | 0.39 | 0.64 | 0.59 | 0.68 | 0.40 | -0.21 |
| LIPOPROTEIN SUBCLASSES | Total lipids in large HDL | L-HDL-L | mmol/l | 1.17 | 0.47 | 1.24 | 0.32 | 1.06 | 0.47 | 1.09 | 0.36 | -0.23 | **0.0077** | 1.36 | 0.54 | 1.38 | 0.42 | 1.28 | 0.42 | 1.31 | 0.43 | -0.09 | 0.2599 | 0.21 | 0.46 | 0.20 | 0.35 | 0.18 | 0.51 | 0.20 | 0.32 | -0.03 |
| LIPOPROTEIN SUBCLASSES | Total lipids in large LDL | L-LDL-L | mmol/l | 1.53 | 0.47 | 1.52 | 0.31 | 1.61 | 0.42 | 1.54 | 0.35 | 0.12 | 0.1783 | 2.49 | 0.61 | 2.52 | 0.55 | 2.34 | 0.69 | 2.44 | 0.55 | -0.09 | 0.2661 | 0.96 | 0.61 | 0.99 | 0.49 | 0.83 | 0.64 | 0.76 | 0.49 | -0.23 |
| LIPOPROTEIN SUBCLASSES | Total lipids in large VLDL | L-VLDL-L | mmol/l | 0.22 | 0.17 | 0.19 | 0.15 | 0.29 | 0.24 | 0.26 | 0.18 | 0.27 | **0.0022** | 0.57 | 0.39 | 0.62 | 0.32 | 0.65 | 0.55 | 0.70 | 0.40 | 0.10 | 0.2056 | 0.36 | 0.36 | 0.41 | 0.30 | 0.31 | 0.39 | 0.41 | 0.34 | -0.05 |
| LIPOPROTEIN SUBCLASSES | Total lipids in medium HDL | M-HDL-L | mmol/l | 1.36 | 0.25 | 1.36 | 0.18 | 1.32 | 0.29 | 1.32 | 0.21 | -0.09 | 0.3252 | 1.45 | 0.36 | 1.45 | 0.30 | 1.42 | 0.37 | 1.43 | 0.26 | -0.05 | 0.5344 | 0.09 | 0.34 | 0.10 | 0.28 | 0.09 | 0.31 | 0.06 | 0.22 | -0.06 |
| LIPOPROTEIN SUBCLASSES | Total lipids in medium LDL | M-LDL-L | mmol/l | 0.53 | 0.20 | 0.50 | 0.14 | 0.58 | 0.16 | 0.56 | 0.15 | 0.21 | **0.0144** | 0.96 | 0.28 | 0.97 | 0.26 | 0.92 | 0.33 | 0.96 | 0.26 | -0.02 | 0.7724 | 0.43 | 0.30 | 0.45 | 0.22 | 0.39 | 0.27 | 0.37 | 0.21 | -0.18 |
| LIPOPROTEIN SUBCLASSES | Total lipids in medium VLDL | M-VLDL-L | mmol/l | 0.44 | 0.21 | 0.40 | 0.17 | 0.51 | 0.23 | 0.50 | 0.20 | 0.23 | **0.0095** | 1.02 | 0.47 | 1.06 | 0.38 | 1.02 | 0.61 | 1.10 | 0.43 | 0.03 | 0.7079 | 0.62 | 0.48 | 0.63 | 0.34 | 0.50 | 0.55 | 0.57 | 0.34 | -0.11 |

| | | | | 1ST TRIMESTER (N=185) | | | | | | | | | | DELIVERY (N=201) | | | | | | | | | | CHANGE BETWEEN DELIVERY AND 1ST TRIMESTER (DELTA) (N=150) | | | | | | | | |
|---|---|---|---|---|---|---|---|---|---|---|---|---|---|---|---|---|---|---|---|---|---|---|---|---|---|---|---|---|---|---|---|---|
| | | | | CONTROL (N=115) | | | | SSRI (N=70) | | | | | | CONTROL (N=112) | | | | SSRI (N=89) | | | | | | CONTROL (N=110) | | | | SSRI (N=40) | | | | |
| GROUP | VARIABLE | ABBRE-VIATION | UNIT | MEDIAN | IQR | MEAN | SD | MEDIAN | IQR | MEAN | SD | r | p | MEDIAN | IQR | MEAN | SD | MEDIAN | IQR | MEAN | SD | r | p | MEDIAN | IQR | MEAN | SD | MEDIAN | IQR | MEAN | SD | r |
| LIPOPROTEIN SUBCLASSES | Total lipids in small HDL | S-HDL-L | mmol/l | 1.22 | 0.16 | 1.22 | 0.12 | 1.25 | 0.21 | 1.23 | 0.16 | 0.09 | 0.2982 | 1.36 | 0.25 | 1.39 | 0.18 | 1.39 | 0.23 | 1.40 | 0.18 | 0.05 | 0.5248 | 0.14 | 0.24 | 0.17 | 0.17 | 0.13 | 0.23 | 0.12 | 0.18 | -0.10 |
| LIPOPROTEIN SUBCLASSES | Total lipids in small LDL | S-LDL-L | mmol/l | 0.24 | 0.08 | 0.23 | 0.05 | 0.26 | 0.06 | 0.26 | 0.06 | 0.18 | **0.0381** | 0.41 | 0.13 | 0.43 | 0.11 | 0.41 | 0.14 | 0.42 | 0.10 | -0.01 | 0.8653 | 0.17 | 0.12 | 0.18 | 0.09 | 0.16 | 0.10 | 0.16 | 0.08 | -0.15 |
| LIPOPROTEIN SUBCLASSES | Total lipids in small VLDL | S-VLDL-L | mmol/l | 0.35 | 0.15 | 0.33 | 0.11 | 0.39 | 0.15 | 0.38 | 0.12 | 0.19 | **0.0300** | 0.85 | 0.35 | 0.88 | 0.27 | 0.86 | 0.39 | 0.90 | 0.30 | 0.04 | 0.6630 | 0.50 | 0.32 | 0.53 | 0.25 | 0.44 | 0.36 | 0.50 | 0.24 | -0.08 |
| LIPOPROTEIN SUBCLASSES | Total lipids in very large HDL | XL-HDL-L | mmol/l | 0.27 | 0.13 | 0.27 | 0.09 | 0.24 | 0.12 | 0.24 | 0.10 | -0.21 | **0.0147** | 0.36 | 0.15 | 0.36 | 0.10 | 0.33 | 0.13 | 0.34 | 0.12 | -0.10 | 0.2369 | 0.08 | 0.11 | 0.09 | 0.09 | 0.09 | 0.13 | 0.09 | 0.09 | 0.01 |
| LIPOPROTEIN SUBCLASSES | Total lipids in very large VLDL | XL-VLDL-L | mmol/l | 0.10 | 0.11 | 0.08 | 0.10 | 0.15 | 0.14 | 0.13 | 0.12 | 0.29 | **0.0011** | 0.35 | 0.27 | 0.38 | 0.21 | 0.39 | 0.34 | 0.45 | 0.26 | 0.14 | 0.0815 | 0.26 | 0.21 | 0.28 | 0.20 | 0.23 | 0.28 | 0.29 | 0.23 | -0.01 |
| LIPOPROTEIN SUBCLASSES | Total lipids in very small VLDL | XS-VLDL-L | mmol/l | 0.34 | 0.10 | 0.33 | 0.09 | 0.36 | 0.13 | 0.34 | 0.09 | 0.07 | 0.4141 | 0.76 | 0.23 | 0.76 | 0.19 | 0.73 | 0.27 | 0.77 | 0.20 | 0.00 | 0.9893 | 0.41 | 0.20 | 0.42 | 0.17 | 0.36 | 0.20 | 0.39 | 0.16 | -0.12 |
| LIPOPROTEIN SUBCLASSES | Total lipids in VLDL | VLDL-L | mmol/l | 1.49 | 0.73 | 1.35 | 0.63 | 1.78 | 0.93 | 1.67 | 0.75 | 0.25 | **0.0050** | 3.96 | 1.91 | 4.01 | 1.47 | 3.97 | 2.40 | 4.33 | 1.71 | 0.09 | 0.2682 | 2.40 | 1.80 | 2.54 | 1.34 | 2.05 | 2.00 | 2.49 | 1.40 | -0.04 |
| LIPOPROTEIN SUBCLASSES | Total phospholipids in lipoprotein particles | Total-PL | mmol/l | 3.19 | 0.55 | 3.18 | 0.40 | 3.22 | 0.69 | 3.16 | 0.49 | 0.03 | 0.7717 | 4.64 | 0.86 | 4.61 | 0.65 | 4.51 | 0.99 | 4.58 | 0.69 | -0.05 | 0.5216 | 1.37 | 0.85 | 1.42 | 0.63 | 1.16 | 0.99 | 1.22 | 0.60 | -0.16 |
| LIPOPROTEIN SUBCLASSES | Triglycerides in chylomicrons and extremely large VLDL | XXL-VLDL-TG | mmol/l | 0.03 | 0.02 | 0.00 | 0.06 | 0.05 | 0.07 | 0.01 | 0.08 | 0.30 | **0.0010** | 0.13 | 0.18 | 0.17 | 0.17 | 0.19 | 0.28 | 0.24 | 0.22 | 0.18 | **0.0247** | 0.12 | 0.16 | 0.15 | 0.16 | 0.14 | 0.26 | 0.20 | 0.22 | 0.08 |
| LIPOPROTEIN SUBCLASSES | Triglycerides in IDL | IDL-TG | mmol/l | 0.12 | 0.03 | 0.11 | 0.03 | 0.12 | 0.03 | 0.12 | 0.03 | 0.10 | 0.2395 | 0.25 | 0.08 | 0.26 | 0.06 | 0.26 | 0.09 | 0.27 | 0.07 | 0.07 | 0.4045 | 0.14 | 0.07 | 0.14 | 0.06 | 0.13 | 0.07 | 0.14 | 0.06 | -0.01 |
| LIPOPROTEIN SUBCLASSES | Triglycerides in large HDL | L-HDL-TG | mmol/l | 0.05 | 0.02 | 0.05 | 0.02 | 0.05 | 0.02 | 0.05 | 0.02 | 0.02 | 0.8308 | 0.10 | 0.04 | 0.10 | 0.03 | 0.10 | 0.03 | 0.11 | 0.03 | 0.05 | 0.5621 | 0.05 | 0.03 | 0.05 | 0.02 | 0.05 | 0.03 | 0.05 | 0.02 | -0.01 |
| LIPOPROTEIN SUBCLASSES | Triglycerides in large VLDL | L-VLDL-TG | mmol/l | 0.12 | 0.10 | 0.10 | 0.09 | 0.16 | 0.13 | 0.14 | 0.10 | 0.27 | **0.0019** | 0.26 | 0.22 | 0.30 | 0.18 | 0.30 | 0.30 | 0.34 | 0.23 | 0.10 | 0.2056 | 0.15 | 0.19 | 0.18 | 0.17 | 0.14 | 0.22 | 0.19 | 0.20 | -0.03 |
| LIPOPROTEIN SUBCLASSES | Triglycerides in LDL | LDL-TG | mmol/l | 0.16 | 0.04 | 0.16 | 0.04 | 0.17 | 0.05 | 0.17 | 0.04 | 0.12 | 0.1720 | 0.37 | 0.12 | 0.38 | 0.09 | 0.37 | 0.14 | 0.39 | 0.10 | 0.09 | 0.2978 | 0.21 | 0.10 | 0.21 | 0.08 | 0.20 | 0.10 | 0.21 | 0.09 | 0.00 |
| LIPOPROTEIN SUBCLASSES | Triglycerides in medium HDL | M-HDL-TG | mmol/l | 0.06 | 0.02 | 0.06 | 0.02 | 0.07 | 0.03 | 0.07 | 0.02 | 0.15 | 0.0844 | 0.13 | 0.04 | 0.13 | 0.03 | 0.13 | 0.04 | 0.13 | 0.03 | 0.11 | 0.1727 | 0.07 | 0.04 | 0.06 | 0.03 | 0.06 | 0.03 | 0.06 | 0.03 | -0.05 |
| LIPOPROTEIN SUBCLASSES | Triglycerides in medium LDL | M-LDL-TG | mmol/l | 0.04 | 0.01 | 0.04 | 0.01 | 0.04 | 0.01 | 0.04 | 0.01 | 0.15 | 0.0951 | 0.09 | 0.03 | 0.09 | 0.02 | 0.08 | 0.04 | 0.09 | 0.03 | 0.09 | 0.2746 | 0.05 | 0.03 | 0.05 | 0.02 | 0.05 | 0.03 | 0.05 | 0.02 | 0.02 |
| LIPOPROTEIN SUBCLASSES | Triglycerides in medium VLDL | M-VLDL-TG | mmol/l | 0.23 | 0.14 | 0.21 | 0.11 | 0.28 | 0.16 | 0.26 | 0.12 | 0.24 | **0.0066** | 0.50 | 0.28 | 0.54 | 0.22 | 0.55 | 0.36 | 0.58 | 0.28 | 0.09 | 0.2630 | 0.29 | 0.26 | 0.31 | 0.21 | 0.24 | 0.29 | 0.30 | 0.23 | -0.05 |
| LIPOPROTEIN SUBCLASSES | Triglycerides in small HDL | S-HDL-TG | mmol/l | 0.05 | 0.02 | 0.05 | 0.02 | 0.06 | 0.02 | 0.05 | 0.02 | 0.21 | **0.0153** | 0.11 | 0.04 | 0.11 | 0.03 | 0.11 | 0.04 | 0.12 | 0.03 | 0.14 | 0.0828 | 0.06 | 0.04 | 0.06 | 0.03 | 0.06 | 0.04 | 0.06 | 0.03 | 0.00 |
| LIPOPROTEIN SUBCLASSES | Triglycerides in small LDL | S-LDL-TG | mmol/l | 0.01 | 0.00 | 0.01 | 0.00 | 0.02 | 0.01 | 0.02 | 0.00 | 0.21 | **0.0166** | 0.04 | 0.01 | 0.04 | 0.01 | 0.04 | 0.01 | 0.04 | 0.01 | 0.14 | 0.1006 | 0.02 | 0.01 | 0.02 | 0.01 | 0.02 | 0.01 | 0.02 | 0.01 | 0.05 |
| LIPOPROTEIN SUBCLASSES | Triglycerides in small VLDL | S-VLDL-TG | mmol/l | 0.16 | 0.07 | 0.15 | 0.06 | 0.18 | 0.07 | 0.17 | 0.06 | 0.18 | **0.0403** | 0.37 | 0.17 | 0.38 | 0.12 | 0.40 | 0.20 | 0.41 | 0.15 | 0.10 | 0.2418 | 0.22 | 0.15 | 0.22 | 0.11 | 0.21 | 0.16 | 0.23 | 0.13 | -0.01 |
| LIPOPROTEIN SUBCLASSES | Triglycerides in very large HDL | XL-HDL-TG | mmol/l | 0.01 | 0.00 | 0.01 | 0.00 | 0.01 | 0.00 | 0.01 | 0.00 | 0.03 | 0.6992 | 0.02 | 0.01 | 0.02 | 0.01 | 0.02 | 0.01 | 0.02 | 0.01 | 0.09 | 0.3001 | 0.01 | 0.01 | 0.01 | 0.01 | 0.01 | 0.01 | 0.01 | 0.01 | 0.05 |
| LIPOPROTEIN SUBCLASSES | Triglycerides in very large VLDL | XL-VLDL-TG | mmol/l | 0.06 | 0.07 | 0.04 | 0.06 | 0.09 | 0.09 | 0.07 | 0.07 | 0.29 | **0.0010** | 0.19 | 0.15 | 0.21 | 0.13 | 0.23 | 0.21 | 0.26 | 0.17 | 0.16 | 0.0510 | 0.13 | 0.12 | 0.15 | 0.12 | 0.14 | 0.20 | 0.17 | 0.15 | 0.01 |
| LIPOPROTEIN SUBCLASSES | Triglycerides in very small VLDL | XS-VLDL-TG | mmol/l | 0.08 | 0.02 | 0.07 | 0.02 | 0.08 | 0.03 | 0.08 | 0.02 | 0.13 | 0.1406 | 0.18 | 0.06 | 0.19 | 0.05 | 0.19 | 0.09 | 0.20 | 0.06 | 0.08 | 0.3568 | 0.11 | 0.06 | 0.11 | 0.05 | 0.10 | 0.07 | 0.11 | 0.05 | 0.00 |
| LIPOPROTEIN SUBCLASSES ( LARGE HDL) | Cholesterol in large HDL | L-HDL-C | mmol/l | 0.55 | 0.23 | 0.59 | 0.16 | 0.49 | 0.26 | 0.50 | 0.19 | -0.24 | **0.0054** | 0.61 | 0.28 | 0.62 | 0.21 | 0.57 | 0.25 | 0.58 | 0.22 | -0.11 | 0.1705 | 0.07 | 0.23 | 0.07 | 0.17 | 0.06 | 0.27 | 0.07 | 0.16 | -0.03 |
| LIPOPROTEIN SUBCLASSES (CHYLOMICRONS AND EXTREMELY LARGE LDL) | Cholesterol in chylomicrons and extremely large VLDL | XXL-VLDL-C | mmol/l | 0.01 | 0.01 | 0.01 | 0.02 | 0.02 | 0.03 | 0.01 | 0.03 | 0.29 | **0.0010** | 0.08 | 0.06 | 0.09 | 0.05 | 0.09 | 0.07 | 0.10 | 0.06 | 0.15 | 0.0749 | 0.07 | 0.05 | 0.07 | 0.04 | 0.06 | 0.06 | 0.08 | 0.05 | 0.03 |
| LIPOPROTEIN SUBCLASSES (IDL) | Cholesterol in IDL | IDL-C | mmol/l | 0.84 | 0.23 | 0.82 | 0.18 | 0.86 | 0.20 | 0.82 | 0.20 | 0.01 | 0.8908 | 1.31 | 0.36 | 1.36 | 0.29 | 1.27 | 0.39 | 1.29 | 0.31 | -0.12 | 0.1311 | 0.48 | 0.35 | 0.51 | 0.27 | 0.41 | 0.40 | 0.39 | 0.28 | -0.21 |
| LIPOPROTEIN SUBCLASSES (LARGE LDL) | Cholesterol in large LDL | L-LDL-C | mmol/l | 1.09 | 0.35 | 1.08 | 0.23 | 1.15 | 0.30 | 1.10 | 0.26 | 0.12 | 0.1702 | 1.72 | 0.46 | 1.75 | 0.39 | 1.63 | 0.48 | 1.67 | 0.39 | -0.11 | 0.1735 | 0.63 | 0.47 | 0.66 | 0.35 | 0.50 | 0.50 | 0.47 | 0.36 | -0.25 |
| LIPOPROTEIN SUBCLASSES (LARGE VLDL) | Cholesterol in large VLDL | L-VLDL-C | mmol/l | 0.06 | 0.05 | 0.05 | 0.04 | 0.07 | 0.06 | 0.07 | 0.04 | 0.25 | **0.0050** | 0.17 | 0.10 | 0.18 | 0.08 | 0.19 | 0.13 | 0.20 | 0.10 | 0.08 | 0.3593 | 0.12 | 0.10 | 0.13 | 0.08 | 0.09 | 0.10 | 0.12 | 0.08 | -0.06 |
| LIPOPROTEIN SUBCLASSES (MEDIUM HDL) | Cholesterol in medium HDL | M-HDL-C | mmol/l | 0.67 | 0.14 | 0.68 | 0.09 | 0.64 | 0.14 | 0.65 | 0.11 | -0.15 | 0.0814 | 0.67 | 0.18 | 0.66 | 0.16 | 0.61 | 0.24 | 0.63 | 0.15 | -0.10 | 0.2190 | -0.03 | 0.17 | -0.01 | 0.15 | -0.02 | 0.18 | -0.03 | 0.12 | -0.06 |
| LIPOPROTEIN SUBCLASSES (MEDIUM HDL) | Cholesteryl esters in medium HDL | M-HDL-CE | mmol/l | 0.55 | 0.11 | 0.55 | 0.07 | 0.52 | 0.11 | 0.53 | 0.08 | -0.16 | 0.0623 | 0.53 | 0.15 | 0.51 | 0.13 | 0.48 | 0.19 | 0.49 | 0.12 | -0.10 | 0.2021 | -0.04 | 0.14 | -0.03 | 0.12 | -0.03 | 0.13 | -0.05 | 0.10 | -0.06 |
| LIPOPROTEIN SUBCLASSES (MEDIUM HDL) | Cholesteryl esters in medium LDL | M-LDL-CE | mmol/l | 0.24 | 0.10 | 0.23 | 0.07 | 0.27 | 0.08 | 0.27 | 0.08 | 0.24 | **0.0064** | 0.47 | 0.16 | 0.47 | 0.13 | 0.46 | 0.17 | 0.47 | 0.13 | -0.01 | 0.8730 | 0.21 | 0.15 | 0.23 | 0.11 | 0.19 | 0.14 | 0.19 | 0.11 | -0.18 |
| LIPOPROTEIN SUBCLASSES (MEDIUM HDL) | Cholesteryl esters in medium VLDL | M-VLDL-CE | mmol/l | 0.06 | 0.03 | 0.06 | 0.03 | 0.06 | 0.03 | 0.06 | 0.03 | 0.07 | 0.4109 | 0.13 | 0.07 | 0.14 | 0.06 | 0.12 | 0.07 | 0.13 | 0.05 | -0.12 | 0.1460 | 0.07 | 0.06 | 0.08 | 0.05 | 0.05 | 0.06 | 0.06 | 0.04 | -0.20 |
| LIPOPROTEIN SUBCLASSES (MEDIUM HDL) | Cholesteryl esters in small HDL | S-HDL-CE | mmol/l | 0.33 | 0.05 | 0.33 | 0.04 | 0.34 | 0.06 | 0.33 | 0.05 | 0.06 | 0.4626 | 0.30 | 0.08 | 0.30 | 0.06 | 0.30 | 0.08 | 0.30 | 0.06 | -0.05 | 0.5720 | -0.04 | 0.07 | -0.03 | 0.05 | -0.05 | 0.08 | -0.04 | 0.06 | -0.16 |
| LIPOPROTEIN SUBCLASSES (MEDIUM HDL) | Cholesteryl esters in small LDL | S-LDL-CE | mmol/l | 0.11 | 0.03 | 0.10 | 0.03 | 0.12 | 0.03 | 0.11 | 0.03 | 0.21 | **0.0171** | 0.19 | 0.06 | 0.19 | 0.05 | 0.18 | 0.07 | 0.19 | 0.05 | -0.02 | 0.8100 | 0.08 | 0.05 | 0.09 | 0.04 | 0.08 | 0.05 | 0.07 | 0.04 | -0.17 |
| LIPOPROTEIN SUBCLASSES (MEDIUM HDL) | Cholesteryl esters in small VLDL | S-VLDL-CE | mmol/l | 0.07 | 0.03 | 0.06 | 0.02 | 0.08 | 0.03 | 0.07 | 0.03 | 0.17 | **0.0499** | 0.19 | 0.08 | 0.19 | 0.07 | 0.19 | 0.09 | 0.19 | 0.07 | -0.01 | 0.8787 | 0.12 | 0.07 | 0.13 | 0.06 | 0.11 | 0.09 | 0.11 | 0.05 | -0.12 |
| LIPOPROTEIN SUBCLASSES (MEDIUM HDL) | Cholesteryl esters in very large HDL | XL-HDL-CE | mmol/l | 0.09 | 0.04 | 0.09 | 0.03 | 0.08 | 0.04 | 0.08 | 0.03 | -0.22 | **0.0129** | 0.12 | 0.05 | 0.12 | 0.03 | 0.11 | 0.04 | 0.11 | 0.04 | -0.12 | 0.1361 | 0.02 | 0.03 | 0.02 | 0.03 | 0.02 | 0.04 | 0.02 | 0.03 | -0.01 |
| LIPOPROTEIN SUBCLASSES (MEDIUM HDL) | Cholesteryl esters in very large VLDL | XL-VLDL-CE | mmol/l | 0.01 | 0.01 | 0.01 | 0.01 | 0.02 | 0.01 | 0.02 | 0.01 | 0.25 | **0.0048** | 0.05 | 0.03 | 0.05 | 0.02 | 0.05 | 0.04 | 0.06 | 0.03 | 0.06 | 0.4441 | 0.04 | 0.03 | 0.04 | 0.02 | 0.03 | 0.03 | 0.03 | 0.02 | -0.08 |
| LIPOPROTEIN SUBCLASSES (MEDIUM HDL) | Cholesteryl esters in very small VLDL | XS-VLDL-CE | mmol/l | 0.11 | 0.03 | 0.11 | 0.03 | 0.12 | 0.04 | 0.11 | 0.03 | 0.04 | 0.6887 | 0.22 | 0.06 | 0.22 | 0.05 | 0.21 | 0.07 | 0.21 | 0.06 | -0.07 | 0.3762 | 0.10 | 0.06 | 0.11 | 0.05 | 0.08 | 0.07 | 0.09 | 0.05 | -0.18 |
| LIPOPROTEIN SUBCLASSES (MEDIUM HDL) | Cholesteryl esters in VLDL | VLDL-CE | mmol/l | 0.29 | 0.14 | 0.28 | 0.10 | 0.33 | 0.13 | 0.31 | 0.11 | 0.17 | 0.0540 | 0.74 | 0.31 | 0.75 | 0.24 | 0.71 | 0.30 | 0.75 | 0.24 | -0.01 | 0.8576 | 0.43 | 0.28 | 0.46 | 0.22 | 0.38 | 0.31 | 0.40 | 0.18 | -0.13 |
| LIPOPROTEIN SUBCLASSES (MEDIUM LDL) | Cholesterol in medium LDL | M-LDL-C | mmol/l | 0.35 | 0.14 | 0.33 | 0.10 | 0.39 | 0.12 | 0.37 | 0.10 | 0.23 | **0.0100** | 0.62 | 0.19 | 0.64 | 0.18 | 0.62 | 0.20 | 0.62 | 0.17 | -0.03 | 0.6934 | 0.27 | 0.19 | 0.29 | 0.15 | 0.24 | 0.20 | 0.23 | 0.14 | -0.19 |

| | | | | 1ST TRIMESTER (N=185) | | | | | | | | | | DELIVERY (N=201) | | | | | | | | | | CHANGE BETWEEN DELIVERY AND 1ST TRIMESTER (DELTA) (N=150) | | | | | | | | |
|---|---|---|---|---|---|---|---|---|---|---|---|---|---|---|---|---|---|---|---|---|---|---|---|---|---|---|---|---|---|---|---|---|
| | | | | CONTROL (N=115) | | | | SSRI (N=70) | | | | | | CONTROL (N=112) | | | | SSRI (N=89) | | | | | | CONTROL (N=110) | | | | SSRI (N=40) | | | | |
| GROUP | VARIABLE | ABBRE-VIATION | UNIT | MEDIAN | IQR | MEAN | SD | MEDIAN | IQR | MEAN | SD | r | p | MEDIAN | IQR | MEAN | SD | MEDIAN | IQR | MEAN | SD | r | p | MEDIAN | IQR | MEAN | SD | MEDIAN | IQR | MEAN | SD | r |
| LIPOPROTEIN SUBCLASSES (MEDIUM VLDL) | Cholesterol in medium VLDL | M-VLDL-C | mmol/l | 0.11 | 0.07 | 0.11 | 0.05 | 0.13 | 0.05 | 0.12 | 0.05 | 0.14 | 0.1198 | 0.28 | 0.13 | 0.28 | 0.10 | 0.25 | 0.14 | 0.27 | 0.10 | -0.08 | 0.3619 | 0.16 | 0.11 | 0.17 | 0.09 | 0.13 | 0.12 | 0.14 | 0.08 | -0.20 |
| LIPOPROTEIN SUBCLASSES (SMALL HDL) | Cholesterol in small HDL | S-HDL-C | mmol/l | 0.46 | 0.06 | 0.46 | 0.05 | 0.47 | 0.07 | 0.47 | 0.06 | 0.07 | 0.4044 | 0.48 | 0.08 | 0.48 | 0.07 | 0.47 | 0.10 | 0.47 | 0.08 | -0.02 | 0.8119 | 0.00 | 0.09 | 0.01 | 0.06 | -0.01 | 0.10 | -0.01 | 0.08 | -0.13 |
| LIPOPROTEIN SUBCLASSES (SMALL LDL) | Cholesterol in small LDL | S-LDL-C | mmol/l | 0.15 | 0.05 | 0.14 | 0.04 | 0.16 | 0.04 | 0.16 | 0.04 | 0.19 | **0.0320** | 0.25 | 0.08 | 0.26 | 0.07 | 0.24 | 0.08 | 0.25 | 0.07 | -0.04 | 0.6401 | 0.10 | 0.07 | 0.11 | 0.06 | 0.09 | 0.06 | 0.09 | 0.05 | -0.18 |
| LIPOPROTEIN SUBCLASSES (SMALL VLDL) | Cholesterol in small VLDL | S-VLDL-C | mmol/l | 0.11 | 0.05 | 0.10 | 0.04 | 0.13 | 0.05 | 0.12 | 0.04 | 0.17 | 0.0548 | 0.30 | 0.13 | 0.30 | 0.10 | 0.29 | 0.15 | 0.30 | 0.10 | -0.02 | 0.7855 | 0.18 | 0.11 | 0.19 | 0.09 | 0.16 | 0.13 | 0.17 | 0.08 | -0.13 |
| LIPOPROTEIN SUBCLASSES (VERY LARGE HDL) | Cholesterol in very large HDL | XL-HDL-C | mmol/l | 0.12 | 0.06 | 0.12 | 0.04 | 0.11 | 0.05 | 0.11 | 0.04 | -0.22 | **0.0135** | 0.16 | 0.06 | 0.16 | 0.04 | 0.15 | 0.05 | 0.15 | 0.05 | -0.11 | 0.1846 | 0.03 | 0.04 | 0.03 | 0.04 | 0.03 | 0.06 | 0.03 | 0.04 | 0.01 |
| LIPOPROTEIN SUBCLASSES (VERY LARGE VLDL) | Cholesterol in very large VLDL | XL-VLDL-C | mmol/l | 0.02 | 0.02 | 0.02 | 0.02 | 0.04 | 0.03 | 0.03 | 0.02 | 0.27 | **0.0024** | 0.09 | 0.05 | 0.10 | 0.04 | 0.10 | 0.07 | 0.10 | 0.05 | 0.08 | 0.3023 | 0.07 | 0.05 | 0.07 | 0.04 | 0.05 | 0.06 | 0.07 | 0.04 | -0.06 |
| LIPOPROTEIN SUBCLASSES (VERY SMALL VLDL) | Cholesterol in very small VLDL | XS-VLDL-C | mmol/l | 0.17 | 0.05 | 0.17 | 0.04 | 0.17 | 0.05 | 0.16 | 0.04 | 0.05 | 0.5761 | 0.34 | 0.10 | 0.34 | 0.08 | 0.33 | 0.12 | 0.33 | 0.09 | -0.05 | 0.5441 | 0.17 | 0.09 | 0.17 | 0.08 | 0.14 | 0.10 | 0.15 | 0.07 | -0.16 |
| OTHER LIPIDS | Phosphatidylcholines | Phosphatidylc | mmol/l | 2.68 | 0.51 | 2.64 | 0.38 | 2.69 | 0.62 | 2.68 | 0.43 | 0.04 | 0.6804 | 3.85 | 0.70 | 3.84 | 0.55 | 3.78 | 0.80 | 3.84 | 0.59 | -0.02 | 0.7817 | 1.15 | 0.67 | 1.17 | 0.54 | 0.95 | 0.85 | 1.02 | 0.51 | -0.16 |
| OTHER LIPIDS | Phosphoglycerides | Phosphoglyc | mmol/l | 2.72 | 0.49 | 2.68 | 0.38 | 2.75 | 0.63 | 2.73 | 0.44 | 0.05 | 0.5956 | 3.90 | 0.69 | 3.88 | 0.55 | 3.81 | 0.87 | 3.89 | 0.59 | -0.01 | 0.8807 | 1.12 | 0.69 | 1.16 | 0.55 | 0.91 | 0.86 | 1.01 | 0.51 | -0.15 |
| OTHER LIPIDS | Ratio of triglycerides to phosphoglycerides | TG/PG | % | 0.41 | 0.15 | 0.38 | 0.13 | 0.47 | 0.19 | 0.44 | 0.15 | 0.25 | **0.0045** | 0.68 | 0.26 | 0.71 | 0.22 | 0.75 | 0.36 | 0.79 | 0.27 | 0.18 | **0.0251** | 0.28 | 0.22 | 0.31 | 0.20 | 0.30 | 0.36 | 0.33 | 0.24 | 0.03 |
| OTHER LIPIDS | Sphingomyelins | Sphingomyelins | mmol/l | 0.50 | 0.08 | 0.49 | 0.06 | 0.50 | 0.07 | 0.49 | 0.07 | -0.02 | 0.8175 | 0.68 | 0.12 | 0.68 | 0.09 | 0.66 | 0.12 | 0.66 | 0.10 | -0.08 | 0.3392 | 0.17 | 0.12 | 0.18 | 0.09 | 0.14 | 0.15 | 0.15 | 0.09 | -0.17 |
| OTHER LIPIDS | Total cholines | Cholines | mmol/l | 3.00 | 0.52 | 3.00 | 0.37 | 3.02 | 0.58 | 3.00 | 0.44 | 0.03 | 0.7501 | 4.15 | 0.70 | 4.16 | 0.56 | 4.09 | 0.87 | 4.14 | 0.60 | -0.04 | 0.6612 | 1.12 | 0.64 | 1.16 | 0.55 | 0.90 | 0.87 | 0.99 | 0.53 | -0.15 |
| RELATIVE LIPOPROTEIN LIPID CONCENTRATIONS | Free cholesterol to total lipids ratio in chylomicrons and extremely large VLDL[* 5] | XXL-VLDL-FC % | % | | | | | | | | | | | 11.98 | 5.79 | 13.46 | 4.63 | 10.65 | 3.91 | 12.21 | 4.53 | -0.20 | **0.0172** | | | | | | | | | |
| RELATIVE LIPOPROTEIN LIPID CONCENTRATIONS | Free cholesterol to total lipids ratio in IDL | IDL-FC % | % | 17.02 | 0.70 | 17.05 | 0.64 | 16.83 | 1.18 | 16.89 | 0.77 | -0.13 | 0.1460 | 16.33 | 0.65 | 16.23 | 0.56 | 16.03 | 0.71 | 15.94 | 0.87 | -0.23 | **0.0046** | -0.77 | 0.83 | -0.80 | 0.62 | -0.88 | 0.68 | -0.99 | 0.98 | -0.14 |
| RELATIVE LIPOPROTEIN LIPID CONCENTRATIONS | Free cholesterol to total lipids ratio in large HDL | L-HDL-FC % | % | 10.55 | 0.52 | 10.56 | 0.44 | 10.41 | 0.68 | 10.47 | 0.65 | -0.12 | 0.1632 | 11.08 | 0.46 | 11.12 | 0.44 | 11.13 | 0.58 | 11.03 | 0.52 | -0.03 | 0.7079 | 0.51 | 0.57 | 0.56 | 0.46 | 0.52 | 0.65 | 0.52 | 0.60 | -0.01 |
| RELATIVE LIPOPROTEIN LIPID CONCENTRATIONS | Free cholesterol to total lipids ratio in large LDL | L-LDL-FC % | % | 18.81 | 1.19 | 18.86 | 0.83 | 18.53 | 1.05 | 18.53 | 0.87 | -0.20 | **0.0264** | 17.34 | 1.22 | 17.20 | 1.06 | 16.75 | 1.50 | 16.72 | 1.45 | -0.23 | **0.0045** | -1.61 | 1.15 | -1.62 | 0.95 | -1.84 | 1.67 | -1.92 | 1.37 | -0.12 |
| RELATIVE LIPOPROTEIN LIPID CONCENTRATIONS | Free cholesterol to total lipids ratio in large VLDL | L-VLDL-FC % | % | 12.24 | 1.97 | 12.32 | 2.12 | 12.43 | 1.27 | 12.60 | 1.37 | 0.08 | 0.3825 | 14.42 | 1.98 | 14.68 | 1.74 | 13.95 | 2.30 | 14.71 | 2.43 | -0.08 | 0.3270 | 2.02 | 2.59 | 2.40 | 2.57 | 1.42 | 3.16 | 2.33 | 2.88 | -0.09 |
| RELATIVE LIPOPROTEIN LIPID CONCENTRATIONS | Free cholesterol to total lipids ratio in medium HDL | M-HDL-FC % | % | 9.25 | 0.58 | 9.22 | 0.41 | 9.15 | 0.65 | 9.18 | 0.52 | -0.12 | 0.1702 | 9.93 | 0.54 | 9.94 | 0.45 | 9.81 | 0.51 | 9.83 | 0.47 | -0.15 | 0.0718 | 0.70 | 0.54 | 0.69 | 0.38 | 0.51 | 0.63 | 0.61 | 0.48 | -0.17 |
| RELATIVE LIPOPROTEIN LIPID CONCENTRATIONS | Free cholesterol to total lipids ratio in medium LDL | M-LDL-FC % | % | 20.15 | 1.85 | 20.37 | 1.49 | 19.68 | 1.86 | 19.95 | 1.49 | -0.19 | **0.0279** | 17.20 | 2.02 | 17.04 | 1.68 | 16.30 | 2.44 | 16.38 | 1.98 | -0.23 | **0.0046** | -3.07 | 2.19 | -3.16 | 1.61 | -3.50 | 2.36 | -3.39 | 1.83 | -0.07 |
| RELATIVE LIPOPROTEIN LIPID CONCENTRATIONS | Free cholesterol to total lipids ratio in medium VLDL | M-VLDL-FC % | % | 12.50 | 2.41 | 12.52 | 1.85 | 12.39 | 2.17 | 12.13 | 1.43 | -0.06 | 0.4626 | 13.61 | 1.80 | 13.64 | 1.41 | 13.29 | 2.42 | 13.38 | 1.93 | -0.14 | 0.0972 | 0.97 | 2.55 | 1.08 | 1.82 | 0.58 | 2.40 | 0.89 | 1.94 | -0.08 |
| RELATIVE LIPOPROTEIN LIPID CONCENTRATIONS | Free cholesterol to total lipids ratio in small HDL | S-HDL-FC % | % | 10.81 | 0.86 | 10.69 | 0.66 | 10.74 | 0.77 | 10.66 | 0.70 | -0.07 | 0.4421 | 12.51 | 1.38 | 12.63 | 1.03 | 12.44 | 1.30 | 12.57 | 1.04 | -0.03 | 0.6755 | 1.62 | 1.10 | 1.83 | 0.87 | 1.76 | 0.98 | 1.77 | 0.89 | 0.01 |
| RELATIVE LIPOPROTEIN LIPID CONCENTRATIONS | Free cholesterol to total lipids ratio in small LDL | S-LDL-FC % | % | 17.89 | 1.54 | 18.15 | 1.74 | 17.71 | 1.62 | 17.87 | 1.43 | -0.13 | 0.1429 | 15.31 | 1.69 | 15.14 | 1.59 | 14.75 | 2.15 | 14.67 | 1.86 | -0.17 | **0.0340** | -2.81 | 1.96 | -2.79 | 1.58 | -2.91 | 1.81 | -2.98 | 1.66 | -0.06 |
| RELATIVE LIPOPROTEIN LIPID CONCENTRATIONS | Free cholesterol to total lipids ratio in small VLDL | S-VLDL-FC % | % | 12.73 | 2.67 | 12.63 | 1.86 | 12.70 | 2.05 | 12.23 | 1.71 | -0.04 | 0.6233 | 12.53 | 1.86 | 12.51 | 1.39 | 12.11 | 1.73 | 12.03 | 1.50 | -0.22 | **0.0065** | -0.26 | 2.08 | -0.27 | 1.67 | -0.62 | 2.29 | -0.65 | 1.61 | -0.11 |
| RELATIVE LIPOPROTEIN LIPID CONCENTRATIONS | Free cholesterol to total lipids ratio in very large HDL | XL-HDL-FC % | % | 11.41 | 1.18 | 11.01 | 1.57 | 11.90 | 1.81 | 11.43 | 1.78 | 0.20 | **0.0219** | 10.48 | 1.01 | 10.50 | 0.94 | 10.52 | 1.08 | 10.60 | 1.08 | 0.06 | 0.4617 | -0.58 | 1.32 | -0.89 | 1.42 | -0.87 | 1.76 | -1.11 | 1.85 | -0.10 |
| RELATIVE LIPOPROTEIN LIPID CONCENTRATIONS | Free cholesterol to total lipids ratio in very large VLDL[1,5] | XL-VLDL-FC % | % | 10.15 | 2.53 | 9.76 | 2.61 | 9.99 | 1.65 | 9.83 | 1.82 | -0.02 | 0.8285 | 11.59 | 1.38 | 11.71 | 1.56 | 11.10 | 1.68 | 11.55 | 2.35 | -0.18 | **0.0258** | 1.57 | 2.30 | 1.34 | 2.42 | 0.91 | 2.32 | 1.33 | 2.27 | -0.08 |
| RELATIVE LIPOPROTEIN LIPID CONCENTRATIONS | Free cholesterol to total lipids ratio in very small VLDL | XS-VLDL-FC % | % | 15.27 | 0.50 | 15.32 | 0.41 | 15.31 | 0.53 | 15.30 | 0.37 | 0.01 | 0.9379 | 15.72 | 0.35 | 15.71 | 0.29 | 15.72 | 0.41 | 15.68 | 0.34 | -0.04 | 0.5954 | 0.44 | 0.48 | 0.43 | 0.37 | 0.35 | 0.51 | 0.37 | 0.42 | -0.08 |
| RELATIVE LIPOPROTEIN LIPID CONCENTRATIONS | Phospholipids to total lipids ratio in chylomicrons and extremely large VLDL[* 5] | XXL-VLDL-PL % | % | | | | | | | | | | | 16.87 | 5.19 | 17.69 | 4.62 | 15.69 | 4.10 | 16.33 | 4.28 | -0.17 | **0.0451** | | | | | | | | | |
| RELATIVE LIPOPROTEIN LIPID CONCENTRATIONS | Phospholipids to total lipids ratio in IDL | IDL-PL % | % | 22.55 | 1.37 | 22.58 | 0.95 | 22.71 | 1.32 | 22.74 | 0.95 | 0.07 | 0.3965 | 22.69 | 1.09 | 22.61 | 0.81 | 22.62 | 1.23 | 22.51 | 0.91 | -0.05 | 0.5770 | 0.03 | 1.30 | 0.02 | 0.89 | -0.18 | 1.09 | -0.24 | 1.17 | -0.15 |
| RELATIVE LIPOPROTEIN LIPID CONCENTRATIONS | Phospholipids to total lipids ratio in large HDL | L-HDL-PL % | % | 49.22 | 1.73 | 48.80 | 1.58 | 49.79 | 2.79 | 49.14 | 2.05 | 0.14 | 0.1039 | 47.42 | 1.57 | 47.54 | 1.39 | 47.57 | 1.91 | 47.94 | 1.67 | 0.11 | 0.1674 | -1.38 | 1.59 | -1.67 | 1.47 | -1.50 | 2.26 | -1.83 | 1.91 | -0.03 |
| RELATIVE LIPOPROTEIN LIPID CONCENTRATIONS | Phospholipids to total lipids ratio in large LDL | L-LDL-PL % | % | 21.38 | 0.93 | 21.42 | 0.79 | 21.33 | 0.87 | 21.19 | 0.72 | -0.10 | 0.2764 | 20.68 | 0.68 | 20.63 | 0.54 | 20.68 | 0.58 | 20.58 | 0.55 | -0.05 | 0.5425 | -0.84 | 0.90 | -0.75 | 0.69 | -0.66 | 1.08 | -0.66 | 0.73 | 0.09 |
| RELATIVE LIPOPROTEIN LIPID CONCENTRATIONS | Phospholipids to total lipids ratio in large VLDL | L-VLDL-PL % | % | 17.47 | 3.10 | 18.41 | 4.70 | 18.23 | 2.13 | 18.95 | 3.17 | 0.09 | 0.3035 | 22.56 | 2.81 | 22.92 | 3.13 | 22.73 | 3.03 | 23.25 | 3.83 | -0.01 | 0.9000 | 4.10 | 3.49 | 5.48 | 5.11 | 4.20 | 4.69 | 5.22 | 5.14 | -0.07 |
| RELATIVE LIPOPROTEIN LIPID CONCENTRATIONS | Phospholipids to total lipids ratio in medium HDL | M-HDL-PL % | % | 45.78 | 1.04 | 45.69 | 0.78 | 46.09 | 1.07 | 46.14 | 0.87 | 0.21 | **0.0161** | 45.96 | 1.22 | 46.05 | 0.97 | 46.25 | 1.49 | 46.47 | 1.15 | 0.21 | **0.0103** | 0.22 | 1.12 | 0.29 | 0.87 | 0.43 | 1.54 | 0.38 | 1.08 | 0.06 |
| RELATIVE LIPOPROTEIN LIPID CONCENTRATIONS | Phospholipids to total lipids ratio in medium LDL | M-LDL-PL % | % | 26.91 | 1.12 | 27.10 | 0.90 | 26.63 | 1.05 | 26.70 | 0.87 | -0.24 | **0.0057** | 25.79 | 0.77 | 25.70 | 0.64 | 25.74 | 0.77 | 25.58 | 0.66 | -0.13 | 0.1101 | -1.24 | 1.11 | -1.20 | 1.02 | -1.05 | 0.72 | -1.08 | 0.87 | 0.08 |

| GROUP | VARIABLE | ABBRE-VIATION | UNIT | 1ST TRIMESTER (N=185) | | | | | | | | | | DELIVERY (N=201) | | | | | | | | | | CHANGE BETWEEN DELIVERY AND 1ST TRIMESTER (DELTA) (N=150) | | | | | | | | |
|---|---|---|---|---|---|---|---|---|---|---|---|---|---|---|---|---|---|---|---|---|---|---|---|---|---|---|---|---|---|---|---|---|
| | | | | CONTROL (N=115) | | | | SSRI (N=70) | | | | | | CONTROL (N=112) | | | | SSRI (N=89) | | | | | | CONTROL (N=110) | | | | SSRI (N=40) | | | | |
| | | | | MEDIAN | IQR | MEAN | SD | MEDIAN | IQR | MEAN | SD | r | p | MEDIAN | IQR | MEAN | SD | MEDIAN | IQR | MEAN | SD | r | p | MEDIAN | IQR | MEAN | SD | MEDIAN | IQR | MEAN | SD | r |
| RELATIVE LIPOPROTEIN LIPID CONCENTRATIONS | Phospholipids to total lipids ratio in medium VLDL | M-VLDL-PL % | % | 21.26 | 2.71 | 21.26 | 2.29 | 21.12 | 2.54 | 20.79 | 1.63 | -0.08 | 0.3887 | 23.09 | 2.17 | 23.03 | 1.69 | 22.69 | 2.71 | 22.88 | 2.45 | -0.09 | 0.2538 | 1.75 | 3.19 | 1.72 | 2.31 | 1.10 | 2.47 | 1.65 | 2.36 | -0.05 |
| RELATIVE LIPOPROTEIN LIPID CONCENTRATIONS | Phospholipids to total lipids ratio in small HDL | S-HDL-PL % | % | 57.92 | 1.47 | 57.91 | 1.18 | 57.71 | 1.34 | 57.86 | 1.06 | -0.07 | 0.4013 | 57.44 | 2.06 | 57.46 | 1.76 | 57.52 | 1.36 | 57.60 | 1.49 | 0.03 | 0.7575 | -0.24 | 1.46 | -0.43 | 1.55 | -0.32 | 1.44 | -0.24 | 1.28 | 0.01 |
| RELATIVE LIPOPROTEIN LIPID CONCENTRATIONS | Phospholipids to total lipids ratio in small LDL | S-LDL-PL % | % | 32.35 | 1.53 | 32.47 | 1.31 | 31.79 | 1.84 | 31.73 | 1.56 | -0.27 | **0.0018** | 31.12 | 1.11 | 31.02 | 0.93 | 31.00 | 1.38 | 30.99 | 1.26 | -0.03 | 0.7042 | -1.35 | 1.91 | -1.33 | 1.50 | -1.09 | 1.84 | -0.81 | 1.50 | 0.23 |
| RELATIVE LIPOPROTEIN LIPID CONCENTRATIONS | Phospholipids to total lipids ratio in small VLDL | S-VLDL-PL % | % | 22.17 | 2.61 | 22.19 | 1.83 | 22.16 | 2.01 | 21.81 | 1.63 | -0.04 | 0.6333 | 21.96 | 1.85 | 21.89 | 1.36 | 21.52 | 1.56 | 21.49 | 1.45 | -0.20 | **0.0127** | -0.41 | 2.08 | -0.33 | 1.61 | -0.42 | 1.83 | -0.61 | 1.47 | -0.07 |
| RELATIVE LIPOPROTEIN LIPID CONCENTRATIONS | Phospholipids to total lipids ratio in very large HDL | XL-HDL-PL % | % | 49.82 | 2.38 | 50.78 | 3.31 | 48.69 | 3.67 | 49.83 | 3.60 | -0.26 | **0.0032** | 50.03 | 2.30 | 49.57 | 2.31 | 49.35 | 2.70 | 49.11 | 2.59 | -0.12 | 0.1407 | -0.36 | 1.84 | -0.28 | 2.44 | -0.21 | 2.10 | 0.13 | 3.78 | 0.05 |
| RELATIVE LIPOPROTEIN LIPID CONCENTRATIONS | Phospholipids to total lipids ratio in very large VLDL [1,5] | XL-VLDL-PL % | % | 14.67 | 3.95 | 15.58 | 3.68 | 15.11 | 3.33 | 15.90 | 3.54 | 0.11 | 0.2300 | 19.61 | 1.42 | 19.73 | 1.61 | 19.23 | 1.68 | 19.59 | 2.11 | -0.12 | 0.1495 | 4.21 | 3.46 | 4.87 | 3.42 | 3.01 | 3.17 | 3.94 | 3.94 | -0.22 |
| RELATIVE LIPOPROTEIN LIPID CONCENTRATIONS | Phospholipids to total lipids ratio in very small VLDL | XS-VLDL-PL % | % | 28.61 | 1.23 | 28.73 | 1.03 | 28.69 | 1.19 | 28.83 | 1.02 | 0.05 | 0.5761 | 30.54 | 0.94 | 30.51 | 0.76 | 30.66 | 1.09 | 30.62 | 0.81 | 0.11 | 0.1782 | 1.76 | 1.12 | 1.92 | 0.98 | 1.99 | 1.21 | 1.95 | 1.06 | 0.05 |
| RELATIVE LIPOPROTEIN LIPID CONCENTRATIONS | Cholesterol to total lipids ratio in chylomicrons and extremely large VLDL* [5] | XXL-VLDL-C % | % | | | | | | | | | | | 28.99 | 17.77 | 36.57 | 18.27 | 26.17 | 12.17 | 32.33 | 17.52 | -0.19 | **0.0200** | | | | | | | | | |
| RELATIVE LIPOPROTEIN LIPID CONCENTRATIONS | Cholesterol to total lipids ratio in IDL | IDL-C % | % | 68.04 | 2.50 | 68.33 | 1.89 | 67.65 | 2.69 | 67.96 | 2.03 | -0.10 | 0.2487 | 65.01 | 3.23 | 64.88 | 2.36 | 64.10 | 3.28 | 63.90 | 2.87 | -0.22 | **0.0081** | -3.13 | 2.78 | -3.16 | 2.15 | -3.73 | 3.12 | -3.97 | 2.74 | -0.17 |
| RELATIVE LIPOPROTEIN LIPID CONCENTRATIONS | Cholesterol to total lipids ratio in large HDL | L-HDL-C % | % | 46.46 | 2.48 | 46.99 | 2.29 | 45.29 | 3.97 | 46.64 | 3.35 | -0.19 | **0.0331** | 44.99 | 3.68 | 44.59 | 2.66 | 44.32 | 3.83 | 43.45 | 3.89 | -0.18 | **0.0314** | -1.62 | 2.93 | -1.92 | 2.44 | -1.69 | 3.01 | -1.89 | 3.67 | 0.01 |
| RELATIVE LIPOPROTEIN LIPID CONCENTRATIONS | Cholesterol to total lipids ratio in large LDL | L-LDL-C % | % | 71.17 | 1.38 | 71.29 | 1.04 | 71.28 | 1.27 | 71.35 | 1.07 | 0.06 | 0.5195 | 69.51 | 2.17 | 69.24 | 1.69 | 68.79 | 2.25 | 68.50 | 1.92 | -0.23 | **0.0052** | -1.78 | 1.87 | -1.95 | 1.62 | -2.93 | 2.47 | -2.97 | 2.08 | -0.34 |
| RELATIVE LIPOPROTEIN LIPID CONCENTRATIONS | Cholesterol to total lipids ratio in large VLDL | L-VLDL-C % | % | 26.27 | 6.18 | 26.06 | 6.14 | 25.67 | 4.08 | 25.28 | 3.73 | -0.09 | 0.3183 | 30.29 | 6.59 | 30.96 | 5.37 | 29.09 | 6.30 | 30.80 | 7.51 | -0.10 | 0.2227 | 3.50 | 8.89 | 4.54 | 7.38 | 3.31 | 9.07 | 5.27 | 8.78 | 0.00 |
| RELATIVE LIPOPROTEIN LIPID CONCENTRATIONS | Cholesterol to total lipids ratio in medium HDL | M-HDL-C % | % | 49.52 | 2.27 | 49.61 | 1.83 | 48.80 | 2.25 | 48.87 | 2.05 | -0.22 | **0.0111** | 45.29 | 4.06 | 44.82 | 3.50 | 44.24 | 4.98 | 43.85 | 3.54 | -0.18 | **0.0267** | -4.29 | 3.55 | -4.77 | 3.20 | -4.05 | 4.41 | -4.89 | 2.98 | -0.04 |
| RELATIVE LIPOPROTEIN LIPID CONCENTRATIONS | Cholesterol to total lipids ratio in medium LDL | M-LDL-C % | % | 66.03 | 2.09 | 66.21 | 1.79 | 66.55 | 2.32 | 66.67 | 1.73 | 0.17 | **0.0493** | 65.40 | 2.24 | 65.18 | 1.59 | 64.84 | 2.22 | 64.70 | 1.75 | -0.16 | **0.0452** | -0.84 | 2.08 | -0.88 | 1.64 | -2.10 | 2.24 | -1.75 | 1.98 | -0.32 |
| RELATIVE LIPOPROTEIN LIPID CONCENTRATIONS | Cholesterol to total lipids ratio in medium VLDL | M-VLDL-C % | % | 26.30 | 9.21 | 26.90 | 6.62 | 25.32 | 7.93 | 24.53 | 5.78 | -0.10 | 0.2395 | 26.91 | 7.45 | 26.98 | 5.45 | 25.21 | 7.73 | 25.72 | 6.83 | -0.15 | 0.0637 | 0.54 | 8.64 | 0.47 | 6.59 | -1.02 | 8.13 | 0.26 | 6.61 | -0.03 |
| RELATIVE LIPOPROTEIN LIPID CONCENTRATIONS | Cholesterol to total lipids ratio in small HDL | S-HDL-C % | % | 37.94 | 2.06 | 38.25 | 1.86 | 37.84 | 1.97 | 37.87 | 1.58 | -0.11 | 0.2113 | 34.66 | 3.10 | 34.50 | 2.12 | 34.14 | 3.30 | 33.83 | 2.31 | -0.17 | **0.0419** | -3.44 | 2.80 | -3.52 | 1.91 | -3.63 | 2.86 | -3.88 | 2.25 | -0.06 |
| RELATIVE LIPOPROTEIN LIPID CONCENTRATIONS | Cholesterol to total lipids ratio in small LDL | S-LDL-C % | % | 61.49 | 2.25 | 61.67 | 1.71 | 62.01 | 2.35 | 62.30 | 1.68 | 0.20 | **0.0196** | 60.68 | 2.69 | 60.37 | 1.92 | 59.75 | 2.69 | 59.68 | 2.22 | -0.20 | **0.0161** | -1.01 | 2.51 | -1.16 | 1.79 | -2.47 | 2.82 | -2.28 | 2.74 | -0.29 |
| RELATIVE LIPOPROTEIN LIPID CONCENTRATIONS | Cholesterol to total lipids ratio in small VLDL | S-VLDL-C % | % | 31.99 | 6.17 | 32.02 | 4.63 | 32.35 | 4.00 | 31.73 | 3.96 | -0.01 | 0.9425 | 34.84 | 4.65 | 34.52 | 3.52 | 33.67 | 5.38 | 33.55 | 4.03 | -0.16 | 0.0501 | 2.24 | 5.38 | 2.42 | 4.33 | 1.71 | 4.84 | 1.38 | 4.34 | -0.11 |
| RELATIVE LIPOPROTEIN LIPID CONCENTRATIONS | Cholesterol to total lipids ratio in very large HDL | XL-HDL-C % | % | 46.39 | 2.23 | 45.81 | 2.96 | 46.75 | 2.99 | 45.99 | 2.75 | 0.08 | 0.3779 | 43.35 | 1.73 | 43.47 | 1.59 | 42.92 | 1.78 | 42.98 | 1.96 | -0.15 | 0.0745 | -2.38 | 2.29 | -2.93 | 2.51 | -2.66 | 3.18 | -3.62 | 3.44 | -0.08 |
| RELATIVE LIPOPROTEIN LIPID CONCENTRATIONS | Cholesterol to total lipids ratio in very large VLDL[1,5] | XL-VLDL-C % | % | 28.04 | 10.99 | 25.80 | 11.48 | 26.19 | 8.62 | 23.37 | 9.90 | -0.11 | 0.2262 | 25.74 | 6.92 | 26.65 | 6.11 | 23.76 | 6.37 | 26.04 | 9.13 | -0.19 | **0.0210** | 0.14 | 11.74 | -2.38 | 11.07 | -0.25 | 8.91 | -0.37 | 8.11 | 0.06 |
| RELATIVE LIPOPROTEIN LIPID CONCENTRATIONS | Cholesterol to total lipids ratio in very small VLDL | XS-VLDL-C % | % | 48.70 | 4.17 | 48.65 | 2.93 | 48.21 | 4.66 | 48.15 | 3.36 | -0.10 | 0.2702 | 44.93 | 3.86 | 44.68 | 3.04 | 43.79 | 3.74 | 43.51 | 3.69 | -0.22 | **0.0088** | -4.31 | 3.96 | -4.09 | 3.15 | -4.83 | 5.14 | -4.83 | 3.55 | -0.12 |
| RELATIVE LIPOPROTEIN LIPID CONCENTRATIONS | Cholesteryl esters to total lipids ratio in chylomicrons and extremely large VLDL*[5] | XXL-VLDL-CE % | % | | | | | | | | | | | 17.73 | 12.31 | 23.12 | 13.95 | 16.13 | 8.69 | 20.13 | 13.35 | -0.18 | **0.0295** | | | | | | | | | |
| RELATIVE LIPOPROTEIN LIPID CONCENTRATIONS | Cholesteryl esters to total lipids ratio in IDL | IDL-CE % | % | 51.02 | 2.32 | 51.12 | 1.71 | 50.82 | 2.55 | 50.94 | 1.83 | -0.04 | 0.6193 | 48.61 | 2.75 | 48.64 | 1.97 | 47.78 | 2.75 | 47.96 | 2.27 | -0.20 | **0.0164** | -2.33 | 2.75 | -2.36 | 1.92 | -2.89 | 2.08 | -2.98 | 2.32 | -0.15 |
| RELATIVE LIPOPROTEIN LIPID CONCENTRATIONS | Cholesteryl esters to total lipids ratio in large HDL | L-HDL-CE % | % | 35.91 | 2.16 | 36.55 | 2.04 | 34.88 | 3.75 | 36.02 | 2.88 | -0.20 | **0.0233** | 33.88 | 3.38 | 33.47 | 2.66 | 33.04 | 4.05 | 32.42 | 3.65 | -0.18 | **0.0274** | -1.97 | 2.58 | -2.48 | 2.46 | -2.13 | 3.03 | -2.41 | 3.37 | 0.03 |
| RELATIVE LIPOPROTEIN LIPID CONCENTRATIONS | Cholesteryl esters to total lipids ratio in large LDL | L-LDL-CE % | % | 52.36 | 1.75 | 52.29 | 1.24 | 52.75 | 1.16 | 52.97 | 1.29 | 0.22 | **0.0117** | 52.06 | 1.43 | 52.04 | 1.15 | 51.93 | 1.77 | 51.78 | 1.43 | -0.07 | 0.4170 | -0.27 | 2.05 | -0.33 | 1.43 | -1.19 | 1.67 | -1.05 | 1.46 | -0.29 |
| RELATIVE LIPOPROTEIN LIPID CONCENTRATIONS | Cholesteryl esters to total lipids ratio in large VLDL | L-VLDL-CE % | % | 14.03 | 4.37 | 13.84 | 4.24 | 13.24 | 2.55 | 12.90 | 2.70 | -0.14 | 0.1158 | 15.87 | 4.71 | 16.28 | 3.68 | 15.05 | 4.24 | 16.09 | 5.14 | -0.11 | 0.1970 | 1.58 | 6.50 | 2.14 | 5.01 | 1.72 | 5.30 | 2.94 | 6.08 | 0.04 |
| RELATIVE LIPOPROTEIN LIPID CONCENTRATIONS | Cholesteryl esters to total lipids ratio in medium HDL | M-HDL-CE % | % | 40.28 | 2.13 | 40.52 | 1.78 | 39.66 | 2.13 | 39.76 | 1.88 | -0.22 | **0.0107** | 35.34 | 3.78 | 34.87 | 3.56 | 34.82 | 5.30 | 34.02 | 3.51 | -0.16 | 0.0549 | -5.13 | 3.58 | -5.46 | 3.28 | -4.94 | 4.29 | -5.49 | 3.05 | -0.03 |
| RELATIVE LIPOPROTEIN LIPID CONCENTRATIONS | Cholesteryl esters to total lipids ratio in medium LDL | M-LDL-CE % | % | 45.88 | 2.77 | 45.95 | 2.18 | 46.87 | 2.83 | 47.04 | 2.41 | 0.27 | **0.0018** | 48.04 | 1.96 | 48.14 | 1.54 | 48.21 | 2.45 | 48.32 | 2.01 | 0.08 | 0.3568 | 2.32 | 2.54 | 2.28 | 2.15 | 1.44 | 2.08 | 1.64 | 2.12 | -0.19 |
| RELATIVE LIPOPROTEIN LIPID CONCENTRATIONS | Cholesteryl esters to total lipids ratio in medium VLDL | M-VLDL-CE % | % | 13.79 | 6.91 | 14.24 | 4.88 | 12.93 | 6.55 | 12.67 | 4.43 | -0.12 | 0.1720 | 13.30 | 5.65 | 13.34 | 4.06 | 11.90 | 5.66 | 12.35 | 4.94 | -0.15 | 0.0606 | -0.50 | 6.15 | -0.62 | 4.86 | -0.99 | 6.13 | -0.64 | 4.76 | -0.01 |
| RELATIVE LIPOPROTEIN LIPID CONCENTRATIONS | Cholesteryl esters to total lipids ratio in small HDL | S-HDL-CE % | % | 27.13 | 2.53 | 27.64 | 2.19 | 27.10 | 2.29 | 27.16 | 1.84 | -0.06 | 0.4678 | 21.94 | 4.01 | 21.87 | 2.70 | 21.41 | 4.09 | 21.26 | 2.99 | -0.11 | 0.1727 | -5.05 | 3.46 | -5.35 | 2.39 | -5.53 | 3.27 | -5.65 | 2.88 | -0.06 |
| RELATIVE LIPOPROTEIN LIPID CONCENTRATIONS | Cholesteryl esters to total lipids ratio in small LDL | S-LDL-CE % | % | 43.60 | 1.66 | 43.58 | 1.45 | 44.30 | 2.15 | 44.28 | 1.89 | 0.27 | **0.0024** | 45.10 | 1.45 | 45.23 | 1.17 | 45.08 | 1.97 | 45.01 | 1.82 | -0.05 | 0.5425 | 1.64 | 2.03 | 1.63 | 1.77 | 1.37 | 3.33 | 0.70 | 2.35 | -0.24 |
| RELATIVE LIPOPROTEIN LIPID CONCENTRATIONS | Cholesteryl esters to total lipids ratio in small VLDL | S-VLDL-CE % | % | 19.26 | 4.13 | 19.57 | 2.98 | 19.66 | 3.54 | 19.23 | 2.43 | 0.02 | 0.7956 | 22.25 | 3.10 | 22.01 | 2.34 | 21.72 | 3.42 | 21.52 | 2.73 | -0.10 | 0.2136 | 2.52 | 3.17 | 2.69 | 2.86 | 2.27 | 3.74 | 2.03 | 2.97 | -0.09 |

| | | | | 1ST TRIMESTER (N=185) | | | | | | | | | | DELIVERY (N=201) | | | | | | | | | | CHANGE BETWEEN DELIVERY AND 1ST TRIMESTER (DELTA) (N=150) | | | | | | | | |
|---|---|---|---|---|---|---|---|---|---|---|---|---|---|---|---|---|---|---|---|---|---|---|---|---|---|---|---|---|---|---|---|---|
| | | | | CONTROL (N=115) | | | | SSRI (N=70) | | | | | | CONTROL (N=112) | | | | SSRI (N=89) | | | | | | CONTROL (N=110) | | | | SSRI (N=40) | | | | |
| GROUP | VARIABLE | ABBRE-VIATION | UNIT | MEDIAN | IQR | MEAN | SD | MEDIAN | IQR | MEAN | SD | r | p | MEDIAN | IQR | MEAN | SD | MEDIAN | IQR | MEAN | SD | r | p | MEDIAN | IQR | MEAN | SD | MEDIAN | IQR | MEAN | SD | r |
| RELATIVE LIPOPROTEIN LIPID CONCENTRATIONS | Cholesteryl esters to total lipids ratio in very large HDL | XL-HDL-CE % | % | 34.98 | 1.89 | 34.90 | 1.83 | 34.85 | 1.95 | 34.80 | 1.66 | -0.04 | 0.6598 | 32.85 | 1.80 | 32.97 | 1.44 | 32.43 | 2.20 | 32.38 | 1.81 | -0.19 | **0.0202** | -1.82 | 1.85 | -2.04 | 1.68 | -1.93 | 2.54 | -2.51 | 2.38 | -0.08 |
| RELATIVE LIPOPROTEIN LIPID CONCENTRATIONS | Cholesteryl esters to total lipids ratio in very large VLDL [5] | XL-VLDL-CE % | % | 17.89 | 9.28 | 15.56 | 9.37 | 16.20 | 7.68 | 14.20 | 8.76 | -0.13 | 0.1423 | 14.13 | 6.09 | 14.94 | 4.62 | 12.72 | 4.65 | 14.48 | 6.86 | -0.18 | **0.0253** | -1.24 | 9.92 | -3.72 | 9.14 | -0.61 | 7.45 | -1.70 | 6.92 | 0.11 |
| RELATIVE LIPOPROTEIN LIPID CONCENTRATIONS | Cholesteryl esters to total lipids ratio in very small VLDL | XS-VLDL-CE % | % | 33.42 | 3.81 | 33.46 | 2.69 | 32.90 | 4.18 | 32.90 | 3.15 | -0.10 | 0.2361 | 29.23 | 3.59 | 28.97 | 2.93 | 28.15 | 3.93 | 27.83 | 3.55 | -0.22 | **0.0083** | -4.84 | 3.78 | -4.52 | 2.96 | -5.03 | 4.72 | -5.21 | 3.34 | -0.12 |
| RELATIVE LIPOPROTEIN LIPID CONCENTRATIONS (LARGE HDL RATIOS) | Triglycerides to total lipids ratio in chylomicrons and extremely large VLDL[*5] | XXL-VLDL-TG % | % | | | | | | | | | | | 53.96 | 22.93 | 45.74 | 20.90 | 57.68 | 16.20 | 51.34 | 19.05 | 0.18 | **0.0260** | | | | | | | | | |
| RELATIVE LIPOPROTEIN LIPID CONCENTRATIONS (LARGE HDL RATIOS) | Triglycerides to total lipids ratio in IDL | IDL-TG % | % | 9.41 | 1.98 | 9.16 | 1.51 | 9.63 | 2.47 | 9.59 | 1.61 | 0.09 | 0.2995 | 11.95 | 2.84 | 12.52 | 2.25 | 13.30 | 3.22 | 13.59 | 2.99 | 0.24 | **0.0030** | 2.90 | 2.19 | 3.14 | 2.00 | 3.88 | 3.44 | 4.21 | 3.12 | 0.22 |
| RELATIVE LIPOPROTEIN LIPID CONCENTRATIONS (LARGE HDL RATIOS) | Triglycerides to total lipids ratio in large HDL | L-HDL-TG % | % | 4.32 | 1.40 | 4.09 | 1.20 | 4.92 | 2.01 | 4.59 | 1.66 | 0.21 | **0.0150** | 7.39 | 2.71 | 7.87 | 2.46 | 8.10 | 2.91 | 8.62 | 2.95 | 0.18 | **0.0301** | 3.17 | 2.32 | 3.59 | 2.32 | 2.72 | 2.73 | 3.72 | 2.57 | 0.00 |
| RELATIVE LIPOPROTEIN LIPID CONCENTRATIONS (LARGE HDL RATIOS) | Triglycerides to total lipids ratio in large LDL | L-LDL-TG % | % | 7.46 | 1.33 | 7.14 | 1.29 | 7.39 | 1.42 | 7.43 | 1.15 | 0.01 | 0.9110 | 9.96 | 2.33 | 10.13 | 1.89 | 10.77 | 2.48 | 10.92 | 2.19 | 0.23 | **0.0045** | 2.61 | 1.79 | 2.71 | 1.63 | 3.65 | 2.59 | 3.63 | 2.24 | 0.30 |
| RELATIVE LIPOPROTEIN LIPID CONCENTRATIONS (LARGE HDL RATIOS) | Triglycerides to total lipids ratio in large VLDL | L-VLDL-TG % | % | 56.25 | 8.00 | 56.37 | 9.84 | 56.10 | 5.32 | 55.86 | 5.99 | 0.01 | 0.9334 | 46.72 | 8.89 | 46.12 | 7.63 | 48.61 | 9.57 | 45.95 | 10.83 | 0.06 | 0.4368 | -7.81 | 12.29 | -10.02 | 11.15 | -6.46 | 13.62 | -10.49 | 13.41 | 0.04 |
| RELATIVE LIPOPROTEIN LIPID CONCENTRATIONS (LARGE HDL RATIOS) | Triglycerides to total lipids ratio in medium HDL | M-HDL-TG % | % | 4.69 | 1.47 | 4.54 | 1.28 | 5.11 | 1.45 | 5.05 | 1.35 | 0.21 | **0.0193** | 8.71 | 3.00 | 9.13 | 2.77 | 9.22 | 3.63 | 9.68 | 2.69 | 0.15 | 0.0730 | 4.06 | 2.89 | 4.48 | 2.56 | 4.28 | 2.95 | 4.50 | 2.29 | 0.05 |
| RELATIVE LIPOPROTEIN LIPID CONCENTRATIONS (LARGE HDL RATIOS) | Triglycerides to total lipids ratio in medium LDL | M-LDL-TG % | % | 7.07 | 1.60 | 6.70 | 1.73 | 6.82 | 1.87 | 6.75 | 1.34 | -0.05 | 0.5838 | 8.86 | 2.29 | 9.12 | 1.70 | 9.65 | 2.20 | 9.73 | 1.92 | 0.21 | **0.0122** | 1.93 | 1.83 | 2.08 | 1.49 | 2.87 | 2.45 | 2.84 | 1.85 | 0.28 |
| RELATIVE LIPOPROTEIN LIPID CONCENTRATIONS (LARGE HDL RATIOS) | Triglycerides to total lipids ratio in medium VLDL | M-VLDL-TG % | % | 52.44 | 12.06 | 51.61 | 8.75 | 53.56 | 11.44 | 54.54 | 7.27 | 0.10 | 0.2641 | 50.03 | 9.30 | 49.99 | 7.10 | 51.97 | 10.37 | 51.39 | 9.20 | 0.14 | 0.0942 | -1.97 | 11.88 | -2.19 | 8.76 | -0.03 | 10.15 | -1.91 | 8.84 | 0.03 |
| RELATIVE LIPOPROTEIN LIPID CONCENTRATIONS (LARGE HDL RATIOS) | Triglycerides to total lipids ratio in small HDL | S-HDL-TG % | % | 4.14 | 1.23 | 4.00 | 1.07 | 4.45 | 1.25 | 4.48 | 1.01 | 0.20 | **0.0216** | 7.77 | 2.31 | 8.04 | 2.11 | 8.46 | 3.11 | 8.57 | 2.20 | 0.16 | 0.0521 | 3.75 | 2.26 | 3.94 | 1.87 | 4.19 | 2.26 | 4.11 | 1.89 | 0.09 |
| RELATIVE LIPOPROTEIN LIPID CONCENTRATIONS (LARGE HDL RATIOS) | Triglycerides to total lipids ratio in small LDL | S-LDL-TG % | % | 6.16 | 1.53 | 5.72 | 1.76 | 6.21 | 1.70 | 6.11 | 1.29 | 0.12 | 0.1641 | 8.41 | 2.19 | 8.61 | 1.71 | 9.11 | 2.62 | 9.33 | 1.97 | 0.25 | **0.0028** | 2.54 | 2.00 | 2.49 | 1.60 | 3.09 | 2.23 | 3.09 | 1.80 | 0.18 |
| RELATIVE LIPOPROTEIN LIPID CONCENTRATIONS (LARGE HDL RATIOS) | Triglycerides to total lipids ratio in small VLDL | S-VLDL-TG % | % | 45.84 | 8.68 | 45.97 | 6.36 | 45.49 | 5.21 | 46.52 | 5.51 | 0.02 | 0.8219 | 43.41 | 6.04 | 43.59 | 4.79 | 45.01 | 6.67 | 44.97 | 5.35 | 0.18 | **0.0279** | -1.87 | 7.38 | -2.09 | 5.84 | -1.01 | 6.49 | -0.77 | 5.65 | 0.10 |
| RELATIVE LIPOPROTEIN LIPID CONCENTRATIONS (LARGE HDL RATIOS) | Triglycerides to total lipids ratio in very large HDL | XL-HDL-TG % | % | 3.79 | 1.43 | 3.54 | 1.24 | 4.56 | 2.22 | 4.01 | 1.92 | 0.22 | **0.0119** | 6.74 | 2.69 | 6.97 | 2.17 | 7.49 | 3.00 | 7.92 | 3.45 | 0.19 | **0.0199** | 3.03 | 2.18 | 3.21 | 2.01 | 2.71 | 2.44 | 3.49 | 3.31 | -0.02 |
| RELATIVE LIPOPROTEIN LIPID CONCENTRATIONS (LARGE HDL RATIOS) | Triglycerides to total lipids ratio in very large VLDL[1,5] | XL-VLDL-TG % | % | 57.29 | 10.37 | 58.89 | 12.80 | 58.70 | 8.96 | 60.34 | 9.46 | 0.07 | 0.4567 | 54.48 | 7.63 | 53.63 | 7.51 | 56.71 | 7.48 | 54.37 | 10.94 | 0.18 | **0.0253** | -3.44 | 13.14 | -2.49 | 12.04 | -3.09 | 10.29 | -3.57 | 9.32 | 0.02 |
| RELATIVE LIPOPROTEIN LIPID CONCENTRATIONS (LARGE HDL RATIOS) | Triglycerides to total lipids ratio in very small VLDL | XS-VLDL-TG % | % | 22.70 | 3.50 | 22.39 | 2.75 | 23.10 | 4.75 | 23.43 | 3.06 | 0.10 | 0.2533 | 24.37 | 3.24 | 24.81 | 2.54 | 25.59 | 3.31 | 25.87 | 3.25 | 0.22 | **0.0072** | 2.40 | 3.51 | 2.17 | 2.83 | 2.87 | 4.97 | 2.88 | 3.33 | 0.12 |
| TOTAL LIPIDS | Total lipids in HDL | HDL-L | mmol/l | 4.02 | 0.86 | 4.11 | 0.56 | 3.87 | 0.80 | 3.96 | 0.64 | -0.17 | 0.0548 | 4.58 | 1.05 | 4.58 | 0.84 | 4.47 | 1.05 | 4.48 | 0.80 | -0.06 | 0.4383 | 0.55 | 0.92 | 0.56 | 0.76 | 0.49 | 0.98 | 0.47 | 0.64 | -0.07 |
| TOTAL LIPIDS | Total lipids in LDL | LDL-L | mmol/l | 2.30 | 0.75 | 2.24 | 0.49 | 2.45 | 0.60 | 2.35 | 0.55 | 0.15 | 0.0814 | 3.86 | 1.07 | 3.92 | 0.90 | 3.67 | 0.99 | 3.82 | 0.89 | -0.07 | 0.4100 | 1.60 | 0.97 | 1.62 | 0.78 | 1.45 | 0.88 | 1.28 | 0.76 | -0.22 |
| TOTAL LIPIDS | Total lipids in lipoprotein particles | Total-L | mmol/l | 9.04 | 1.71 | 8.97 | 1.39 | 9.36 | 2.31 | 8.97 | 1.65 | 0.10 | 0.2498 | 14.59 | 3.43 | 14.60 | 2.55 | 14.14 | 4.01 | 14.64 | 2.71 | -0.01 | 0.8710 | 5.34 | 3.04 | 5.57 | 2.38 | 4.83 | 3.10 | 4.91 | 2.23 | -0.16 |
| TRIGLYCERIDES | Total triglycerides | Total-TG | mmol/l | 1.13 | 0.44 | 1.02 | 0.46 | 1.31 | 0.63 | 1.21 | 0.52 | 0.24 | **0.0063** | 2.71 | 1.29 | 2.79 | 0.99 | 2.84 | 1.70 | 3.08 | 1.20 | 0.14 | 0.1001 | 1.58 | 1.30 | 1.68 | 0.92 | 1.46 | 1.50 | 1.73 | 1.03 | 0.00 |
| TRIGLYCERIDES | Triglycerides in HDL | HDL-TG | mmol/l | 0.18 | 0.06 | 0.16 | 0.06 | 0.18 | 0.07 | 0.18 | 0.05 | 0.14 | 0.1198 | 0.37 | 0.11 | 0.36 | 0.08 | 0.38 | 0.12 | 0.38 | 0.09 | 0.10 | 0.2163 | 0.19 | 0.12 | 0.19 | 0.08 | 0.18 | 0.10 | 0.19 | 0.08 | -0.02 |
| TRIGLYCERIDES | Triglycerides in large LDL | L-LDL-TG | mmol/l | 0.11 | 0.03 | 0.11 | 0.03 | 0.12 | 0.03 | 0.11 | 0.03 | 0.09 | 0.2852 | 0.25 | 0.08 | 0.25 | 0.06 | 0.25 | 0.09 | 0.26 | 0.07 | 0.07 | 0.3841 | 0.13 | 0.07 | 0.14 | 0.05 | 0.13 | 0.06 | 0.14 | 0.06 | 0.00 |
| TRIGLYCERIDES | Triglycerides in VLDL | VLDL-TG | mmol/l | 0.67 | 0.38 | 0.57 | 0.37 | 0.84 | 0.55 | 0.74 | 0.43 | 0.25 | **0.0039** | 1.65 | 1.00 | 1.79 | 0.82 | 1.87 | 1.39 | 2.03 | 1.02 | 0.14 | 0.0942 | 1.05 | 0.92 | 1.13 | 0.76 | 0.97 | 1.14 | 1.19 | 0.89 | 0.00 |

p < 0.05 bolded
[*] Values from 1st trimester and delta removed for having 59% missing values
[1] 1st trimester n = 167; [2] 1st trimester n = 184; [3] delivery n = 189; [4] delivery n = 195,[5] delivery n = 200
C , cholesterol; IDL , intermediate-density lipoprotein; IQR , interquartile range; LDL , low-density lipoprotein; SD , standard deviation; SSRI , selective serotonin reuptake inhibitor; VLDL , very low-density lipoprotein

**Table S3**. Linear regression model from 1st trimester timepoint with the NMR analysis variables as the dependent variable, body mass index (BMI) as a predictor and study group (selective serotonin reuptake inhibitor (SSRI)-case vs. control), gestational diabetes mellitus (GDM) and additional medication use as cofactors.

| **1st trimester** | | | | | | | | | | | | |
|---|---|---|---|---|---|---|---|---|---|---|---|---|
| **n = 174 (Control = 95, SSRI = 68)** | | | | | | | | | | | | |
| **Group** | **Variable** | **Overall regression** | | | **SSRI medication** | | **BMI** | | **GDM** | | **Additional medication use** | |
| | | **Adjusted $R^2$** | **F** | **p** | **B** | **p** | **B** | **p** | **B** | **p** | **B** | **p** |
| Lipoprotein subclasses | Cholesteryl esters in chylomicrons and extremely large VLDL (mmol/l) | 0.132 | 7.61 | **< 0.001** | 0.139 | **0.012** | 0.016 | **0.005** | 0.062 | 0.349 | 0.056 | 0.283 |
| Lipoprotein subclasses | Concentration of chylomicrons and extremely large VLDL particles (mmol/l) | 0.086 | 5.09 | **< 0.001** | 0.138 | **0.024** | 0.014 | **0.004** | 0.049 | 0.506 | 0.055 | 0.343 |
| Lipoprotein subclasses | Concentration of very large VLDL particles (mmol/l) | 0.152 | 8.76 | **< 0.001** | 0.112 | **0.016** | 0.016 | **< 0.001** | 0.047 | 0.402 | 0.055 | 0.217 |
| Lipoprotein subclasses | Free cholesterol in chylomicrons and extremely large VLDL (mmol/l) | 0.107 | 6.19 | **< 0.001** | 0.130 | **0.024** | 0.016 | **< 0.001** | 0.051 | 0.451 | 0.048 | 0.378 |
| Lipoprotein subclasses | Free cholesterol in large VLDL (mmol/l) | 0.167 | 9.66 | **< 0.001** | 0.100 | **0.017** | 0.016 | **< 0.001** | 0.038 | 0.448 | 0.047 | 0.240 |
| Lipoprotein subclasses | Free cholesterol in very large VLDL (mmol/l) | 0.160 | 9.22 | **< 0.001** | 0.125 | **0.010** | 0.017 | **< 0.001** | 0.044 | 0.449 | 0.062 | 0.184 |
| Lipoprotein subclasses | Phospholipids in chylomicrons and extremely large VLDL (mmol/l) | 0.096 | 5.62 | **< 0.001** | 0.137 | **0.025** | 0.016 | **0.001** | 0.045 | 0.541 | 0.053 | 0.364 |
| Lipoprotein subclasses | Phospholipids in very large VLDL (mmol/l) | 0.151 | 8.67 | **< 0.001** | 0.131 | **0.012** | 0.017 | **< 0.001** | 0.047 | 0.457 | 0.066 | 0.187 |
| Lipoprotein subclasses | Total lipids in chylomicrons and extremely large VLDL (mmol/l) | 0.080 | 4.74 | **0.001** | 0.130 | **0.031** | 0.014 | **0.005** | 0.037 | 0.607 | 0.071 | 0.218 |
| Lipoprotein subclasses | Total lipids in large VLDL (mmol/l) | 0.133 | 7.62 | **< 0.001** | 0.097 | **0.020** | 0.013 | **< 0.001** | 0.039 | 0.437 | 0.048 | 0.227 |
| Lipoprotein subclasses | Total lipids in very large VLDL (mmol/l) | 0.152 | 8.76 | **< 0.001** | 0.125 | **0.012** | 0.016 | **< 0.001** | 0.045 | 0.443 | 0.069 | 0.145 |
| Lipoprotein subclasses | Triglycerides in chylomicrons and extremely large VLDL (mmol/l) | 0.058 | 3.68 | **0.006** | 0.124 | 0.048 | 0.012 | **0.018** | 0.029 | 0.696 | 0.080 | 0.180 |
| Lipoprotein subclasses | Triglycerides in large VLDL (mmol/l) | 0.114 | 6.56 | **< 0.001** | 0.100 | **0.020** | 0.012 | **< 0.001** | 0.043 | 0.401 | 0.049 | 0.231 |
| Lipoprotein subclasses | Triglycerides in very large VLDL (mmol/l) | 0.137 | 7.87 | **< 0.001** | 0.130 | **0.013** | 0.016 | **< 0.001** | 0.052 | 0.400 | 0.070 | 0.153 |
| Lipoprotein subclasses | Cholesterol in chylomicrons and extremely large VLDL (mmol/l) | 0.125 | 7.20 | **< 0.001** | 0.135 | **0.015** | 0.016 | **< 0.001** | 0.057 | 0.386 | 0.054 | 0.309 |
| Lipoprotein subclasses | Cholesterol in very large VLDL (mmol/l) | 0.182 | 10.61 | **< 0.001** | 0.108 | **0.013** | 0.017 | **< 0.001** | 0.03 | 0.565 | 0.062 | 0.139 |

| | | | | | | | | | | | | |
|---|---|---|---|---|---|---|---|---|---|---|---|---|
| Relative lipoprotein lipid concentrations | Phospholipids to total lipids ratio in small LDL (%) | 0.261 | 16.31 | **< 0.001** | -0.003 | 0.257 | -0.001 | **< 0.001** | -0.01 | **0.006** | 0.0002 | 0.953 |
| Relative lipoprotein lipid concentrations | Cholesteryl esters to total lipids ratio in medium LDL (%) | 0.193 | 11.40 | **< 0.001** | 0.006 | 0.119 | 0.002 | **< 0.001** | 0.001 | 0.850 | 0.002 | 0.486 |
| Relative lipoprotein lipid concentrations | Cholesteryl esters to total lipids ratio in small LDL (%) | 0.305 | 19.98 | **< 0.001** | 0.003 | 0.245 | 0.001 | **< 0.001** | 0.010 | **0.002** | -0.001 | 0.551 |

BMI, Body Mass Index; GDM, Gestational diabetes mellitus; IDL, intermediate-density lipoprotein; LDL, low-density lipoprotein; SSRI, selective serotonin reuptake inhibitor; VLDL, very low-density lipoprotein. p< 0.05 bolded.

**Table S4**. Linear regression model from delivery timepoint with the NMR analysis variable as the dependent variable, body mass index (BMI) and cotinine level as predictors and study group (selective serotonin reuptake inhibitor (SSRI)-case vs. control), gestational diabetes mellitus (GDM) and additional medication use as cofactors.

| **Delivery n =180 (Control = 107, SSRI = 73)** | | | | | | | | | | | | | |
|---|---|---|---|---|---|---|---|---|---|---|---|---|---|
| **Group** | **Variable** | **Overall regression** | | | **SSRI medication** | | **BMI** | | **GDM** | | **Cotinine level (ng/ml)** | | **Additional medication use** |
| | | **Adjusted $R^2$** | **F** | **p** | **B** | **p** | **B** | **p** | **B** | **p** | **B** | **p** | **B** | **p** |
| Fatty acids (ratios) | Ratio of polyunsaturated fatty acids to total fatty acids (%) | 0.060 | 3.29 | **0.007** | -0.015 | **0.028** | -0.001 | 0.242 | -0.012 | 0.133 | 0 | 0.158 | 0 | 1.000 |
| Relative lipoprotein lipid concentrations | Triglycerides to total lipids ratio in small LDL (%) | 0.085 | 4.31 | **0.001** | 0.036 | **0.014** | 0.001 | 0.324 | 0.037 | **0.034** | 0.0006 | 0.117 | 0.006 | 0.674 |
| Relative lipoprotein lipid concentrations | Triglycerides to total lipids ratio in IDL (%) | 0.114 | 5.60 | **< 0.001** | 0.033 | **0.021** | 0.002 | 0.045 | 0.035 | **0.039** | 0.001 | 0.081 | -0.003 | 0.802 |

IDL, intermediate-density lipoprotein; LDL, low-density lipoprotein; SSRI, selective serotonin reuptake inhibitor; VLDL, very low-density lipoprotein. p< 0.05 bolded.

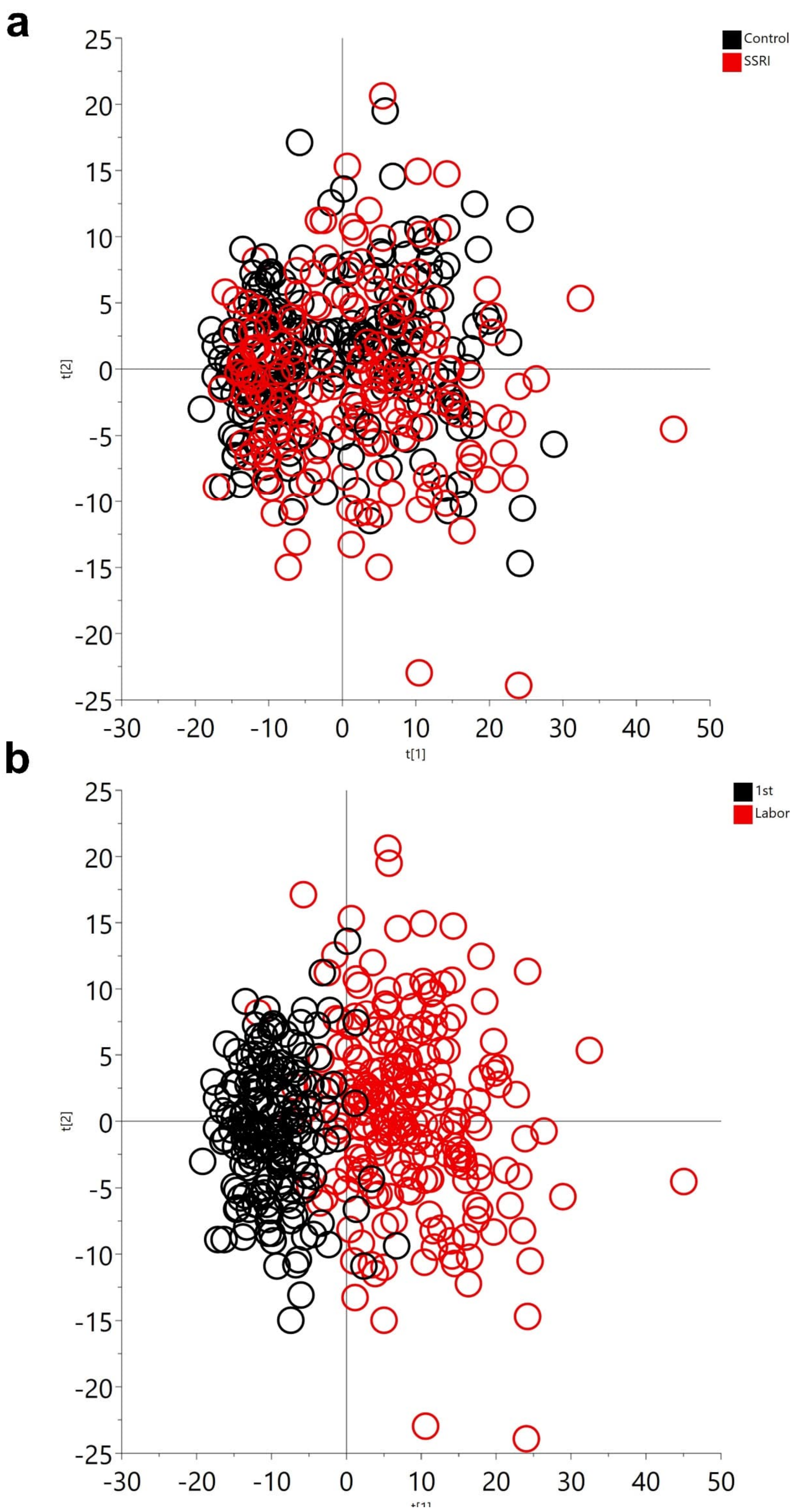


**Fig S1** Principal component analysis (PCA) showing patterns of a metabolic profile between (a) SSRI users (n = 122) and controls (n = 117) and (b) 1st trimester and delivery timepoints of pregnancy (n= 239)

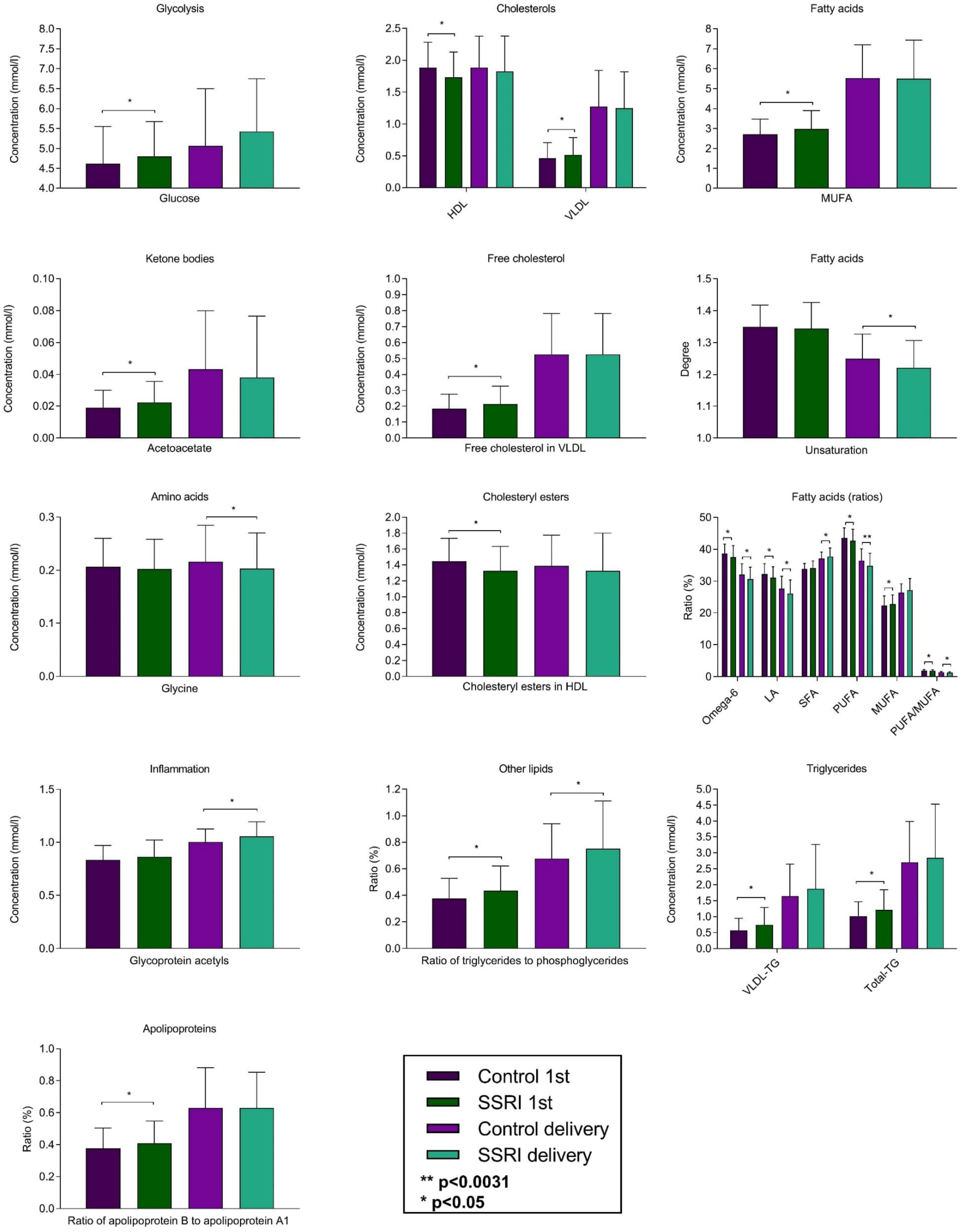


**Fig S2** Comparison of the variable levels from the first trimester and delivery timepoints of pregnancy between SSRI-users (n=122) and control women (n=117). Wilcoxon rank-sum test was used to compare the variable levels between groups. Significant differences at the multiple testing corrected α-level ($p < 0.0031$) and the nominal p-value < 0.05 are represented. Error bars represent the median value and interquartile range (IQR). HDL, high-density lipoprotein; LDL, low-density lipoprotein; MUFA, Monounsaturated fatty acids; Omega-6, ratio of omega-6 fatty acids to total fatty acids; LA, ratio of linoleic acid to total fatty acids; SFA, saturated fatty acids; PUFA, ratio of polyunsaturated fatty acids to total fatty acids; MUFA %, ratio of monounsaturated fatty acids to total fatty acids; PUFA/MUFA, ratio of polyunsaturated fatty acids to monounsaturated fatty acids; VLDL-TG, Triglycerides in VLDL; Total TG, Total triglycerides

### Lipoprotein subclasses

### Relative lipoprotein lipid concentrations

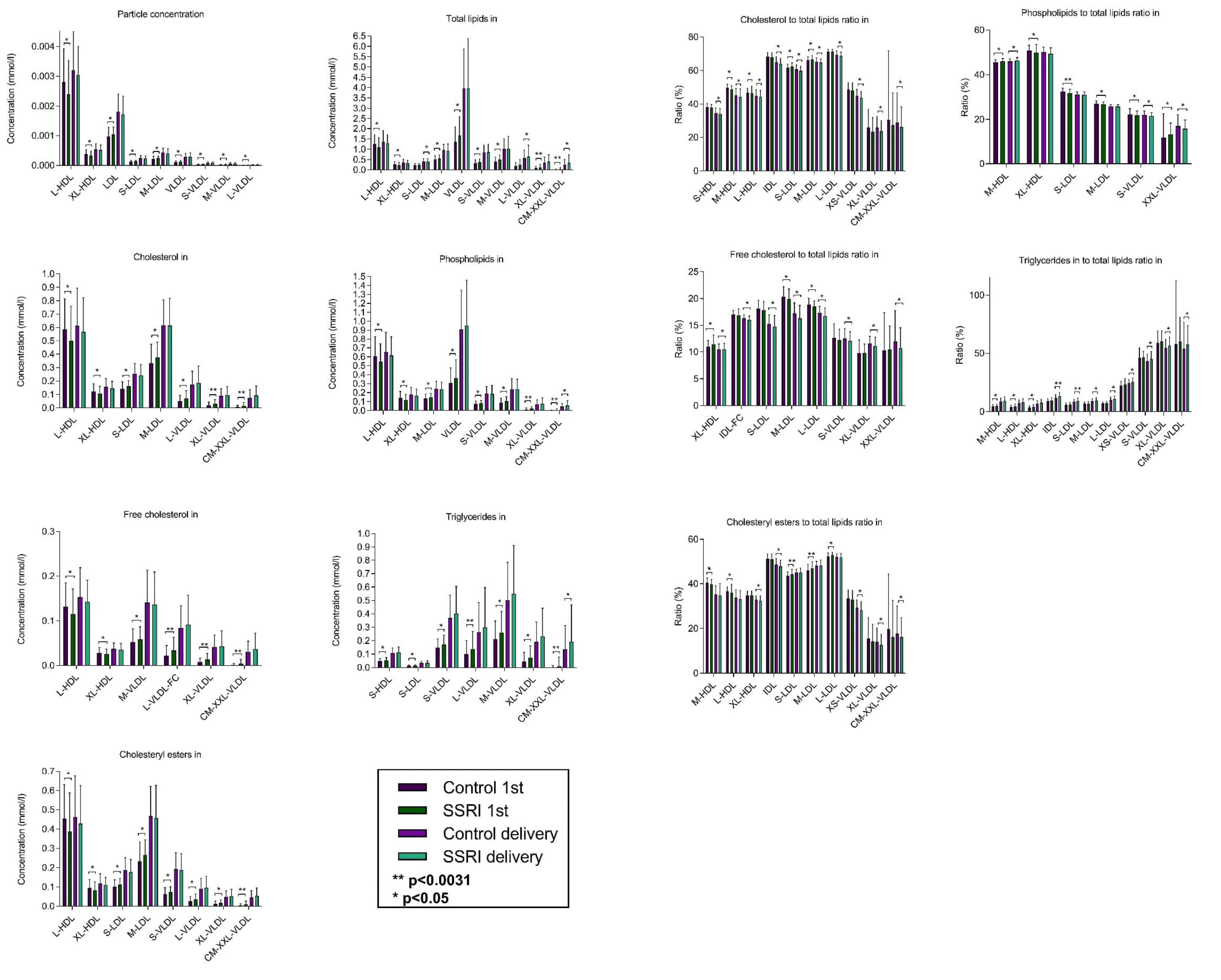


**Fig S3** Comparison of the variable levels from the first trimester and delivery timepoints of pregnancy between SSRI-users (n=122) and control women (n=117). Wilcoxon rank-sum test was used to compare the variable levels between groups. Significant differences at the multiple testing corrected α-level ($p < 0.0031$) and the nominal p-value < 0.05 are represented. Error bars represent the median value and interquartile range (IQR). XS, very small; S, small; M, medium; L, large; XL, very large; XXL, extremely large; CM, chylomicron; VLDL, very low-density lipoprotein; LDL, low-density lipoprotein; IDL, intermediate-density lipoprotein; HDL, high-density lipoprotein

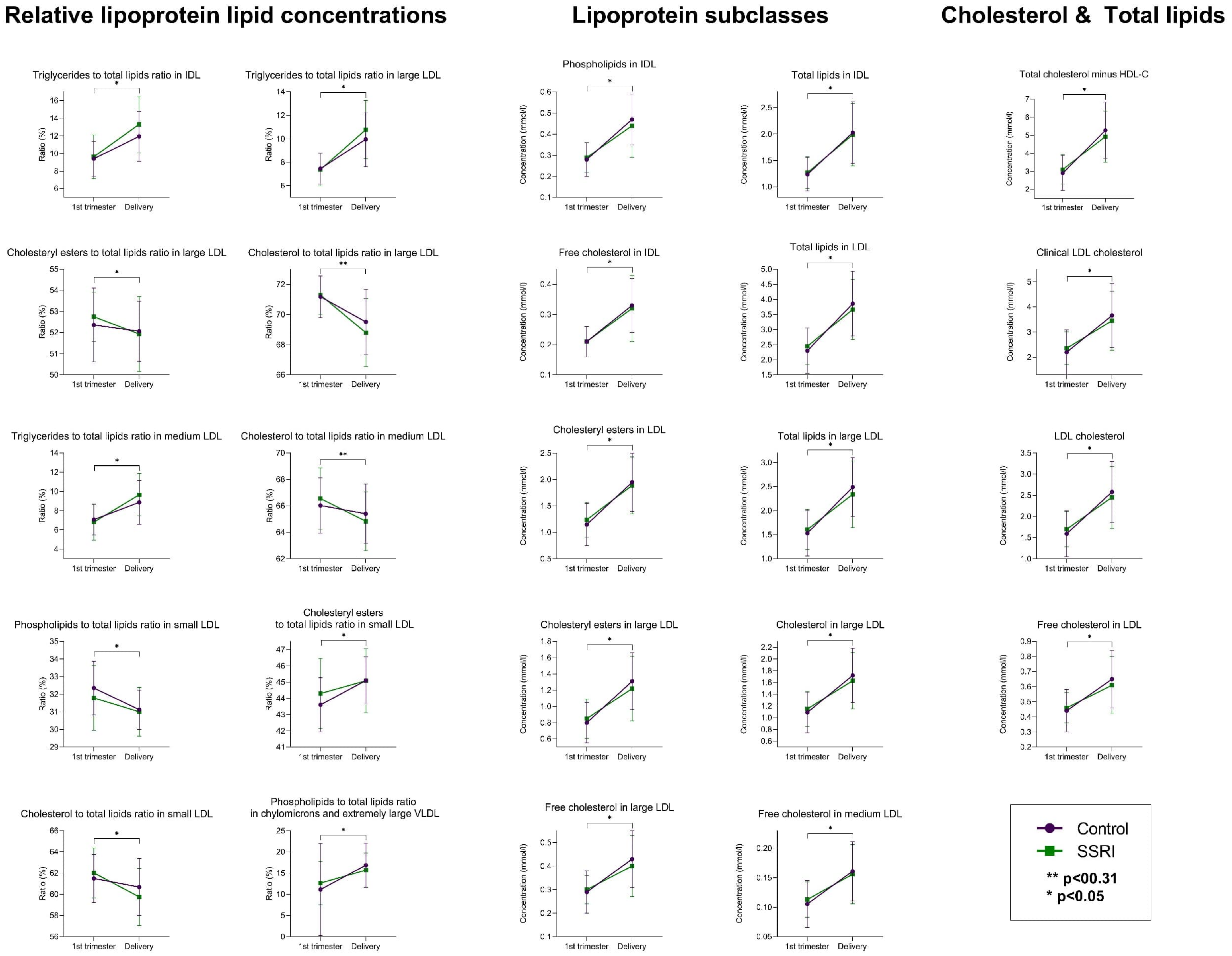


**Fig S4** Comparison of the changes on the metabolic variables during pregnancy between SSRI-users (n=37) and control women (n=113).

Wilcoxon rank-sum test was used to compare the metabolite levels between groups (n = 150). Significant differences with the nominal p-value < 0.05 are represented.

Error bars represent the median value and interquartile range (IQR)

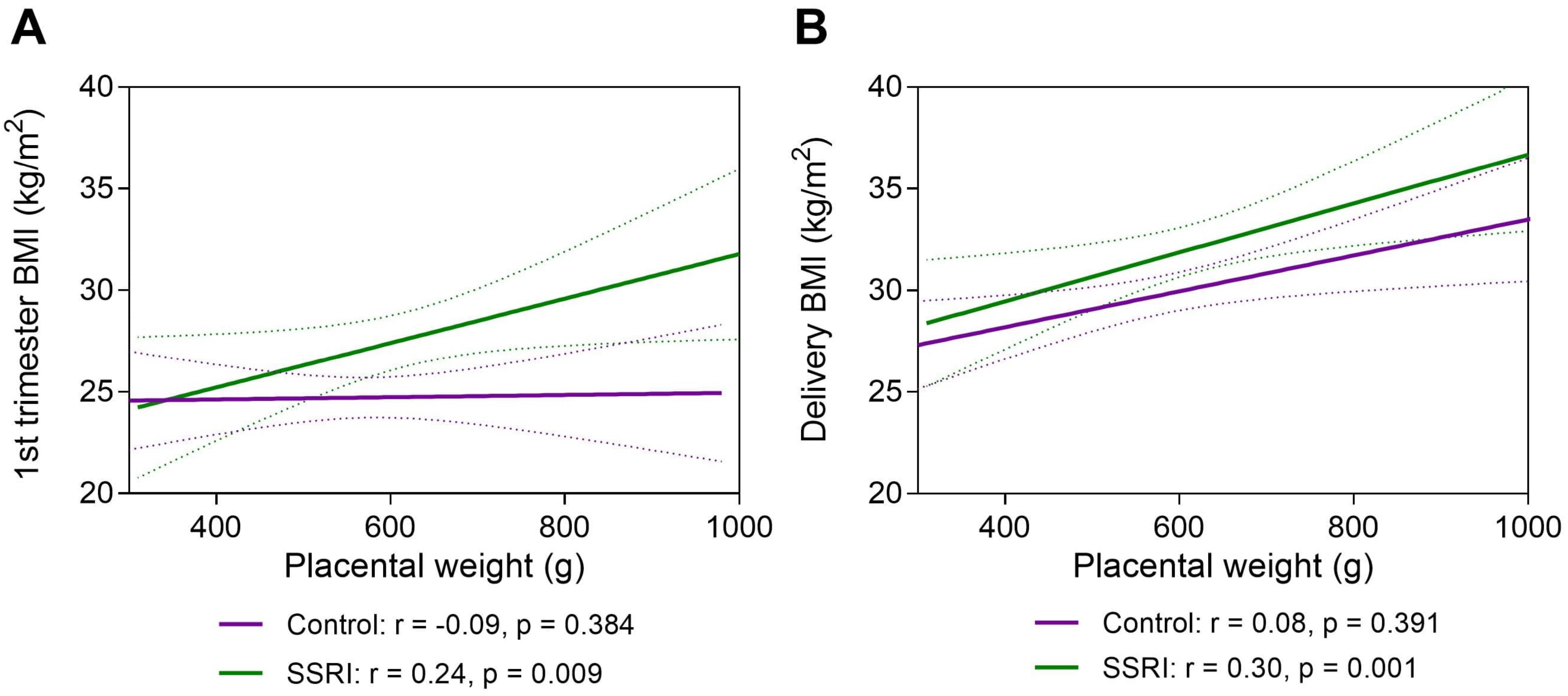


**Fig S5** Spearman's correlation with 95 % confidence interval between placental weight and **(**a) 1st trimester body mass index (BMI) and **(**b) BMI at delivery in SSRI users (n = 122) and controls (n = 117)

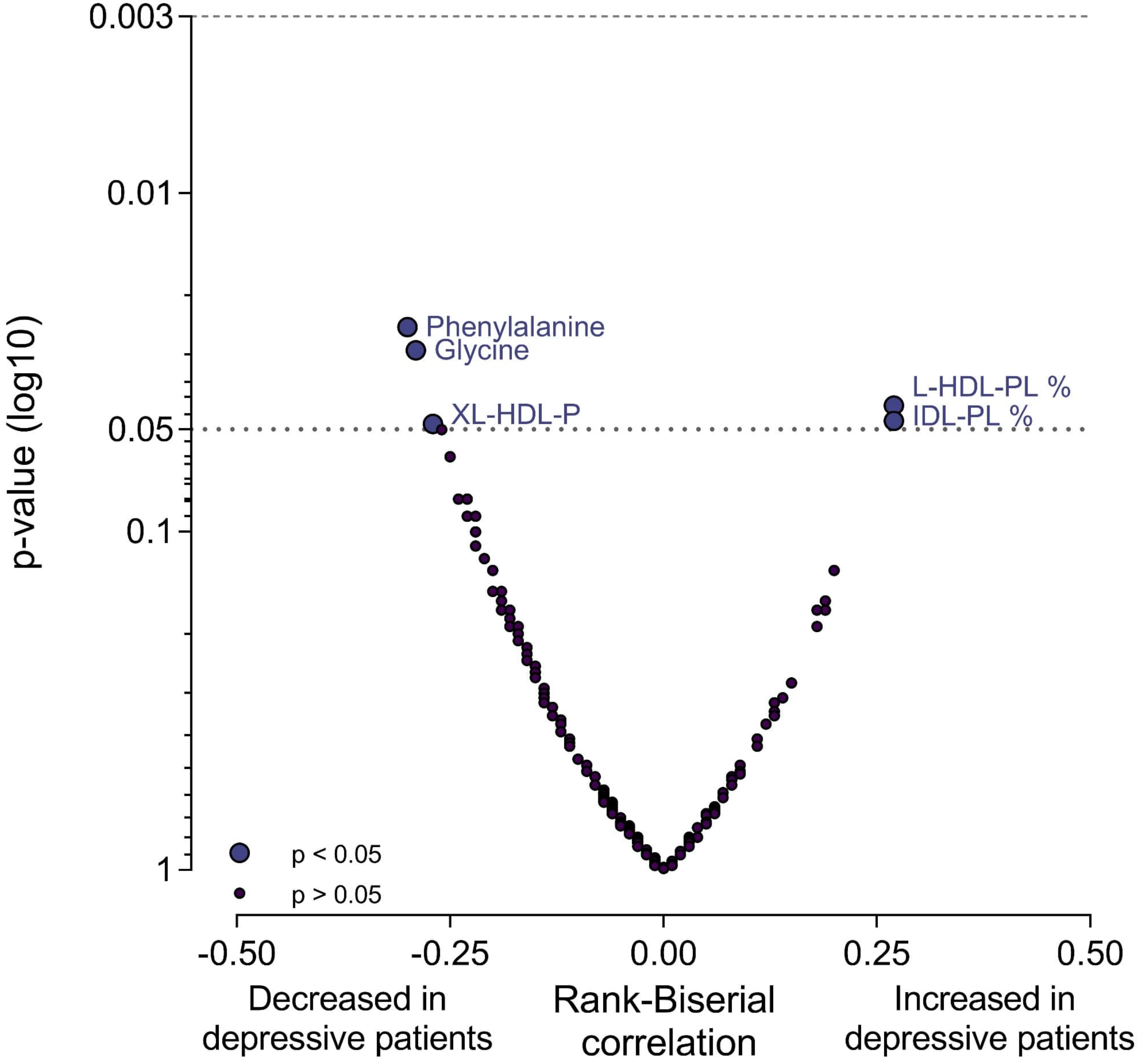


**Fig S6** Volcano plot summarizing the impact of depression on the metabolic variables at delivery timepoint. The significance and magnitude of the observed changes between individuals suffering depression (n =62) and other mental health disorder (n = 28). Wilcoxon rank-sum test was used to compare the variables between the subgroups within the SSRI group. The effect is represented as rank-biserial correlation. XL-HDL-P, Concentration of very large HDL particles; L-HDL-PL %, Phospholipids to total lipids ratio in large high-density lipoprotein; IDL-PL %, Phospholipids to total lipids ratio in IDL
